# Screening electrocardiogram in young athletes and military members: a systematic review and meta-analysis

**DOI:** 10.1101/2020.09.13.20193706

**Authors:** Aaron Lear, Niraj Patel, Chanda Mullen, Marian Simonson, Vince Leone, Constantinos Koshiaris, David Nunan

## Abstract

**Objective:** To determine the effect of electrocardiogram (ECG) screening on prevention of sudden cardiac arrest and death (SCA/D) in young athletes and military members.

**Data Sources:** MEDLINE, Embase, Cochrane CENTRAL, Web of Science, BIOSIS, Scopus, SPORT discus, PEDro, and clinicaltrials.gov were searched from inception to dates between 2/21/19 and 7/29/19.

**Study Selection:** Randomized and non-randomized controlled trials, where pre-participation examination including ECG was the primary intervention used to screen athletes or military 40 years of age or younger. Accepted controls were no screening, usual care, or pre-participation examination without ECG. 3 published studies, and one conference abstract were identified for inclusion.

**Data Extraction:** In all four studies, risk of bias was assessed with the Cochrane risk of bias tool, and found to be generally high. Two studies had data extracted for random effects meta-analysis, and the remaining study and conference abstract were included in narrative review.

**Data Synthesis:** 4 studies (11,689,172 participants) were included, all at high risk of bias. Pooled data from two studies (n= 3,869,274; very low quality) showed a 42% relative decrease in sudden cardiac death, equating to an absolute risk reduction of .0016%. Uncertainty was high, with a potential 67% relative decrease to a 45% relative increase in those screened with ECG based on 95% confidence intervals (RR 0.58; 95%CI 0.23, 1.45). Heterogeneity was found to be high as measured with I^2^ statistic (71%).

**Conclusion:** There is very low quality evidence ECG screening decreases risk of sudden cardiac death in young athletes and military members. Decisions need to consider evidence that ECG screening could also increase risk of sudden cardiac death based on the findings of meta-analysis.

**PROSPERO Registration:** CRD42019125560

**Key Points:** ECG screening of athletes has been shown to be more effective than history and physical examination alone to diagnose conditions which put the athlete at risk for sudden cardiac arrest or death (SCA/D). Few data are available to answer the question of the effectiveness of ECG screening in preventing SCA/D in young athletes.

We identified only four published accounts (3 full papers and one conference abstract) of non-randomized trials reporting on the effectiveness of ECG screening to prevent SCA/D in young athletes and military members. The quality of the published evidence is judged to be of very low quality to answer the question of whether ECG screening prevents episodes of SCA/D. No difference was identified between screened and non-screened athletes in data synthesis of two of the published articles eligible for meta-analysis (RR 0.58; 95%CI 0.23, 1.45).

## Introduction

Efforts to reduce the incidence of sudden cardiac arrest and death (SCA/D) in young athletes has led the European Society of Cardiology ^1^ to recommend electrocardiogram (ECG) screening as part of a pre-participation examination (PPE) of young competitive athletes prior to participation in 2005, and updates have confirmed their belief in screening with ECG^2^. Professional bodies around the world have followed this recommendation with statements of their own. With some in agreement including the International Olympic Committee ^3^; some agreeing but with limitations, such as the Australasian Society for Sports Physicians ^4^; while organizations in the United States have resisted calls for blanket screening ^5,6^. The evidence-base to support inclusion of ECG screening for reducing incidence of SCA and SCD in young athletes has not undergone systematic review. A previous systematic review assessed the effectiveness of ECG screening to detect potentially lethal cardiac disorders, but did not address the impact on SCA, SCD and the potential negative effects of ECG screening ^7^.

The provision of a systematic summary of existing data on the outcomes of ECG screening will provide both the public, health care practitioners, and policy makers with vital information about the health effects of ECG screening in these populations when compared with history and physical examination alone.

The aim of this study was to review all available evidence assessing the effect of the addition of ECG screening as part of PPE in young athletic, and military populations on incidence of SCA/D, and to synthesize available research to evaluate the effect of the addition of ECG on the occurrence of SCA/D.

## Methods

This review is part of a project with two objectives: identifying the global incidence of SCA/D in athletes and military members^8^, and evaluating the effect of screening ECG on SCA/D in the same population. It was performed according to the Preferred Reporting Items for Systematic Reviews and Meta-Analyses (PRISMA)^9^ guidelines, and was registered at PROSPERO March 18^th^ 2019 under CRD42019125560 ^10^.

## Data Sources and Searches

The search strategy was designed in conjunction with a medical librarian experienced in systematic reviews (MS), and the search strategy used combined the dual objectives into a single search. The search strategy is included in the supplementary appendix. We searched MEDLINE, Embase, Cochrane CENTRAL, Web of Science, BIOSIS, Scopus, SPORT discus, PEDro, between 2/22/19 and 3/1/19 and Clinicaltrials.gov on 7/29/19. Review articles and position statements were reviewed for eligible articles ^11–13^. There was no limitation on language or date of publication.

### Study Selection

Studies eligible for inclusion were randomized and non-randomized controlled trials, where pre-participation examination including ECG was the primary intervention used to screen athletes or military 40 years of age or younger. Accepted controls were no screening, usual care, or pre-participation examination without ECG. Age 40 was selected due to the increased incidence of coronary artery disease as cause of SCA/D with increasing age, and the desire to focus on etiologies other than coronary artery disease ^11,14^.

The pre-specified primary outcome was the difference in SCA/D in athletes and military populations screened with ECG compared to control groups not screened with ECG. Secondary outcomes were planned but not carried out due to the lack of existing data in the literature. Details on these outcomes are listed in the supplementary appendix.

### Data Extraction and Quality Assessment

Independent dual-investigator article screening, selection, risk of bias (ROB) assessments, and extraction was performed with Covidence (Covidence.org, Veritas Health Innovation, Melbourne, Australia 2018). The primary author (AL) screened all titles, and the second reviewer was from a team of three (CM, NP, VL). The Cochrane risk of bias tool ^15^, native to Covidence was used for ROB assessment. Disagreements were resolved by discussion with primary author (AL), and the second reviewer.

### Data Synthesis and Analysis

Meta-analysis was performed with the statistical package native to Review Manager (Cochrane Collaboration, London, UK) using the random-effects Mantel-Haenszel method based on the clinical heterogeneity within the included studies^16^. Data are presented as relative risk with 95% confidence interval in those screened with ECG compared with those not screened with ECG where possible. Heterogeneity is reported with summary statistics *I*^2^ and Chi^2^, with pre-specified values of <30% considered low; 30-70% considered moderate; and >70% considered high. A p-value of 0.10 or lower for Chi^2^ statistic was indicated statistical heterogeneity. Sensitivity analysis and publication bias assessment (e.g. funnel plot asymmetry) were planned but not performed due the small number of included studies.

## Ethical Approval

Ethical approval was not necessary as only publicly available data was included in this review.

## Role of the Funding Source

No funding was received for this project.

## Results

After removal of 10,780 duplicates, and addition of a further 11 titles after hand search, 20,059 titles and abstracts were screened from database searches, and additions from review of recent review articles. Full text screening was carried out on 322 articles, four of these met criteria for inclusion including three published articles, and one conference abstract. (Figure 1).

**Figure 1:**
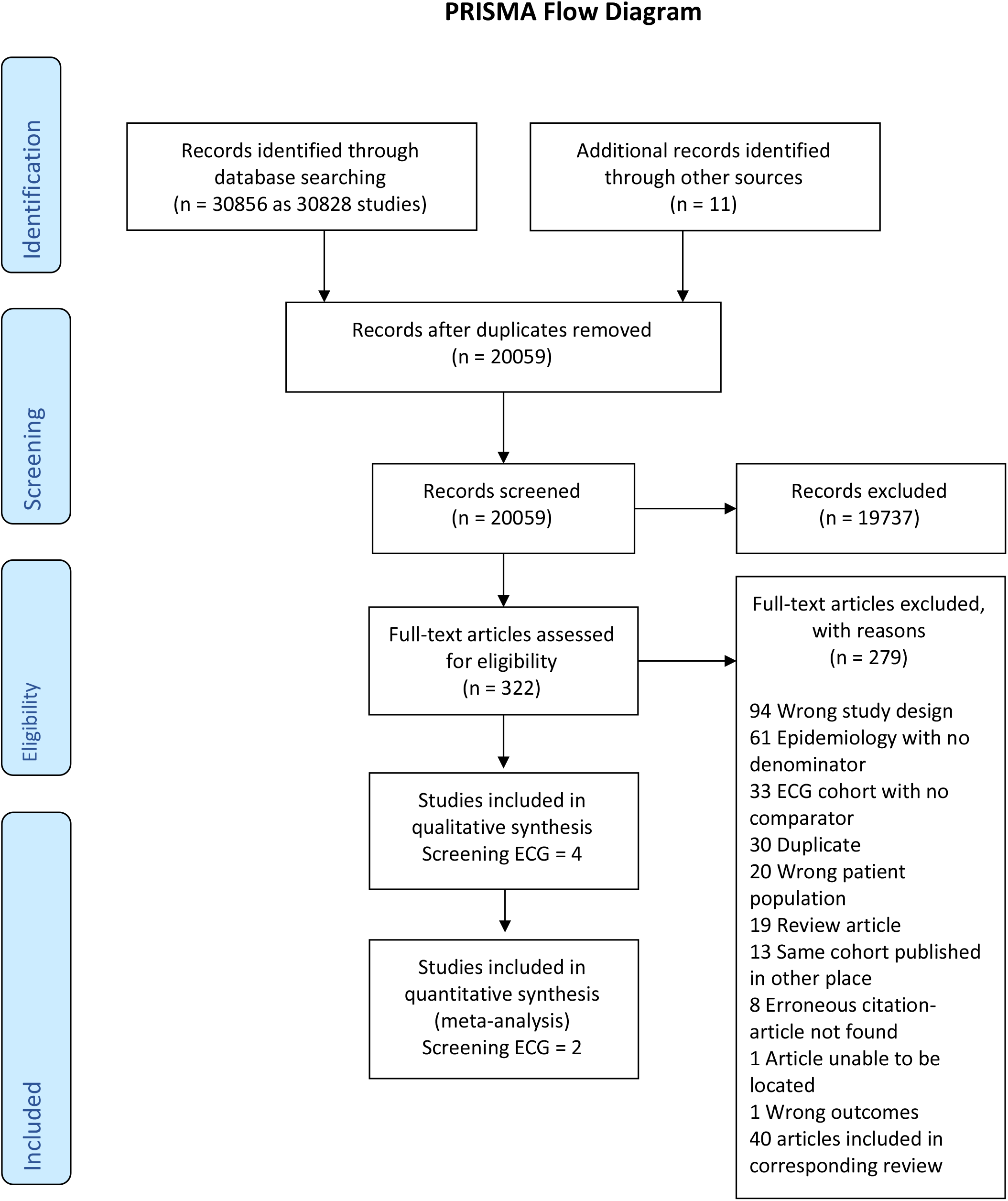
Preferred Reporting Items for Systematic Reviews and Meta-Analyses (PRISMA) flow sheet

### Included studies

The three studies and single conference abstract included were all non-randomized controlled studies. The three articles were on athletes^17–19^ and included a total of 6,431,380 subjects, and the included conference abstract was in military males only^20^ including 5,257,792 subjects. Two studies, and the conference abstract included a historical control group, comparing the rate of SCD in a control cohort prior to the implementation of ECG screening, to an intervention cohort after ECG screening began ^17,19,20^. The third included study compared two separate previously published cohorts of athletes, one that had not been screened with ECG as part of their PPE, and one that had been screened with ECG^18^. No studies were identified which reported an outcome of SCA.

Only two studies were included in meta-analysis, both on athletes^17,18^. In both studies included in meta-analysis it was unclear if the historical control group received a screening PPE, or no examination^17,18^. In those included in the meta-analysis, the intervention group in Corrado, et. al. (2009) received ECG screening as part of their PPEs; and Steinvil, et. al. (2011) included an intervention group that received PPE with ECG, and every four year exercise stress test in those aged 17-34, with annual stress test over this age.

In the two entries not included in the meta-analysis, the conference abstract^20^ did not provide extractable data for meta-analysis; and Maron, et. al. (2009) included a portion the Corrado, et. al. (2006) cohort in their analysis and was left out of meta-analysis to avoid double counting included subjects. The conference abstract including only military males^20^ compared conscripts prior to the initiation of ECG screening, to those after the initiation of ECG screening upon entry to the army. Maron, et. al. (2009) compared the rate of SCD in a cohort who had been screened with PPE including ECG, with the rate in a cohort that received a PPE without ECG. Full descriptions and characteristics of these four studies are presented in the supplementary appendix.

### Risk of Bias Assessment

All included studies were found to have either unknown, or high risk of bias in the large majority of categories evaluated (Table 1). No included studies reported funding which was determined to increase their risk of bias. Further details on the risk of bias determination is included in the characteristics of included studies table in the supplementary appendix.

**Table 1:**
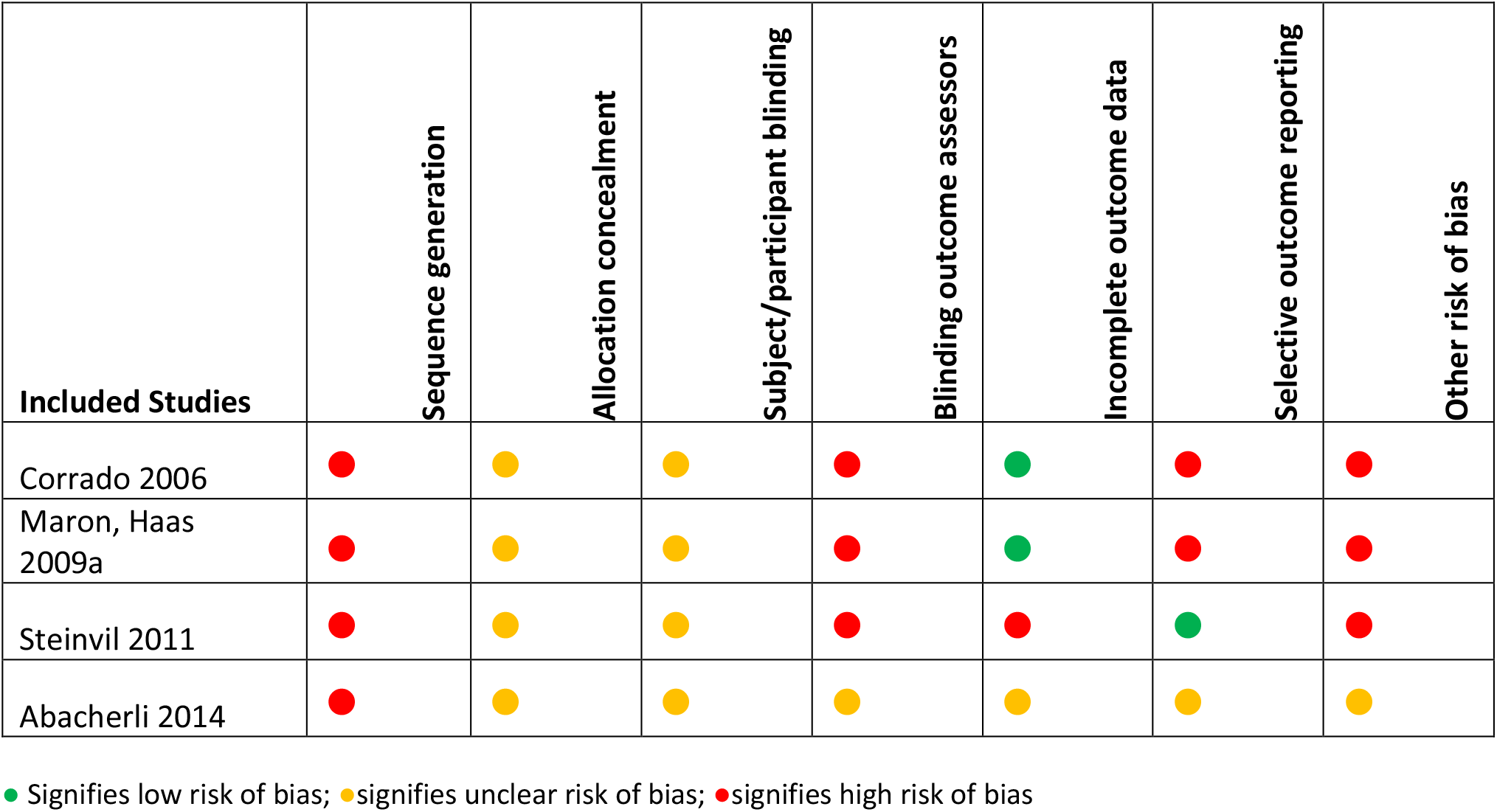
Risk of bias in included non-randomized trials in objective 2; based on Cochrane risk of bias tool

### Effectiveness of ECG screening

Corrado, et. al. (2006) reports an analysis on SCD in athletes in the Veneto region of Italy before and after the initiation of a mandated ECG screening program. The authors report a primary outcome of an 89% decrease in incidence of SCD after the implementation of ECG screening program for athletes in 1982. This is done by comparing a two-year time period prior to the initiation of screening (1979-80), to the final two years of the screening period (2003-04). In our analysis, we compared the data from 23 years of athletes screened with ECG compared with the three years of athletes not screened with ECG showed a 63% decreased risk in the ECG screened group (RR 0.37; 95%CI 0.20 to 0.69).

Steinvil, et. al. (2011) compares SCD events in athletes reported in two Israeli newspapers covering 90% of the country, before and after the initiation of a cardiac screening program. The required intervention began in 1997, and the authors compared media reports in the newspapers for the 12 years before 1997, to the 12 years after the initiation of the law. The authors showed a non-significant 5% decrease in risk in those athletes undergoing cardiac screening with ECG and stress test, to those which did not receive this screening (RR: 0.95; 95%CI 0.43 to 2.13). The findings were highly uncertain, with a potential 57% decrease in risk of SCD to 113% increased risk noted in the 95% confidence interval.

A conference abstract by Abacherli, et. al. (2014) details a comparison of SCD in Swiss male military conscripts separated into age groups 16-19, 20-14, 25-29. The authors compare episodes of SCD after the initiation of ECG screening, compared to historical controls prior to ECG screening. A statistically significant reduction in the ECG screened 20-24 age group with a point estimate of 0.56 (CI: 95% 0.35 to 0.91) was reported. The same comparison in men aged 16-19 was found to be 0.89, and 25-29 was found to 1.04. These were described as non-significant, with only the point estimates, and no confidence intervals reported. The abstract is unclear as to whether the statistical method used was relative risk or odds ratio, making interpretation of the findings difficult. No extractable data were present. Contact with the author revealed that there was no full text article produced from this data, and the authors were unable to share the data at the time of contact.

Maron, et. al. (2009), in comparing a cohort of U.S. athletes in the state of Minnesota who have undergone PPE without ECG screening, with a proportion of ECG screened athletes from the Corrado, et. al. (2006) Italian cohort over a similar time period, report a 6% decrease in risk of SCD but estimates were also uncertain with a potential 59% decrease in risk to a 112% increase in risk in the 95% confidence interval (RR 0.94; 95%CI 0.41, 2.12).

Two studies ^17,19^ including athlete participants were included in meta-analysis (Figure 2). The results show a non-significant relative decrease risk of 42% for SCD in athletes screened by ECG but uncertainty was high, with a potential 77% relative decrease to a 45% relative increase in those screened with ECG (RR 0.58, 95%CI 0.23 to 1.45; I^2^ = 71%; Chi^2^ 3.41, p=0.06). The heterogeneity present in the analysis was high by both I^2^, and Chi^2^ methods.

**Figure 2:**
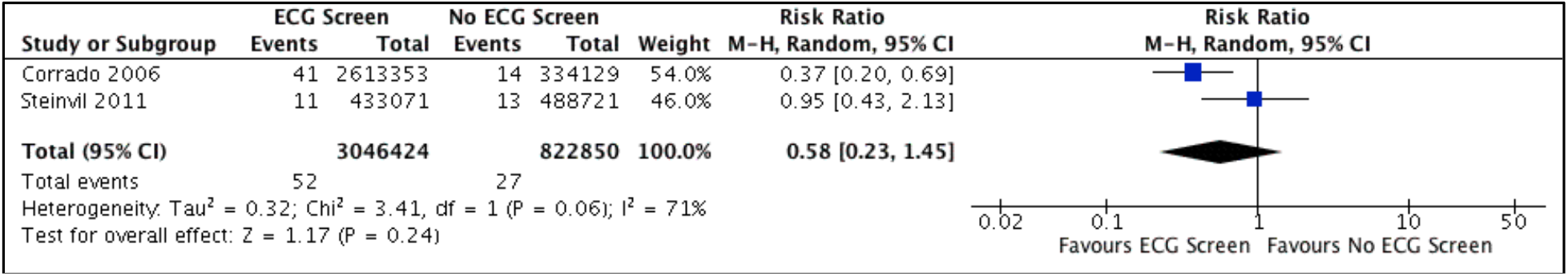
Meta-analysis of studies reporting data on outcomes of sudden cardiac death in athletes screened with ECG (experimental), compared to athletes not screened with ECG (control)

Corrado, et. al. (2006) reported data beyond the outcome of SCD, and this was done only in a portion of the intervention cohort, not allowing comparison with the control group. The authors report that 9% (3,914 of 42,386) of athletes were referred for further testing after their initial ECG screening and a further 2 % (879 of 42,386) were ultimately removed from sport. The authors did not describe the treatment rendered to athletes with positive screening ECG beyond the further diagnostic studies used and did not report on athletes returning to sport after treatment.

### Quality of evidence

For the primary outcome of SCD we judged the evidence to be of very low certainty due mainly to high risk of bias, serious inconsistency, and serious imprecision.

## Discussion

Our systematic, pre-registered, comprehensive, and up-to-date review found very low-quality evidence that ECG screening decreases risk of sudden cardiac death in young athletes and military members. However, caution is needed when considering this finding. Firstly, we were able to perform meta-analysis only with two studies. The absolute risk reduction from pooling these studies was 0.00157%. Using a single year assessment^21^, this would result in a number needed to screen to prevent one death in one year of 63,694. Secondly, only one of the four studies included reached statistical significance when evaluating the effect of ECG screening on SCD, and the remaining studies report confidence intervals which include both considerable decreased and increased risk with ECG screening. Thirdly, the findings of the included conference abstract are based on unpublished data^20^. And fourth, due to high risk of bias, high heterogeneity and poor precision of effect estimates, the overall certainty of evidence on the effectiveness of ECG screening was judged to be very low. Taken together, we have very low confidence that our findings would not change substantially with further high-quality research.

The existing evidence base to support the use of ECG screening to prevent SCD in athletes is largely confined to the data presented in the Corrado, et. al. (2006) article included in the review. There are significant methodologic concerns about this article, including the inherent bias, and likely confounding present, when comparing a small historical control group to a much larger intervention group some 20 years later. There have also been concerns raised about the transparency of the data reported, and further follow up data on the Italian screening program^22^. While there have been no recent controlled studies published, two recently published cohort studies may call into question the ability of screening athletes to prevent SCA/D. Both report on cohorts of mostly male, professional soccer players who underwent ECG screening as part of pre-participation examinations^23,24^. Both reported results with relatively high rates of SCA/D at 6.8, and 63 per 100,000 athlete years compared to accepted estimates of SCA/D in athletic populations. When considering the ability of ECG screening to prevent episodes of SCA/D, it is notable that published data on events of SCA/D suggest that approximately 60% of cardiac conditions which cause SCA/D in athletes may be identifiable with ECG screening^25,26^.

While there remains disagreement, and a general lack of empiric data to support the use of ECG to prevent SCA/D, multiple authors have advocated it’s use as an effective addition to the pre-participation examination due to the ability to better identify conditions putting athletes at risk of SCA/D^27^. A systematic review in 2015^7^ compared the likelihood of history (hx), physical exam (PE), and ECG to identify potentially lethal cardiac disorders reporting the superiority of ECG in sensitivity (ECG=94%; hx 20%; PE 9%), positive likelihood ratio (ECG=14.8; hx=3.22; PE=2.93), and false positive rate (ECG=6%; hx=8%; PE=10%). A recent publication by the National Screening Committee in the United Kingdom^28^ has reviewed available data on ECG screening in athletes and recommended against its use based on the overall low incidence of SCA/D, confirmed by our systematic review^8^, as well as the lack of an effective screening test to identify those at risk of SCA/D. Results in a recent cohort study focusing on collegiate athletes in the U.S. comparing the results history and physical exam to additional ECG screening in the same cohort of patients^29^ show false positive rates of 33.3% for history alone, 2% with physical exam alone, and 3.4% with ECG alone. Sensitivity with ECG was reported as 100% compared to 15.4% for history, and 7.7% for physical exam.

As demonstrated in this review, there remains little trial data to compare information on how ECG screening affects athlete’s removal from sport, follow up treatment, and potential return to sport. There is cohort data on some of these outcomes, the most notable again being from the included Corrado, et. al. (2006) article which reports on a subgroup of the screened athletes included, with 9% found to be abnormal and receiving further testing, ultimately resulting in 2% excluded from sport. A 2014 scientific statement from the American Heart Association^30^ details a selection of published articles on ECG screening cohort studies, with no control groups and community ECG screening projects in athletes 12-25 years old. Articles reviewed report abnormal ECG rates from 2.5-25%, further testing of 2.5-24%, and disqualification from sport rates 0f 0.2-2% of those screening. More modern cohort studies referenced above ^29,31^ comparing history and physical exam to additional ECG screening in the same cohort of patients have shown lower levels of initially positive results in ECG screened athletes to those completing the American Heart Association^32^ history questionnaire, and physical exam alone. It is difficult to compare how these findings affected further testing and treatment, as all subjects received history, physical, and ECG screening interventions. Drezner, et. al. (2016) report the identification of 0.25% of athletes with a serious cardiac condition after full evaluation in collegiate athletes in the United States ^29^. Drezner, et. al. (2016) also report an average of 2.6 days out of sport to evaluate those with ECG abnormalities on screening. It is also notable, that within studies using ECG screening and reporting the cardiac abnormalities identified, that Wolff-Parkinson-White syndrome is often far and away the most frequent finding, often making up the majority of identifiable cardiac conditions considered serious ^29,31,33^. Concerns have been raised about identifying asymptomatic individuals with this condition on ECG and how large an impact on prevention of SCA/D^34^ this may provide.

Over the past decade, refinement of the ECG criteria for diagnosing these potentially lethal cardiac conditions has continued to improve the sensitivity, and decrease the false positive rates^35^. While these advancements in the diagnostic capability of ECG screening have occurred, there have been no controlled trials published on independent cohorts of patients comparing the ability of ECG to PPE with history and physical alone to prevent SCA/D. There is great need for a prospective study which tests the utility of screening ECG to prevent SCA/D in athletes.

Carrying out a prospective study on this topic would be a daunting task, and with the rarity of the condition may not be possible. To undertake such a project, one could consider randomizing clusters of high school and collegiate athletes to ECG screening with PPE compared to PPE. A model such as this may be even more feasible in the military, where large numbers of recruits enter into service every year. It would be possible to randomize subjects in this setting, which could lead to the data needed. Short of these two trials, it should be possible to compare the rates of SCA/D in a cohort study in the U.S. Many universities have transitioned to testing their athletes, and comparing the rates of SCA/D in these universities, to comparable universities who do not screen, would be technically feasible.

We believe the strength of this review lies in the breadth of the search for controlled trials of any kind which report on the ability of screening ECG in athletes or military members to prevent SCA/D. The primary limitation of our review is the low quality of evidence provided by the included studies leading to uncertainty for decision making. The limitations lie in both the paucity of, and the poor quality of the identified research reporting outcomes on SCA/D in our population.

## Conclusion

There is very low-quality evidence ECG screening decreases risk of sudden cardiac death in young athletes and military members. Decisions regarding the use of screening ECG need to consider evidence here that ECG screening could also increase risk of sudden cardiac death. We have very low confidence that our findings would not change substantially with further high-quality research.

## Data Availability

Reasonable request for data will be provided after author contact.

## GRADE Summary of Findings Table

| Does screening ECG in athletes/military members prevent sudden cardiac arrest and death? |  |  |  |  |  |  |  |
| --- | --- | --- | --- | --- | --- | --- | --- |
| **Population:** Competitive athletes participating in an organized sport, and active duty military members age 40 or younger.<br>**Intervention:** Performance of pre-participation examination (PPE) with screening electrocardiogram (ECG) with or without echocardiogram to find conditions which are known to lead to sudden cardiac arrest associated with sports/athletic activity.<br>**Comparison:** No PPE, or PPE without screening ECG<br>**Outcome:** 1) SCA/D in screened vs. those not screened with ECG 2) Rate of athletes/military members removed from sport/activity 3) Rate of athletes/military members with abnormal findings, who underwent treatment as a result of screening 4) The number of treated athletes/military members who returned to sport/activity |  |  |  |  |  |  |  |
| Outcome | Intervention | Number of Studies | Absolute Risk No ECG or Exam with no ECG | Absolute Risk ECG screened | Relative Effect (95% CI) | Number of Participants | Quality of the Evidence* |
| Sudden cardiac death in athletes | ECG in 1 study, ECG + exercise stress test in 1 study | 2 studies evaluating prevention of sudden cardiac death | .00328% | .00171% | 0.58 (0.23, 1.45) | 3,869,274 (2 historically controlled trials) | Very Low |

\*Summary of decisions on quality of the evidence:
**Study design:** Non-randomized historically controlled trials at high risk of bias
**Risk of bias:** Very serious concern. Downgraded for significant risk of bias in these historically controlled trials, including in categories of randomization, allocation, blinding and sequence generation, as well as risk of significant confounding due to the historical nature of the studies.
**Inconsistency:** Serious concern. Based on the differences in the estimate of effect and the high heterogeneity with $I^2$ of 71%, and significant $\text{Chi}^2$ p value of 0.06.
**Indirectness:** No concern. The studies included evaluate the event of interest, in the population of interest.
**Imprecision:** Serious concern. Downgraded for wide confidence intervals despite meeting the optimal information size. With the calculated event rates in these combined studies, a power analysis with $p=0.05$ , and $\text{Beta}=0.80$ , a total of 3,177,808 [1,548,556 per group] is suggested for a randomized trial.
**Publication Bias:** not evaluated due to small number of studies.
**Other factors:** There is likely significant confounding which has occurred in these trials. The confounding present is likely to have increased the effect estimate.

## Notes

### Competing Interest Statement

The authors have declared no competing interest.

### Clinical Trial

Prospero: CRD42019125560

### Funding Statement

No funding was provided for this project

### Author Declarations

Systematic review of literature. IRB exempt.

## References

1. Corrado D, Pelliccia A, Bjørnstad HH, et al. Cardiovascular pre-participation screening of young competitive athletes for prevention of sudden death: Proposal for a common European protocol - Consensus Statement of the Study Group of Sport Cardiology of the Working Group of Cardiac Rehabilitation an. Eur Heart J. 2005;26(5):516–524. doi:10.1093/eurheartj/ehi108

2. Pelliccia A, Sharma S, et al. 2020 ESC Guidelines on sports cardiology and exercise in patients with cardiovascular disease. Eur Heart J. 2020:1–80. doi:10.1093/eurheartj/ehaa605

3. Ingersoll CD. The periodic health evaluation of elite athletes: A consensus statement from the international olympic committee. J Athl Train. 2009;44(5):453. doi:10.4085/1062-6050-44.5.453

4. Exeter D, Kuah D, Carbon R, Shawdon A, Bolzonello D. Australasian College of Sport and Exercise Physicians (ACSEP) Position Statement on Pre-Participation Cardiac Evaluation in Young Athletes. https://www.acsep.org.au. Published 2018.

5. Hainline B, Drezner J, Baggish A, et al. Interassociation consensus statement on cardiovascular care of college student-athletes. J Athl Train. 2016;51(4):344–357. doi:10.4085/j.jacc.2016.03.527

6. Maron BJ, Douglas PS, Graham TP, Nishimura RA, Thompson PD. Task Force 1: Preparticipation screening and diagnosis of cardiovascular disease in athletes. J Am Coll Cardiol. 2005;45(8):1322–1326. doi:10.1016/j.jacc.2005.02.007

7. Harmon KG, Zigman M, Drezner JA. The effectiveness of screening history, physical exam, and ECG to detect potentially lethal cardiac disorders in athletes: A systematic review/meta-analysis. J Electrocardiol. 2015;48(3):329–338. doi:10.1016/j.jelectrocard.2015.02.001

8. Lear A, Patel N, Mullen C, et al. Global incidence of sudden cardiac arrest in young athletes and military members: a systematic review and meta-analysis. medRxiv Prepr. 2020;(MEDRXIV/2020/193714). medRxiv 2020.09.13.20193714.

9. PRISMA. http://prisma-statement.org. Published 2009. Accessed July 16, 2019.

10. Lear A. A systematic review to determine the rate of sudden cardiac arrest and death in athletes, and to determine the benefits and harms of the electrocardiogram screening of athletes. PROSPERO. http://www.crd.york.ac.uk/PROSPERO/display_record.php?ID=CRD42019125560. Published 2019.

11. Harmon KG, Drezner JA, Wilson MG, Sharma S. Incidence of sudden cardiac death in athletes: A state-of-the-art review. Br J Sports Med. 2014;48(15):1185–1192. doi:10.1136/bjsports-2014-093872

12. Drezner JA, O’Connor FG, Harmon KG, et al. AMSSM Position Statement on Cardiovascular Preparticipation Screening in Athletes: Current evidence, knowledge gaps, recommendations and future directions.[Erratum appears in Br J Sports Med. 2018 Mar 6; PMID: 29510974]. Br J Sports Med. 2017;51(3):153–167.

13. Mohananey D, Masri A, Desai RM, et al. Global Incidence of Sports-Related Sudden Cardiac Death. J Am Coll Cardiol. 2017;69(21):2672–2673. doi:10.1016/j.jacc.2017.03.564

14. Drezner JA, Sharma S, Baggish A, et al. International criteria for electrocardiographic interpretation in athletes: Consensus statement. Br J Sports Med. 2017;51(9):704–731. doi:10.1136/bjsports-2016-097331

15. Higgins JP, Green S, eds. Cochrane Handbook for Systematic Reviews of Interventions. 5.1.0. Cochrane Collaboration; 2011. www.handbook.cochrane.org.

16. Cochran Collaboration. Review Manager (RevMan). 2014.

17. Steinvil A, Chundadze T, Zeltser D, et al. Mandatory electrocardiographic screening of athletes to reduce their risk for sudden death proven fact or wishful thinking? J Am Coll Cardiol. 2011;57(11):1291–1296.

18. Maron BJ, Haas TS, Doerer JJ, Thompson PD, Hodges JS. Comparison of U.S. and Italian experiences with sudden cardiac deaths in young competitive athletes and implications for preparticipation screening strategies. Am J Cardiol. 2009;104(2):276–280.

19. Corrado D, Basso C, Pavei A, Michieli P, Schiavon M, Thiene G. Trends in sudden cardiovascular death in young competitive athletes after implementation of a preparticipation screening program. JAMA. 2006;296(13):1593–1601.

20. Abacherli R, Schmid R, Kobza R, Franz F, Schmid J, Erne P. Pre-paricitation ECG screening preventing SCD-insight from the Swiss Army. Hear Rhythm. 2014;11(5):S224.

21. Rembold C. Number needed to screen: development of a statistic for disease screening. BMJ. 1998;317:307–312.

22. Van Brabandt H, Desomer A, Gerkens S, Neyt M. Harms and benefits of screening young people to prevent sudden cardiac death. BMJ. 2016;353(April):1–5. doi:10.1136/bmj.i1156

23. Malhotra A, Dhutia H, Finocchiaro G, et al. Outcomes of Cardiac Screening in Adolescent Soccer Players. N Engl J Med. 2018;379(6):524–534.

24. Berge HM, Andersen TE, Bahr R. Cardiovascular incidents in male professional football players with negative preparticipation cardiac screening results: an 8-year follow-up. Br J Sports Med. October 2018:bjsports-2018–099845. doi:10.1136/bjsports-2018-099845

25. Harmon KG, Asif IM, Maleszewski JJ, et al. Incidence and Etiology of Sudden Cardiac Arrest and Death in High School Athletes in the United States. Mayo Clin Proc. 2016;91(11 PG-1493-1502):1493–1502.

26. Maron BJ, Haas TS, Murphy CJ, Ahluwalia A, Rutten-Ramos S. Incidence and causes of sudden death in U.S. college athletes. J Am Coll Cardiol. 2014;63(16):1636–1643. doi:10.1016/j.jacc.2014.01.041

27. Drezner JA, Harmon KG, Asif IM, Marek JC. Why cardiovascular screening in young athletes can save lives: a critical review. Br J Sports Med. 2016:bjsports-2016-096606. doi:10.1136/bjsports-2016-096606

28. Couper AK, Poole K, Bradlow W, Field R, Perkins GD, Royle P. Screening for cardiac conditions associated with sudden cardiac death in the young External review against programme appraisal criteria for the UK National Screening Committee Version?: FINAL About the UK National Screening Committee (UK NSC). 2019;(October).

29. Drezner JA, Owens DS, Prutkin JM, et al. Electrocardiographic Screening in National Collegiate Athletic Association Athletes. Am J Cardiol. 2016;118(5):754–759. doi:10.1016/j.amjcard.2016.06.004

30. Maron BJ, Friedman RA, Kligfield P, et al. Assessment of the 12-lead electrocardiogram as a screening test for detection of cardiovascular disease in healthy general populations of young people (12-25 years of age): A scientific statement from the american heart association and the american colleg. J Am Coll Cardiol. 2014;64(14):1479–1514. doi:10.1016/j.jacc.2014.05.006

31. Williams EA, Pelto HF, Toresdahl BG, et al. Performance of the American Heart Association (AHA) 14-Point Evaluation Versus Electrocardiography for the Cardiovascular Screening of High School Athletes: A Prospective Study. J Am Heart Assoc. 2019;8(14):1–9. doi:10.1161/JAHA.119.012235

32. Maron BJ, Thompson PD, Ackerman MJ, et al. Recommendations and considerations related to preparticipation screening for cardiovascular abnormalities in competitive athletes: 2007 update: a scientific statement from the American Heart Association Council on Nutrition, Physical Activity, and Metabol. Circulation. 2007;115(12):1455–1643.

33. Fudge J, Harmon KG, Owens DS, et al. Cardiovascular screening in adolescents and young adults: A prospective study comparing the Pre-participation Physical Evaluation Monograph 4th Edition and ECG. Br J Sports Med. 2014;48(15):1172–1178. doi:10.1136/bjsports-2014-093840

34. Maron BJ, Thompson PD, Maron MS. There is No Reason to Adopt ECGs and Abandon American Heart Association/American College of Cardiology History and Physical Screening for Detection of Cardiovascular Disease in the Young. J Am Heart Assoc. 2019;8(14):1–4. doi:10.1161/JAHA.119.013007

35. Drezner JA, Sharma S, Baggish A, et al. International criteria for electrocardiographic interpretation in athletes: Consensus statement. Br J Sports Med. 2017;51(9):704–731.

