## Supplemental appendix for "Screening electrocardiogram in young athletes and military members: a systematic review and meta-analysis"

### Supplementary Appendix

### Global incidence of sudden cardiac arrest in young athletes and military members: a systematic review and meta-analysis

Aaron Lear MD, MSc; Cleveland Clinic Akron General
Niraj Patel, DO HeyDoctor by GoodRx
Chanda Mullen, PhD; Cleveland Clinic Akron General
Marian Simonson, MS; Cleveland Clinic Foundation
Vince Leone, MD; Northeast Ohio Medical University (NEOMED)
Constantinos Koshiaris, PhD; University of Oxford
David Nunan, PhD; University of Oxford

Contact author:
Aaron Lear


**Table of contents online appendix global incidence of sudden cardiac arrest in young athletes and military members: a systematic review and meta-analysis**

Appendix A: Original written protocol 4

Appendix B: Search strategy 14

Appendix C: Custom risk of bias tool for prevalence studies 23

Appendix D: Deviations from Protocol 26

Appendix E: Characteristics and ROB of included studies (alphabetical) 28

Appendix F: Notable excluded studies 138

Appendix G: Subgroup analysis 139

Figures and tables:

- Table S2: Included studies with SCD by race as categorized by authors 147
- Table S3: Individual sports extracted from included studies. 153
- Figure S1: Incidence of SCD in athletes ≤25 years of age in included studies 139
- Figure S2: Incidence of SCA in athletes ≤25 years of age in included studies 150
- Figure S3: Incidence of SCD in male athletes/military in included studies 141
- Figure S4: Included studies reporting SCA in male athletes 142
- Figure S5: SCD in female athletes reported by included studies 142
- Figure S6: Included studies reporting SCA in female athletes 143
- Figure S7: Incidence of SCD in Elite athletes in included studies 144
- Figure S8: Included studies reporting SCA in elite athletes 144
- Figure S9: Included studies reporting SCD in university athletes 145
- Figure S10: Incidence of SCD in scholastic athletes in included studies 146
- Figure S11: Incidence of SCA in scholastic athletes in included studies 146
- Figure S12: Incidence of SCD in competitive athletes in included studies 147
- Figure S13: Incidence of SCA in competitive athletes in included studies 148
- Figure S14: Incidence of soccer athlete SCD in included studies 150
- Figure S15: Incidence of soccer athlete SCA in included studies 150
- Figure S16: Incidence of SCD in American football players in included studies 151
- Figure S17: Incidence of SCA in American football players in included studies 151

Appendix H: Assessment of publication bias 153
Figure S22: Funnel plot for publication bias of included incidence studies 154
References 155

### Appendix A: Original Written Protocol

**Global incidence of sudden cardiac arrest in young athletes and military members**
**PECO**
**Population:** Competitive athletes participating in an organized sport, and active duty military members age 40 or younger.
**Exposure:** Competitive athletes, or active military members
**Comparison:** No comparison
**Outcome:** Population rate of sudden cardiac arrest and death

#### Types of studies

Studies eligible for the rate of SCA/D portion of the review will include any prospective, or retrospective cohort study; randomized controlled trials (RCT), or other comparative trials (controlled or otherwise) which report a rate of sudden cardiac arrest or death in the screened, or unscreened population of athletes or military service members.

##### Types of participants

We will include athletes 40 years of age or younger, to include active duty military members. For this review, we will consider competitive athletes those participating in athletics of any form or level, including scholastic, recreational, club, collegiate/university level, and professional. We will consider subjects to be athletes even if out of season. In settings where the subjects are described as athletes, without a clear description of their sport, or what they activity they regularly participate in, we will include the article. We anticipate this may occur in articles detailing the rate of SCA/D in athletes, particularly if done retrospectively using medical records, or population data.

We will limit the age at 40 years due to the increased likelihood of coronary artery disease becoming more a more prevalent cause of SCA and death once the age of 30 is reached [1,2] and our desire to focus on causes of SCA in younger athletes outside of coronary artery disease. Active duty military members will be included due to the high level of activity that is required of them, and the similar age profile, and interest in preventing SCA/D. In cases of studies which include athletes that extend past age 40, an effort will be made to separate those 40 and younger, including contacting the authors of the paper. However, if this is not possible if the subjects over age 40 comprise 10% or less of the subjects in the paper, we will include it. If there are multiple papers with athletes over 40 years of age included, we will consider sensitivity testing the results to determine their inclusion.

##### Type of outcome measures

Primary outcomes
The primary outcome is the rate of SCA/D in athletes age 40 and under.

Secondary outcomes

Secondary outcomes for both objectives include subgroup analyses on:

1. Subjects 25 years of age or less, and age 26-40 in all categories of primary outcomes.
2. Male and Female subjects in all categories of primary outcomes.
3. Primary outcomes evaluated by race.
4. Primary outcomes evaluated by sport played.
5. Primary outcomes evaluated by level of sport.

**Search methods for identification of studies**

We aim to identify trials and articles which meet the inclusion criteria in any language, published at any time. In cases where unpublished data is located, we intend to contact the authors and include this information if possible. We will use search electronic databases as detailed below, and intend to hand search the bibliographies of articles selected for the review.

**Electronic searches**

Searches will be run in Ovid/Medline, Embase, Cochrane Central, Sports discus, PEDro, Web of Science, Scopus, clinicaltrials.gov, and BIOSIS. All electronic searches will be developed with a medical librarian with experience and specialization in systematic reviews.

**Searching other resources**

The bibliographies of selected articles will be hand searched for relevant citations.

**Data Collection and Analysis**

Search results will be downloaded into Covidence (Covidence.org, Veritas Health Innovation, Melbourne, Australia 2018) for review, and Mendeley desktop citation software (Mendeley version 1.19.2 Mendeley Ltd 2018) for writing and bibliography production. Covidence will be accessed via institutional subscription.

**Selection of studies**

After search results have been entered into Covidence, two separate reviewers will review the abstracts of all studies included in the final results independently. One reviewer (AL) works in the field of sports medicine, and is familiar with the available literature base, while the other reviewer (CM) works in a family medicine research department and is unfamiliar with the literature base on cardiac screening in athletes. Articles not meeting criteria will be eliminated from consideration. Articles initially selected by both authors will have the full texts read by both authors independently. A single author will review the bibliographies of the selected articles, and request relevant abstracts and articles for review. Articles and publications from bibliography and conference abstract searches will also be identified and subject to the same assessment as articles from the original database search. After review of abstracts and reading the agreed upon full text articles, those meeting the inclusion criteria will be selected for inclusion in the review. Disagreements between the two authors will be resolved by consensus, and where consensus cannot be reached, a third reviewer will be involved to settle the dispute. Reasons for exclusion of all full text articles considered will be recorded. A final flow diagram of the search results, full text articles reviewed, and final included articles will be reported.

In cases where multiple reports have been published on one data set, the most complete article will be used, and the others excluded from the review. The reviewers will link the repeat reports after searching, and reviewing available data, and those not used will be excluded from the study. Those excluded will be reported I the final flow diagram as duplicate reports. In the case of multiple articles describing updates to a single cohort are identified, only the most recent article will be used.

**Data Extraction and Management**
A custom data extraction tool will be developed using Covidence ahead of review and data extraction. Our aim is to pre-select the data extraction form, it may be necessary to alter or adjust the form based on studies identified [3]. The data extraction tool will include raw data extraction, risk of bias information, as well as characteristics of the included studies, guided by the Cochrane handbook checklists for non-randomized studies. The characteristics of the study will include data on: whether there was a comparison made; If participants were allocated to groups, how this was done; which parts of the study were prospective; and on what variables the groups (if any) were compared. Two reviewers will independently review and extract data on all articles which are to be fully reviewed. Disagreements will be resolved by discussion between the two reviewers, and in the cases where discussion does not bring a resolution, a third reviewer will settle the disagreement.

**Assessment of risk of bias, and quality in studies included**

ROB-NOS, use of published guidelines on reporting-GATHER/STROBE/STARD; quality (GRADE if possible);

All studies included will be evaluated for the risk of bias. The risk of bias tool will be included on the custom data collection form developed in Covidence. Risk of bias will be assessed by two reviewers independently. Where there is disagreement, the authors will attempt to resolve the disagreement by discussion. If this is not possible, a third author will settle the dispute. Studies will be rated as low, high, or unclear risk of bias. We intend to produce risk of bias summaries for included studies.

For observational studies we will focus on the selection of the cohort or cohorts, whether the cohort is representative of the population of interest, the ascertainment of the outcome data (in this case the occurrence of SCA/D), the comparability of the groups being compared, and outcome assessment. We will also attempt to include an assessment of the how the authors addressed possible confounding within the studies included.

Where areas of unclear bias is present due to lack of reporting, we will make attempts to contact the authors of the included studies.

We will consider a hierarchy in considering the quality of the studies included in the review. In considering the rate of SCA/SCD, prospective cohorts are expected to be the highest level of evidence available; data gathered prospectively but reviewed in a retrospective manner will be considered of higher quality than fully retrospective analyses. It will be considered of high importance how the rate reported in the studies included come to their conclusion about rates of SCA/SCD. This will be particularly notable when considering the denominator included in these studies [2]. Articles which produce a rate using a cohort which has been followed, or in which the authors are able to articulate the athletes under consideration from medical records, records of enrollment such as the National Collegiate Athletic Association (NCAA) or military records will be considered the highest level of evidence. Articles which use records from insurance company, or other reporting databases in which the population of athletes is estimated, or created based on population estimates, will be considered lower quality data. Articles which report SCA/D rates primarily gathered from media reports will be considered the lowest quality evidence.

**Measure of treatment effect**
We will attempt to report a rate of sudden cardiac arrest, and sudden death. These will be reported as events/100,000 athlete years. Based on the results of the search, we may present two separate summary statistics for screened and unscreened populations. We will also attempt to report these data for subgroups including age 25 and under; age 26-40; boys/men, girls/women, race, sport played, and level of sport.

**Unit of analysis issues**
We anticipate presenting a rate of SCA/D per 100,000 athlete years. It is possible that we will find reporting of overall prevalence percentages, annual incidence rates with different denominators. Our intention is to collect the available data, and if possible, transform the reported rates to our intended unit of analysis. Where this is not possible, the article will be presented in narrative form, and not be included in any potential meta-analysis.

**Dealing with missing data**
We will attempt to contact authors regarding missing or unclear outcome data, or for potentially updated but as of yet unpublished datasets.

If there are methodological criteria for the risk of bias assessment which are missing from the studies reviewed, we will make an attempt to contact authors to retrieve this information.

**Assessment of heterogeneity**
We intend to perform a formal measure of heterogeneity. We believe that we will find primarily observational studies, and that they will carry significant differences in their methodologies,

such as prospective cohorts, insurance/medical records data, and review of sport governing bodies' records. We will perform visual inspection of forest plots and discuss heterogeneity based on these findings.

If we are able to produce summary statistics, we intend to report the $I^2$ and $Chi^2$ formal measures of heterogeneity.

We will report $I^2$ and will consider levels of 0-30% as likely not important; 31-70% as moderate heterogeneity; 71-100% as substantial heterogeneity.

We will also report $Chi^2$ statistic if formal meta-analysis is undertaken, and will consider a level of 0.10 as a significant p value.

**Assessment of publication bias**
If we are able to perform an analysis of publication bias with a funnel plot, we will do so for both objective one and two. If there are clear differences between the studies included in the review, such as results of large and small cohorts, it may be necessary to perform publication bias evaluations in subsets for each objective.

**Data Syntheses**
Our intention is to pool data where possible. Based on the state of the evidence, we expect to be able to pool data to come to a summary statistic on the rate of SCA/D in athletes. We will also attempt to provide summary statistics for rate of SCA/D in subgroups.

The decision to pool data for a summary statistic will be made after review of included studies. Our intention is to combine non-randomized (NRS) and randomized studies if possible. This decision will be taken only after considering the differences that exist in the included studies, including in level of bias, quality of the evidence, confounding, and statistical heterogeneity.

Subgroup analysis and investigation of heterogeneity
If, as anticipated, there is heterogeneity discovered in a formal meta-analysis this will be investigated with subgroup analysis. As previously stated, we will perform subgroup analysis by age, gender, race, and by type of sport or military, as well as level of sport, if reported.

It is possible we will discover other sources of heterogeneity once undertaking the review, and may add to, or reduce these categories. Other potential sources of heterogeneity could include the source of data for incidence, such as studies which analyze media reports of SCA/D vs. studies which report on prospective cohorts.

**Sensitivity analysis**
If formal meta-analysis is undertaken, we do anticipate opportunities for sensitivity analysis. As referenced in the previous section, this may include analysis of articles of different sources of data. As examples, these include military data, articles produced from prospective cohorts,

articles produced from insurance or medical records, or articles produced by analysis of media/internet reports.

**Summary of findings tables**

We anticipate producing summary of findings tables in which the GRADE approach will be used. It is our intention to provide a table. The table will serve as our primary summarization of the quality of evidence. If meta-analysis is not possible, this will serve as our primary summary of each objective.

We also anticipate a summary of important excluded articles, with brief explanations as to why the articles were excluded from review.

**Data Extraction Guide:**

The data included on the extraction form is based on the recommendations from the Cochrane Handbook for Systematic Reviews [4] and is proposed to include:

*Identification:*
Study ID created by review authors
Citation details (title, year, journal, etc.)
Type of data: published article, unpublished article, abstract/conference presentation, direct communication with authors, other
Setting
Funding source (if available)
Author contact details

*Eligibility:*
Included/excluded from review
Reason for exclusion

*Methodology:*
Study design

1. Was there a comparison made?
    a. If yes, how were participants allocated/assigned to groups
    b. If yes, on what variables was comparability between groups assessed
2. Which parts of the study were prospective?
3. SCA/D study or Effect of screening study or both

RCT (group vs individual)
CCT (Group vs. individual)
Comparative study (Describe: time based; before/after)
Cohort (retrospective/prospective)
Cross sectional

Length of study

Setting
Diagnostic criteria (ECG, ECHO, diagnosis of SCA/D attributed to heart-how made)
Concerns about bias (Cochrane risk of bias information will be collected based on study design, such as randomized controlled trials, non-randomized controlled trials, comparative studies, cohort studies)

*Subjects:*
Number of subjects
Demographic breakdown if included to include: Age (range, and statistics provided [mean, SD, etc.]), sex, race
Location of study
Level of athlete: unknown, other, club, scholastic, collegiate, professional, recreational,
Type of athlete: team athlete, individual athlete, unknown

*Interventions:*
Total number of intervention groups
For each group:
Type of screening (ECG alone, ECG and echocardiogram, ECG followed by echocardiogram if abnormal, history and physical alone, no screening, etc.)
Criteria used for ECG interpretation (European criteria, Seattle criteria, etc.)

*Results*

1. For Rate of SCA and death:
    a. Prospective/retrospective
    b. Length of study (or period data collected)
        i. Total number of athlete years collected)
    c. Population: true number accounted for with records; estimate based on population and sport participation data;
    d. Data collected from what source:
        i. Medical/service records
        ii. Insurance
        iii. Sport/service database
        iv. Media reports
        v. other
    e. Number of events
        i. SCA
        ii. SCD
        iii. Other death if reported
    f. Rate of events reported
        i. Rate of SCA
        ii. Rate of sudden cardiac death
        iii. Overall death rate if reported
        iv. Number of resuscitations after SCA if reported
    g. Subgroup rates if reported

            i. Race
            ii. Gender
            iii. Age
            iv. Type of athlete/sport
            v. Level of sport

2. For Effects of screening:
    a. Type of study (RCT, CCT, Comparative trial-what type, Cohort)
    b. Intervention groups (if comparative study)
    c. Number of Subjects allocated to each intervention group
    d. Sample size
    e. Missing participants reported
        i. Missing participants calculated
    f. Summary data for each group
        i. 2x2 table for dichotomous outcomes (+/- screen)
        ii. If available
            1. # athletes suffered SCA/D
            2. # of subjects diagnosed with abnormality
            3. # of subjects withheld from sport
            4. # of subjects treated and how (Medicine, Implantable cardiac defibrillator [ICD], electrophysiology procedure such as ablation, etc.)
                a. Number returned to sport after treatment
    g. Means/SD, etc. for continuous outcomes
    h. Data for ordinal or categorical data-such as type of abnormality found
        i. WPW, HCM, Long QT, etc.
    i. Subgroup analysis
    j. Estimate of effect (if any) with confidence intervals, p value
3. Are results collected raw data?
    a. If a comparison between groups was made, was this adjusted, or unadjusted.

*Miscellaneous*
Conclusions
Notable comments from Study authors

References found in bibliography
Comments from reviewer

*Risk of Bias*
**Cohort study Newcastle Ottawa Scale customized:**
**Selection**
1. <u>Representativeness of the exposed cohort</u>
    a. truly representative of the average athlete/military member in the community being described (elite athletes, club athletes, etc.)*

    b. somewhat representative of the average athlete/military member in the community *
    c. selected group of users: i.e. volunteers for community screening
    d. no description of the derivation of the cohort
2. Selection of the non-exposed cohort
    a. drawn from the same community as the exposed cohort *
    b. drawn from a different source
    c. no description of the derivation of the non-exposed cohort
3. Ascertainment of exposure (that the subject is athlete/military member0
    a. secure record (Medical records, military record, team records, etc.)*
    b. Self-report that one is competitive athlete*
    c. media report/population data
    d. no description
4. Demonstration that athlete/military member did not have condition (SCA/D @ Start of study)
    a. yes *
    b. no

**Comparability**

    c. Comparability of cohorts on the basis of the design or analysis
        a) Study attempts to control for important confounders when making comparison (difference in race, sex, age, sport, etc.)*
            1. List items controlled for
        b) Controlling for confounders not appropriate/necessary-list why*
    d. No attempt to address confounders, or control for significant issues when appropriate*

**Outcome**

5. Assessment of outcome
    a. independent blind assessment *
    b. record linkage *
    c. Panel verification/adjudication of outcome*
    d. Self-report
    e. Media report
    f. no description
6. Was follow-up appropriate for outcome to occur (sufficient number of enrollees, and/or sufficient length of time-would expect minimum around 100,000 athlete years)
    a. yes *
    b. no
        a) List number of athlete years
7. Adequacy of follow up of cohorts
    a. complete follow up - all subjects accounted for *
    b. subjects lost to follow up unlikely to introduce bias - small number lost - > 90 % follow up, or description provided of those lost) *
    c. follow up rate < 89% and no description of those lost
    d. no statement regarding follow up

**Cochrane risk of bias tool for RCT, CCT, Comparative trials:**

Allocation

1. Allocation performed:
    a. How was sequence generated?
    b. Random
    c. Other (before/after, etc.):
2. Allocation concealment:
    a. Was allocation concealed until trial begun
    b. How was allocation concealed?
        i. Adequate
        ii. Inadequate
        iii. Not mentioned
3. Baseline imbalance in the groups: yes/no
    a. Causes:
        i. issues with allocation
        ii. Non-randomized trial

Maintenance/Blinding

1. Were participants blinded during intervention?
2. Were trial personnel blinded during intervention?
3. Were outcomes assessors blinded?
4. Was there deviation from intervention that would be different than in practice?
    a. Were these deviations unbalanced between groups?
5. Were any participants analyzed in groups different than originally assigned?
    a. Is this likely to have affected outcome?
    b. Was this unbalanced?
6. Was the intervention carried out as planned?
7. Did the participants adhere to the plan for intervention?

Missing Data

1. Was outcome data available for all or nearly all subjects included in the study?
    a. If there is substantial missing data-is this unbalanced across groups?
2. Was missing data dealt with appropriately (imputation, sensitivity analysis, etc.)
3. Was there evidence or suggestion of selective data reporting?

Other Source of Bias

1. Please list any other possible source of bias.

### Appendix B: Search Strategy

#### Search Strategy Summary:

This strategy was developed in conjunction with medical librarian (MS) familiar with systematic review. After discussing the plan, the databases to be searched, and the strategy, the search was performed by the librarian. The following is a summary of the search which she has prepared:

Database: Ovid MEDLINE(R) ALL <1946 to February 21, 2019> Search Strategy:
--------------------------------------------------------------------------------
1 athletes/ or exp sports/ or military personnel/ (208586)
2 (athlete$1 or athletic$1 or sport$1 or military or army or "air force" or navy).mp. (219952)
3 (baseball or basketball or boxing or boxer$1 or football or golf* or gymnastic* or hockey).mp. or ((bicycling or
cyclist* or cycling) and (athlet* or sport*)).tw. (30321)
4 ("martial arts" or "tai ji" or "tai chi" or mountaineer* or "rock climb*" or "racquet sports" or tennis or running
or jogging or runner$1 or marathoner$1 or skating or skater$1).mp. (77951)
5 ("snow sports" or skiing or skiier$1 or snowboard* or "ski jump*" or soccer or "track and field" or volleyball or
swimmer$1 or swimming or diving or diver$1).mp. (63716)
6 ("weight lift*" or wrestling or wrestler$1 or rugby).mp. (9498)
7 1 or 2 or 3 or 4 or 5 or 6 (398183)
8 exp electrocardiography/ or exp echocardiography/ (307818)
9 (electrocardiogra* or ecg or ecgs or ekg or ekgs or echocardiogra*).mp. (395672)
10 mass screening/ or medical history taking/ or physical examination/ (146275)
11 (pre-participation or preparticipation or screening or ppe).mp. (550203)
12 ((pre-season or preseason or medical or cardiac or cardiovascular or sport*) adj2 (clearance or exam* or screen*
or physical or evaluat*)).tw. (44659)
13 ((pre-participation or preparticipation) adj3 (screen* or physical* or exam* or clearance or evaluat*)).tw.
(1011)
14 8 or 9 or 10 or 11 or 12 or 13 (1019248)
15 exp Heart Diseases/di, dg, mo, pc [Diagnosis, Diagnostic Imaging, Mortality, Prevention & Control] (426749)
16 ((heart or cardiac or cardiovascular) adj3 abnormal*).tw. (15007)
17 ((heart or cardiac or cardiovascular) adj3 (lethal or fatal* or mortalit*)).tw. (44311)
18 ((heart or cardiac or cardiovascular) adj3 patholog*).tw. (9443)
19 ((heart or cardiac) adj3 (death or arrest)).tw. (60335)
20 (("sudden death" or scd or sca) not sickle).tw. (31437)
21 15 or 16 or 17 or 18 or 19 or 20 (532916)
22 7 and 14 and 21 (3729)

23 exp heart diseases/mo (79233)
24 17 or 19 or 20 or 23 (184538)
25 vital statistics/ or morbidity/ or incidence/ or prevalence/ or mortality/ or "cause of death"/ or child
mortality/ or fatal outcome/ or mortality, premature/ or survival rate/ (776559)
26 (rate or rates or statistic* or epidemiolog* or incidence or prevalence or frequency).tw. (4827926)
27 25 or 26 (5107453)
28 7 and 24 and 27 (1708)
29 22 or 28 (4770)
30 29 not (letter or news or comment or editorial).pt. (4447)

Database: Embase <1974 to 2019 February 21> Search Strategy:
--------------------------------------------------------------------------------
1 exp athlete/ or exp sport/ or soldier/ (199744)
2 (athlete$1 or athletic$1 or sport$1 or military or army or "air force" or navy).mp. (240894)
3 (baseball or basketball or boxing or boxer$1 or football or golf* or gymnastic* or hockey).mp. or ((bicycling or
cyclist* or cycling) and (athlet* or sport*)).tw. (34495)
4 ("martial arts" or "tai ji" or "tai chi" or mountaineer* or "rock climb*" or "racquet sports" or tennis or running
or jogging or runner$1 or marathoner$1 or skating or skater$1).mp. (96438)
5 ("snow sports" or skiing or skiier$1 or snowboard* or "ski jump*" or soccer or "track adj3 field" or volleyball or
swimmer$1 or swimming or diving or diver$1).mp. (68917)
6 ("weight lift*" or wrestling or wrestler$1 or rugby).mp. (10918)
7 1 or 2 or 3 or 4 or 5 or 6 (404841)
8 exp electrocardiography/ or exp echocardiography/ (428456)
9 (electrocardiogra* or ecg or ecgs or ekg or ekgs or echocardiogra*).mp. (574447)
10 mass screening/ or medical history/ or physical examination/ or medical assessment/ (371074)
11 (pre-participation or preparticipation or screening or ppe).mp. (961400)
12 ((pre-season or preseason or medical or cardiac or cardiovascular or sport*) adj2 (clearance or exam* or screen*
or physical or evaluat*)).tw. (63248)
13 ((pre-participation or preparticipation) adj3 (screen* or physical* or exam* or clearance or evaluat*)).tw.
(1419)
14 8 or 9 or 10 or 11 or 12 or 13 (1835968)
15 exp heart disease/ (1669899)
16 ((heart or cardiac or cardiovascular) adj3 abnormal*).tw. (21384)
17 ((heart or cardiac or cardiovascular) adj3 (lethal or fatal* or mortalit*)).tw. (67144)
18 ((heart or cardiac or cardiovascular) adj3 patholog*).tw. (13481)
19 ((heart or cardiac) adj3 (death or arrest)).tw. (96217)
20 (("sudden death" or scd or sca) not sickle).tw. (46305)
21 15 or 16 or 17 or 18 or 19 or 20 (1738361)
22 7 and 14 and 21 (8540)
23 limit 22 to human (7429)

24 limit 23 to (editorial or letter or note) (623)
25 23 not 24 (6806)
26 mortality/ or cardiovascular mortality/ or childhood mortality/ or exp mortality rate/ or premature mortality/
(774870)
27 vital statistics/ or morbidity/ or incidence/ or prevalence/ or "cause of death"/ or fatality/ or survival rate/
(1562652)
28 (rate or rates or statistic* or epidemiolog* or incidence or prevalence or frequency).tw. (6379022)
29 26 or 27 or 28 (7161609)
30 7 and 21 and 29 (8219)
31 limit 30 to human (6920)
32 limit 31 to (editorial or letter or note) (343)
33 31 not 32 (6577)
34 25 or 33 (10368)

Cochrane CENTRAL
Date Run: 25/02/2019 21:07:29

ID Search Hits
#1 [mh athletes] or [mh sports] or [mh "military personnel"] 15005
#2 athlete* or athletic* or sport or sports or military or army or "air force" or navy 27710
#3 baseball or basketball or boxing or boxer* or football or golf* or gymnastic* or hockey 1999
#4 (bicycling or cyclist* or cycling) and (athlet* or sport*) 2009
#5 "martial arts" or "tai ji" or "tai chi" or mountaineer* or "rock climbing" or "racquet sports" or tennis or running or runner* or jogging or jogger* or runner or marathon* or skating or skater* 7653
#6 "snow sports" or skiing or skiier* or snowboard* or "ski jump*" or soccer or "track and field" or volleyball or swimmer* or swimming or diving or divers 4794
#7 "weight lifting" or "weight lifters" or "weight lifter" or wrestling or wrestler* or rugby 1332
#8 #1 or #2 or #3 or #4 or #5 or #6 or #7 42075
#9 [mh electrocardiography] or [mh echocardiography] 12185
#10 electrocardiogra* or ecg or ecgs or ekg or ekgs or echocardiogra* 29827
#11 [mh ^"mass screening"] or [mh ^"medical history taking"] or [mh ^"physical examination"] 3857
#12 pre-participation or preparticipation or screening or ppe 34905
#13 (pre-season or preseason or medical or cardiac or cardiovascular or sport*) near/2 (clearance or exam* or screen* or physical or evaluat*) 5112
#14 (pre-participation or preparticipation) near/3 (screen* or physical* or exam* or clearance or evaluat*) 12
#15 #9 or #10 or #11 or #12 or #13 or #14 67737
#16 [mh "heart diseases"/DI,DG,MO,PC] 18825
#17 (heart or cardiac or cardiovascular) NEAR/3 abnormal* 873

#18 (heart or cardiac or cardiovascular) NEAR/3 (lethal or fatal* or mortalit*) 7124
#19 (heart or cardiac or cardiovascular) NEAR/3 patholog* 239
#20 (heart or cardiac) NEAR/3 (death or arrest) 9006
#21 ("sudden death" or scd or sca) not sickle 1859
#22 #16 OR #17 OR #18 OR #19 OR #20 OR #21 32227
#23 #8 AND #15 AND #22 371
#24 [mh "heart diseases"/mo] 6083
#25 #18 or #20 or #21 or #24 19853
#26 [mh ^"vital statistics"] or [mh ^morbidity] or [mh ^incidence] or [mh ^prevalence] or [mh ^mortality] or [mh ^"cause of death"] or [mh ^"child mortality"] or [mh ^"fatal outcome"] or [mh ^"mortality, premature"] or [mh ^"survival rate"] 23815
#27 rate or rates or statistic* or epidemiolog* or incidence or prevalence or frequency 482166
#28 #26 or #27 482567
#29 #8 and #25 and #28 506
#30 #23 or #29 in Trials 253

Web of Science Core Collection: Citation Indexes 27-2-19
# 1
1,123,261
TOPIC: (athlete* or athletic* or sport* or military or army or "air force" or navy) OR TOPIC: (baseball or basketball or boxing or boxer* or football or golf* or gymnastic* or hockey) OR TOPIC: ((bicycling or cyclist* or cycling) and (athlet* or sport*)) OR TOPIC: ("martial arts" or "tai ji" or "tai chi" or mountaineer* or "rock climb*" or "racquet sports" or tennis or running or jogging or runner* or marathoner* or skating or skater*) OR TOPIC: ("snow sports" or skiing or skiier* or snowboard* or "ski jump*" or soccer or "track and field" or volleyball or swimmer* or swimming or diving or divers) OR TOPIC: ("weight lift*" or wrestling or wrestler* or rugby)
Indexes=SCI-EXPANDED, SSCI, A&HCI, CPCI-S, CPCI-SSH, ESCI Timespan=All years

# 2
1,116,849
TS=(electrocardiogra* or ecg or ecgs or ekg or ekgs or echocardiogra*) OR TS=("pre-participation" or preparticipation or screening or ppe) OR TS=((pre-season or preseason or medical or cardiac or cardiovascular or sport*) near/2 (clearance or exam* or screen* or physical or evaluat*)) OR TS=((pre-participation or preparticipation) near/3 (screen* or physical* or exam* or clearance or evaluat*))
Indexes=SCI-EXPANDED, SSCI, A&HCI, CPCI-S, CPCI-SSH, ESCI Timespan=All years

# 3
575,492
TS=((heart or cardiac or cardiovascular) near/3 (disease* or abnormal*)) OR TS=((heart or cardiac or cardiovascular) near/3 (lethal or fatal* or mortalit*)) OR TS=((heart or cardiac or cardiovascular) near/3 patholog*) OR TS=((heart or cardiac) near/3 (death or arrest)) OR TS=(("sudden death" or scd or sca) not sickle)

Indexes=SCI-EXPANDED, SSCI, A&HCI, CPCI-S, CPCI-SSH, ESCI Timespan=All years

# 4
3,130
#3 AND #2 AND #1
Indexes=SCI-EXPANDED, SSCI, A&HCI, CPCI-S, CPCI-SSH, ESCI Timespan=All years

# 5
6,387,434
TS=(morbidity or statistic* or epidemiolog* or incidence or prevalence or frequency or sensitivity or specificity)
Indexes=SCI-EXPANDED, SSCI, A&HCI, CPCI-S, CPCI-SSH, ESCI Timespan=All years

# 6
167,344
TS=((heart or cardiac or cardiovascular) near/3 (lethal or fatal* or mortalit* or death or arrest)) OR TS=(("sudden death" or scd or sca) not sickle)
Indexes=SCI-EXPANDED, SSCI, A&HCI, CPCI-S, CPCI-SSH, ESCI Timespan=All years

# 7
1,714
#6 AND #5 AND #1
Indexes=SCI-EXPANDED, SSCI, A&HCI, CPCI-S, CPCI-SSH, ESCI Timespan=All years

# 8
4,099
#7 OR #4
Indexes=SCI-EXPANDED, SSCI, A&HCI, CPCI-S, CPCI-SSH, ESCI Timespan=All years

Biosis Citation Index 1969 to date 27-2-19
# 1
410,863
TS= (athlete* or athletic* or sport* or military or army or "air force" or navy) OR TS= (baseball or basketball or boxing or boxer* or football or golf* or gymnastic* or hockey) OR TS= ((bicycling or cyclist* or cycling) and (athlet* or sport*)) OR TS= ("martial arts" or "tai ji" or "tai chi" or mountaineer* or "rock climb*" or "racquet sports" or tennis or running or jogging or runner* or marathoner* or skating or skater*) OR TS=("snow sports" or skiing or skiier* or snowboard* or "ski jump*" or soccer or "track and field" or volleyball or swimmer* or swimming or diving or divers) OR TS= ("weight lift*" or wrestling or wrestler* or rugby)
Indexes=BCI Timespan=All years

# 2
769,756

TS=(electrocardiogra* or ecg or ecgs or ekg or ekgs or echocardiogra*) OR TS=("pre-participation" or preparticipation or screening or ppe) OR TS=((pre-season or preseason or medical or cardiac or cardiovascular or sport*) near/2 (clearance or exam* or screen* or physical or evaluat*)) OR TS=((pre-participation or preparticipation) near/3 (screen* or physical* or exam* or clearance or evaluat*))
Indexes=BCI Timespan=All years

# 3
2,310,299
TS=((heart or cardiac or cardiovascular) near/3 (disease* or abnormal*)) OR TS=((heart or cardiac or cardiovascular) near/3 (lethal or fatal* or mortalit*)) OR TS=((heart or cardiac or cardiovascular) near/3 patholog*) OR TS=((heart or cardiac) near/3 (death or arrest)) OR TS=(("sudden death" or scd or sca) not sickle)
Indexes=BCI Timespan=All years

# 4
4,794
#3 AND #2 AND #1
Indexes=BCI Timespan=All years

# 5
4,515,520
TS=(morbidity or statistic* or epidemiolog* or incidence or prevalence or frequency or sensitivity or specificity)
Indexes=BCI Timespan=All years

# 6
109,729
TS=((heart or cardiac or cardiovascular) near/3 (lethal or fatal* or mortalit* or death or arrest)) OR TS=(("sudden death" or scd or sca) not sickle)
Indexes=BCI Timespan=All years

# 7
969
#6 AND #5 AND #1
Indexes=BCI Timespan=All years

# 8
5,360
#7 OR #4
Indexes=BCI Timespan=All years

Scopus 5556 hits 27-2-19

( ( ( TITLE-ABS-KEY ( athlete* OR athletic* OR sport* OR military OR army OR "air force" OR navy ) OR TITLE-ABS-KEY ( ( bicycling OR cyclist* OR cycling ) AND ( athlet* OR sport* ) ) OR TITLE-ABS-KEY ( "martial arts" OR "tai ji" OR "tai chi" OR mountaineer* OR "rock climb*" OR "racquet sports" OR tennis OR running OR jogging OR runner* OR marathoner* OR skating OR skater* ) OR TITLE-ABS-KEY ( "snow sports" OR skiing OR skiier* OR snowboard* OR "ski jump*" OR soccer OR "track and field" OR volleyball OR swimmer* OR swimming OR diving OR divers ) OR TITLE-ABS-KEY ( "weight lift*" OR wrestling OR wrestler* OR rugby ) OR TITLE-ABS-KEY ( baseball OR basketball OR boxing OR boxer* OR football OR golf* OR gymnastic* OR hockey ) ) ) AND ( ( TITLE-ABS-KEY ( electrocardiogra* OR ecg OR ecgs OR ekg OR ekgs OR echocardiogra* ) OR TITLE-ABS-KEY ( "pre-participation" OR preparticipation OR screening OR ppe ) OR TITLE-ABS-KEY ( ( pre-season OR preseason OR medical OR cardiac OR cardiovascular OR sport* ) W/2 ( clearance OR exam* OR screen* OR physical OR evaluat* ) ) OR TITLE-ABS-KEY ( ( pre-participation OR preparticipation ) W/3 ( screen* OR physical* OR exam* OR clearance OR evaluat* ) ) ) ) AND ( ( TITLE-ABS-KEY ( ( heart OR cardiac OR cardiovascular ) W/3 ( disease* OR abnormal* OR lethal OR fatal* OR mortalit* OR patholog* ) ) OR TITLE-ABS-KEY ( ( heart OR cardiac ) W/3 ( death OR arrest ) ) OR TITLE-ABS-KEY ( ( "sudden death" OR scd OR sca ) AND NOT sickle ) ) ) ) OR ( ( ( TITLE-ABS-KEY ( athlete* OR athletic* OR sport* OR military OR army OR "air force" OR navy ) OR TITLE-ABS-KEY ( ( bicycling OR cyclist* OR cycling ) AND ( athlet* OR sport* ) ) OR TITLE-ABS-KEY ( "martial arts" OR "tai ji" OR "tai chi" OR mountaineer* OR "rock climb*" OR "racquet sports" OR tennis OR running OR jogging OR runner* OR marathoner* OR skating OR skater* ) OR TITLE-ABS-KEY ( "snow sports" OR skiing OR skiier* OR snowboard* OR "ski jump*" OR soccer OR "track and field" OR volleyball OR swimmer* OR swimming OR diving OR divers ) OR TITLE-ABS-KEY ( "weight lift*" OR wrestling OR wrestler* OR rugby ) OR TITLE-ABS-KEY ( baseball OR basketball OR boxing OR boxer* OR football OR golf* OR gymnastic* OR hockey ) ) ) AND ( TITLE-ABS-KEY ( morbidity OR statistic* OR epidemiolog* OR incidence OR prevalence OR frequency OR sensitivity OR specificity ) ) AND ( ( TITLE-ABS-KEY ( ( heart OR cardiac OR cardiovascular ) W/3 ( lethal OR fatal* OR mortalit* OR death OR arrest ) ) OR TITLE-ABS-KEY ( ( "sudden death" OR scd OR sca ) AND NOT sickle ) ) ) ) AND ( LIMIT-TO ( DOCTYPE , "ar" ) OR LIMIT-TO ( DOCTYPE , "re" ) OR LIMIT-TO ( DOCTYPE , "cp" ) OR LIMIT-TO ( DOCTYPE , "ch" ) OR LIMIT-TO ( DOCTYPE , "sh" ) OR LIMIT-TO ( DOCTYPE , "ip" ) OR LIMIT-TO ( DOCTYPE , "bk" ) OR LIMIT-TO ( DOCTYPE , "cr" ) )
Limited to article, review, conference paper, book chapter, short survey, article in press, book, or conference review
SportDiscus Friday, March 01, 2019 1:12:16 PM
S1 (DE "ATHLETES" OR DE "ABORIGINAL Australian athletes" OR DE "AFRICAN athletes" OR DE "AMATEUR athletes" OR DE "ARAB athletes" OR DE "ARCHERS" OR DE "ASIAN athletes" OR DE "ATHLETES as actors" OR DE "ATHLETES in art" OR DE "ATHLETES with disabilities" OR DE "BADMINTON players" OR DE "BASEBALL players" OR DE "BASKETBALL players" OR DE "BLACK athletes" OR DE "BOBSLEDDERS" OR DE "BODYBUILDERS" OR DE "BOWLERS" OR DE "BOXERS (Sports)" OR DE "BULLFIGHTERS" OR DE "CANADIAN athletes" OR DE "CANOEISTS" OR DE "CELEBRITY athletes" OR DE "CHILD athletes" OR DE "CHILDREN of athletes" OR DE "CHRISTIAN athletes" OR DE "COLLEGE athletes" OR DE "CRICKET players" OR DE "CROQUET players" OR DE

"CURLERS (Athletes)" OR DE "CYCLISTS" OR DE "DEFENSIVE players" OR DE "DIABETIC athletes" OR DE "DIRTBOARDERS" OR DE "ELITE athletes" OR DE "ENDURANCE athletes" OR DE "EUROPEAN athletes" OR DE "FENCERS" OR DE "FOOTBALL players" OR DE "GAY athletes" OR DE "GLADIATORS" OR DE "GOLFERS" OR DE "GYMNASTS" OR DE "HANDBALL players" OR DE "HIGH school athletes" OR DE "HOCKEY players" OR DE "INTERSEX athletes" OR DE "JAI alai players" OR DE "JEWISH athletes" OR DE "JUNIOR high school athletes" OR DE "KABADDI players" OR DE "LACROSSE players" OR DE "LAWN bowlers" OR DE "LGBT athletes" OR DE "LONG-term athlete development" OR DE "MALE athletes" OR DE "MARTIAL artists" OR DE "MEXICAN athletes" OR DE "MIDDLE school athletes" OR DE "MOUNTAINEERS" OR DE "MUSLIM athletes" OR DE "NATIVE American athletes" OR DE "NETBALL players" OR DE "OFFENSIVE players" OR DE "OLDER athletes" OR DE "OLYMPIC athletes" OR DE "ORIENTEERS" OR DE "PACIFIC Islander athletes" OR DE "PROFESSIONAL athletes" OR DE "ROWERS" OR DE "RUGBY football players" OR DE "RUNNERS (Sports)" OR DE "SKATERS" OR DE "SKIERS" OR DE "SKYDIVERS" OR DE "SNOWBOARDERS" OR DE "SOCCER players" OR DE "SOFTBALL players" OR DE "SQUASH players" OR DE "STARTING players" OR DE "SUBSTITUTE players" OR DE "SURFERS" OR DE "SWIMMERS" OR DE "TABLE tennis players" OR DE "TEAM handball players" OR DE "TENNIS players" OR DE "TRACK & field athletes" OR DE "TRIATHLETES" OR DE "VOLLEYBALL players" OR DE "WATER polo players" OR DE "WEIGHT lifters" OR DE "WINDSURFERS (Persons)" OR DE "WOMEN athletes" OR DE "WRESTLERS") OR (DE "ATHLETICS" OR DE "AMATEUR sports" OR DE "ATHLETIC tryouts" OR DE "BAG punching" OR DE "BOXING" OR DE "COLLEGE sports" OR DE "DUATHLON" OR DE "FENCING" OR DE "GOODWILL Games" OR DE "GYMNASTICS" OR DE "HIGHLAND games" OR DE "JIU-jitsu" OR DE "MIXED martial arts" OR DE "PANCRATIUM" OR DE "PARKOUR" OR DE "POWERLIFTING" OR DE "PROFESSIONALISM in sports" OR DE "SENIOR Olympics" OR DE "SKATING" OR DE "SWIMMING" OR DE "TETRATHLON" OR DE "TRACK & field" OR DE "TRIATHLON" OR DE "WALKING" OR DE "WEIGHT lifting" OR DE "WRESTLING")

S2 KW ( Athlete or athletes or athletic or athletics ) OR TI ( Athlete or athletes or athletic or athletics ) OR AB ( Athlete or athletes or athletic or athletics ) Search modes - Boolean/Phrase Interface - EBSCOhost Research Databases

S3 DE "MILITARY sports" OR TX military Search modes - Boolean/Phrase Interface - EBSCOhost Research Databases

S4 DE "PERIODIC health examinations" Search modes - Boolean/Phrase Interface - EBSCOhost Research Databases

S5 ( (DE "ELECTROCARDIOGRAPHY" OR DE "VECTORCARDIOGRAPHY") OR (DE "ECHOCARDIOGRAPHY" OR DE "STRESS echocardiography") ) OR ( electrocardiogra* or ecg or ecgs or ekg or ekgs or echocardiogra* ) Search modes - Boolean/Phrase Interface - EBSCOhost Research Databases

S6 DE "MEDICAL screening" OR ( pre-participation or preparticipation or screening or ppe ) Search modes - Boolean/Phrase Interface - EBSCOhost Research Databases

S7 ( (pre-season or preseason or medical or cardiac or cardiovascular or sport*) W2 (clearance or exam* or screen* or physical or evaluat*) ) OR ( (pre-participation or preparticipation) w3 (screen* or physical* or exam* or clearance or evaluat*) ) Search modes - Boolean/Phrase Interface - EBSCOhost Research Databases

S8 DE "HEART diseases" OR DE "ARRHYTHMIA" OR DE "CARDIAC arrest" OR DE "CARDIAC hypertrophy" OR DE "CARDIOMYOPATHIES" OR DE "CORONARY heart disease" OR DE "HEART block" OR DE "HEART dilatation" OR DE "HEART failure" Search modes - Boolean/Phrase Interface - EBSCOhost Research Databases
S9 DE "SUDDEN death" Search modes - Boolean/Phrase Interface - EBSCOhost Research Databases
S10 (heart or cardiac or cardiovascular) w3 abnormal* Search modes - Boolean/Phrase Interface - EBSCOhost Research Databases
S11 (heart or cardiac or cardiovascular) w3 (lethal or fatal* or mortalit*) Search modes - Boolean/Phrase Interface - EBSCOhost Research Databases
S12 (heart or cardiac or cardiovascular) w3 patholog* Search modes - Boolean/Phrase Interface - EBSCOhost Research Databases
S13 (heart or cardiac) w3 (death or arrest) Search modes - Boolean/Phrase Interface - EBSCOhost Research Databases
S14 ( ("sudden death" or scd or sca) ) NOT sickle Search modes - Boolean/Phrase Interface - EBSCOhost Research Databases
S15 S1 OR S2 OR S3 Search modes - Boolean/Phrase Interface - EBSCOhost Research Databases
S16 ( s1 or s2 or s3 ) AND ( s4 or s5 or s6 or s7 ) AND ( s8 or s9 or s10 or s11 or s12 or s13 or s14 ) Search modes - Boolean/Phrase Interface - EBSCOhost Research Databases
S17 S9 OR S11 OR S13 OR S14 Search modes - Boolean/Phrase Interface - EBSCOhost Research Databases
S18 (DE "DEATH rate") AND (DE "MORTALITY" OR DE "ATHLETE mortality") Search modes - Boolean/Phrase Interface - EBSCOhost Research Databases
S19 morbidity or statistic* or epidemiolog* or incidence or prevalence or frequency or sensitivity or specificity Search modes - Boolean/Phrase Interface - EBSCOhost Research Databases
S20 s15 AND s17 AND ( s18 or s19 ) Search modes - Boolean/Phrase Interface - EBSCOhost Research Databases
S21 S16 OR S20 Search modes - Boolean/Phrase Interface - EBSCOhost Research Databases

PEDRO 1/3/19 -- no hits
Sudden AND death and sport*
OR
Sudden AND death AND athlet*

Clinicaltrials.gov July 29, 2019 -57 hits
Sudden cardiac death
AND
ECG
Screening
Athletes
Military

### APPENDIX C: Custom Risk of Bias Tool for Prevalence studies

- EPIDEMIOLOGY INTERNAL VALIDITY: CASE DEFINITION
  - LOW RISK: Acceptable definition was used.
    - When the study identifies sudden cardiac death, and considers and rules out other possibilities, such as sickle cell, or exertion heat illness. The answer is: Yes (LOW RISK).
  - HIGH RISK: An acceptable case definition was NOT used
    - For a study on sudden cardiac death, and all sudden exertion deaths are included without considering other possibilities. The answer is: No (HIGH RISK).
  - Unclear (HIGH RISK)
    - The definition is not detailed, or explained and only reported as sudden cardiac death. Answer is unclear (High risk)
- EPIDEMIOLOGY INTERNAL VALIDITY: NUMERATOR AND DENOMINATOR
  - LOW RISK: The paper presented appropriate numerator(s) AND denominator(s) for the number of subjects monitored, and the number suffering SCA/SCD
  - HIGH RISK: The paper did present numerator(s) AND denominator(s) for the parameter of interest but one or more of these were inappropriate or it is not clear where these numbers come from.
  - Unclear (HIGH RISK)
- EPIDEMIOLOGY INTERNAL VALIDITY: STUDY PERIOD
  - Was the length of shortest prevalence period for parameter of interest appropriate? (Study contains at least 100,000 athlete years)
  - LOW RISK: The shortest prevalence period for the parameter of interest was appropriate (study contains at least 100,000 athlete/military years), Yes (LOW RISK).
  - HIGH RISK: The shortest prevalence period for the parameter of interest was not appropriate (ex: 3,000 athletes monitored for 3 years), the answer is No (HIGH RISK).
  - Unclear (HIGH RISK), the total person years included in the study is not reported.
- EPIDEMIOLOGY EXT VALIDITY: NONRESPONSE BIAS/FOLLOW UP OF COHORT (Survey only)
  - LOW RISK: Was likelihood of nonresponse bias minimal?
    - The response rate for the study was >/=75%, OR, an analysis was performed that showed no significant difference in relevant demographic characteristics between responders and non- responders
    - The response rate was 68%; however, the researchers did an analysis and found no significant difference between responders and non-responders in terms of age, sex, occupation and socio- economic status. The answer is: Yes (LOW RISK).
  - HIGH RISK: Was likelihood of nonresponse bias high?

        - The response rate was <75%, and if any analysis comparing responders and non-responders was done, it showed a significant difference in relevant demographic characteristics between responders and non-responders.
        - The response rate was 65% and the researchers did NOT carry out an analysis to compare relevant demographic characteristics between responders and non-responders. The answer is: No (HIGH RISK).
        - The response rate was 69% and the researchers did an analysis and found a significant difference in age, sex and socio-economic status between responders and non-responders. The answer is: No (HIGH RISK).
    - Unclear (High Risk): Response rate not detailed.
- EPIDEMIOLOGY EXT VALIDITY: REPRESENTATIVE SAMPLE
    - The target population refers to the group of people or entities to which the results of the study will be generalized.
    - LOW RISK: Was the study population a close representation of intended/target study population in relation to relevant variables?
        - Yes (LOW RISK): The study's target population was a close representation of the national population.
        - Examples:  The study was a national health survey of people 15 years and over and the sample was drawn from a list that included all individuals in the population aged 15 years and over. The answer is: Yes (LOW RISK).
    - HIGH RISK: No (HIGH RISK): Was the study population a close representation of intended/target study population in relation to relevant variables?
        - The study's target population was clearly NOT representative of the national population.
        - The study was undertaken in one village only and it is clear this was not representative of the national population. The answer is: No (HIGH RISK).
    - Unclear (High Risk): No description, or poor description of generalizability
        - The study was conducted in one province only, and it is not clear if this was representative of the national population. The answer is: No (HIGH RISK).
- EPIDEMIOLOGY INTERNAL VALIDITY: DATA COLLECTION SOURCE
    - LOW RISK:
        - All data were collected directly from reliable sources (medical records, autopsy reports)? The answer is: Yes (LOW RISK)
    - HIGH RISK:
        - In some instances, data were collected from a proxy (media report, web reports, insurance reports)? The answer is: No (HIGH RISK)
    - Unclear (High Risk)
        - The data source is not described, the answer is unclear (HIGH RISK).
- EPIDEMIOLOGY INTERNAL VALIDITY: MODE OF DATA COLLECTION
    - The mode of data collection is the method used for collecting information from the subjects. In this case, this could be media/web reports; insurance claims on

hospital stays or life insurance; medical or autopsy records; Reporting databases such as national sudden death registry, NCAA injury reporting, military records.

  - LOW RISK
    - The same mode of data collection was used for all subjects. The answer is Yes: (LOW RISK).
    - All eligible subjects had records recovered from NCAA/Military database. The answer is: Yes (LOW RISK).
  - HIGH RISK
    - The same mode of data collection was NOT used for all subjects.
    - Some subjects had databased information, while others were strictly web reports/media reports with no corroboration. The answer is: No (HIGH RISK).
  - Unclear (High Risk)
- EPIDEMIOLOGY EXT VALIDITY: SAMPLING/SUBJECTS INCLUDED
  - The sampling frame is a list of the sampling units in the target population and the study sample is drawn from this list.
  - LOW RISK:
    - Was the sampling frame a true or close representation of the target population? Yes (low risk)
    - The sampling frame was a list of almost every individual within the target population. The answer is: Yes (LOW RISK).
    - The cluster sampling method was used and the sample of clusters/villages was drawn from a list of all villages in the target population. The answer is: Yes (LOW RISK).
  - HIGH RISK:
    - The sampling frame was a list of just one particular ethnic group/sex (i.e. having 95% white athletes in US when studying all athletes) within the overall target population, which comprised many groups. The answer is: No (HIGH RISK).
  - Unclear (High Risk)/N/A
- EPIDEMIOLOGY FINAL SUMMARY:
  - LOW RISK OF BIAS: Further research is very unlikely to change our confidence in the estimate.
  - MODERATE RISK OF BIAS: Further research is likely to have an important impact on our confidence in the estimate and may change the estimate.
  - HIGH RISK OF BIAS: Further research is very likely to have an important impact on our confidence in the estimate and is likely to change the estimate.

#### Appendix D: Deviations from original protocol

There were several deviations from the original protocol which must be explained. In several cases, articles which did not explicitly explain the ages included were selected for inclusion. This was done when the articles appeared overwhelmingly to include subjects in the eligible age range. One study was selected for inclusion which included a cohort of American firefighters, which not in our pre-specified inclusion criteria, but was elected for inclusion due to the similarity in their training and age profile to military members. When screening articles, it was discovered that in two cases, cross sectional surveys reported incidence rates based on the results of their surveys. These were not originally eligible for inclusion, but were included based on the reporting of incidence rates. The search strategy varied slightly from the original protocol in two ways: 1) Due to a mis-communication with the primary author, and the medical librarian, Clinicaltrials.gov was not originally searched with other databases and the search occurred late in the process, and the results were screened only by the primary author; 2) The references of included studies were not searched as originally planned. Due to the volume of the studies included in the original search, the search was felt to be adequate, and the plan to do so was dropped.

Peterson, et. al. (2020) was included in the study outside of the original search. We elected to include this study as we were aware of the database during the writing of the original dissertation that preceded this paper. The senior author declined to share the data ahead of publication. As we were aware of the data ahead of completion of this systematic review, and had intended to include it, we elected to include the published paper.

The original plan for ROB determination in observational studies was to be done with the Newcastle-Ottawa scale [5]. It was decided to use a validated tool to evaluate risk of bias in prevalence studies [6] which more effectively assessed the cohort studies included which reported on incidence data. The data extraction, and ROB determination was originally intended to be double reviewed by the primary author, and co-reviewer. This was modified for timing reasons, and availability of second reviewers. Two more second reviewers were added for screening, review, and extraction; and only a portion (51%) of the included articles were double reviewed and extracted.

Primary authors were contacted during the screening process for questions regarding eligibility for inclusion; however, authors were not contacted based on risk of bias questions as indicated in the original protocol.

The original protocol described an expectation of visual inspection of funnel plots when pooling more than ten studies. This was done only with the two primary analyses, and not subgroup or sensitivity analysis. This decision was made keeping in mind the difficulty of assessing publication bias in subgroups extracted from fully reported datasets.

Lastly, it was determined that GRADE summary of findings table would not be used to describe the incidence portion of the review, based on the observational nature, and the lack of a test of an intervention.

### Appendix E: Characteristics of Included Studies

#### *Assanelli 1995*

| | |
|---|---|
| **Methods** | **Study design:** Prospective cohort study<br>**Prospective cohort:** Poorly described but appears prospective for 1 year on cohort of athletes screened by ECG. Describes the SCA/SCD events occurring in this cohort. |
| **Participants** | **Age:** Unclear. Deaths reported in ages 17-36, and includes amateur athletes<br>**Sport:** combination<br>**Level of Sport:** combination<br>**Gender:** both<br>**ECG Screened:** yes |
| **Intervention** | Observational |
| **Outcomes** | **SCD:** Yes<br>**SCA:** Yes<br>**Exertional:** not detailed<br>**Method of death ID:** Medical Records |
| **Identification** | **Sponsorship source:** None described<br>**Country:** Italy<br>**Setting:** Brescia Province<br>**Comments:** Community athletes, reported as low level amateurs<br>**Authors name:** Dr. Deodata Assanelli<br>**Institution:** Cattedra di Cardiologia<br>**Email:** none listed<br>**Address:** Dr. Deodato Assanelli Cattedra di Cardiologia University degli Studi Piazzale Spedali Civili1-25100 Brescia, Italia |
| **Notes** | Italian language, needed translation |

Risk of bias table

| **Bias** | **Authors' judgement** | **Support for judgement** |
|---|---|---|
| EPIDEMIOLOGY EXT VALIDITY: REPRESENTATIVE SAMPLE | Unclear risk | Judgement Comment: Appear to include all athletes in Brescia province, but again not well described. |
| EPIDEMIOLOGY INTERNAL VALIDITY: CASE DEFINITION | Unclear risk | Judgement Comment: No case definition given, only described the 7 cases reported in this paper, did not describe differences between how one was considered a case vs. not a case |
| EPIDEMIOLOGY EXT VALIDITY: | High risk | Judgement Comment: Not described |

| | | |
|---|---|---|
| SAMPLING/SUBJECTS INCLUDED | | |
| EPIDEMIOLOGY INTERNAL VALIDITY: STUDY PERIOD | High risk | Judgement Comment: only 16000 athlete years |
| EPIDEMIOLOGY INTERNAL VALIDITY: DATA COLLECTION SOURCE | Unclear risk | Judgement Comment: Appears to be data from medical records and autopsy records. But again, not described. |
| EPIDEMIOLOGY INTERNAL VALIDITY: NUMERATOR AND DENOMINATOR | High risk | Judgement Comment: No explanation of how the cases, and the denominator were arrived at. Is only a case series with a denominator included in the article. |
| EPIDEMIOLOGY INTERNAL VALIDITY: MODE OF DATA COLLECTION | Unclear risk | Judgement Comment: Not described. |
| EPIDEMIOLOGY FINAL SUMMARY: | High risk | Judgement Comment: Very high risk of bias. Poorly described methodology for how the authors came to the numbers in either numerator/denominator. No info on ID of the cases. The authors do provide these numbers-but we will take this with little confidence. |
| EPIDEMIOLOGY EXT VALIDITY: NONRESPONSE BIAS/FOLLOW UP OF COHORT | N/A | |

#### *Berge 2018*

| | |
|---|---|
| **Methods** | **Study design:** Retrospective cohort study<br>**Retrospective cohort:** male football players from 28/30 Norwegian professional teams btwn 2008-2016. 595 of 604 players consented to be included |
| **Participants** | **Included criteria:** Male professional football players in Norway. Had to consent to be included<br>**Excluded criteria:** Players did not consent to be included<br>**Age:** 18-40<br>**Gender:** Male<br>**Sport:** Soccer<br>**Level of Sport:** Professional<br>**ECG Screened:** Yes |
| **Intervention** | Observational |

| | |
|---|---|
| **Outcomes** | **SCA**: Yes<br>**SCD:** (none occurred)<br>**Exertional:** all occurrences<br>**Method of ID cases**: Media occurrences/web reports to identify incidents, medical records to confirm |
| **Identification** | **Sponsorship source:** The research reported in this article was supported by the Oslo Sports Trauma Research Center and the Norwegian Olympic Sports Center, Oslo, Norway<br>**Country:** Norway<br>**Setting:** Norwegian Professional Men's football<br>**Comments:** NA<br>**Authors name:** Hilde Moseby Berge<br>**Institution:** Oslo Sports Trauma Research Center<br>**Email:**<br>**Address:** Oslo Sports Trauma Research Center, Department of Sports Medicine, Norwegian School of Sport Sciences, Oslo 0806, Norway |

Risk of bias table

| **Bias** | **Authors' judgement** | **Support for judgement** |
|---|---|---|
| EPIDEMIOLOGY EXT VALIDITY: REPRESENTATIVE SAMPLE | Low risk | Author Judgement: 595 of 604 players consented to be included |
| EPIDEMIOLOGY INTERNAL VALIDITY: CASE DEFINITION | Low risk | Quote: "A CV incident was defined as any diagnosis of CVD, SCA or SCD recorded in the player's medical record." |
| EPIDEMIOLOGY EXT VALIDITY: SAMPLING/SUBJECTS INCLUDED | Low risk | Judgement Comment: Nearly all players approached, consented for project |
| EPIDEMIOLOGY INTERNAL VALIDITY: STUDY PERIOD | High risk | Less than 5,000 person years |
| EPIDEMIOLOGY INTERNAL VALIDITY: DATA COLLECTION SOURCE | High risk | Judgement Comment: ID of events occurred with web report |
| EPIDEMIOLOGY INTERNAL VALIDITY: NUMERATOR AND DENOMINATOR | Low risk | Quote: "We followed a cohort with an exact denominator, making the incidence estimate more reliable. “ |
| EPIDEMIOLOGY INTERNAL VALIDITY: MODE OF DATA COLLECTION | Low risk | Quote: "For the follow-up study we used Retriever (Tekinetics), soft- ware that retrieves desired content from the worldwide web, to identify |

| | | |
|---|---|---|
| | | football players from the cohort who had suffered CVD, SCA or SCD after the screening. We searched for CV incidents between 1 February 2008 and 30 April 2016 using the search terms (in Norwegian) 'football' AND 'SCD, SCA, angina pectoris, myocardial infarction, transient ischemic attack, stroke, cerebral hemorrhage or infarction, atrial flutter or fibrillation, septum, blood clots, myocarditis or pericarditis'. We excluded search terms related to hypertension since we thought such cases were less likely to be covered by the media. In addition, detailed results on the prevalence of hypertension in this cohort have been published already. A medical student (SB) checked the results manually and we included all reports on players in the original cohort." |
| EPIDEMIOLOGY FINAL SUMMARY: | Moderate Risk | Media source for identification of deaths presents some bias for missed cases, and the small number of person years allows for a possible random variation to suggest higher rate than one might otherwise see; however, the article is transparent, and appears well done otherwise. |
| EPIDEMIOLOGY EXT VALIDITY: NONRESPONSE BIAS/FOLLOW UP OF COHORT | N/A | |

#### *Biasco 2013*

| | |
|---|---|
| **Methods** | **Study design:** Retrospective cohort study<br>**Study grouping:** NA<br>**Cross sectional:** NA<br>**Prospective cohort:** NA<br>**Retrospective cohort:** Athletes screened, and followed by unclear method. Authors report 13.3 median years of follow up (10-17 interquartile range). Primary goal of the article was to detail J point elevation in soccer players, but followed up the included cohort, and reported events of SCA/D |
| **Participants** | **Included criteria:** Male professional soccer players screened at Turin Sports Medicine Institute |

| | |
|---|---|
| | **Excluded criteria:** None listed<br>**Age:** avg 23.6, (SD 5.3) No age range given, only the mean and SD<br>**Gender:** Male<br>**Sport:** Soccer<br>**Level of Sport:** Professional<br>**ECG Screened:** Yes |
| **Intervention** | Observational |
| **Outcomes** | **SCA:** Not described<br>**SCD:** Yes (No events occurred)<br>**Exertional: N**ot described<br>**Method of ID Cases:** Not described |
| **Identification** | **Sponsorship source:** none reported<br>**Country:** Italy<br>**Setting:** Turin institute of Sports Medicine Professional Italian Football players<br>**Comments:** NA<br>**Authors name:** Luigi Biasco, MD (1st): Fiorenzo Gaita, MD (last)<br>**Institution:** Division of Cardiology, Department of Medical Science, Città della Salute e della Scienza University of Torino, Turin, Italy<br>**Email:**<br>**Address:** Division of Cardiology, Department of Medical Science, University of Torino, C.so Bramante 88, 10126 Torino, Italy |
| **Notes** | *Aaron Lear* on 17/06/2019 06:05<br>**Included**<br>0 cardiovascular deaths in 332 soccer players over 13.3 years avg of follow up (4415.6 athlete years) |

Risk of bias table

| **Bias** | **Authors' judgement** | **Support for judgement** |
|---|---|---|
| EPIDEMIOLOGY EXT VALIDITY: REPRESENTATIVE SAMPLE | Low risk | Judgement Comment: Professional soccer players |
| EPIDEMIOLOGY INTERNAL VALIDITY: CASE DEFINITION | Unclear risk | Judgement Comment: No cases, so no risk of bias- but no description of their follow up either, or how one would have identified this. |
| EPIDEMIOLOGY EXT VALIDITY: SAMPLING/SUBJECTS INCLUDED | Low risk | Quote: "338 male professional elite athletes, members of soccer clubs participating in the Italian national soccer championships, who underwent the first preparticipation screening between June 1980 and April 2008 at the Turin Institute of Sport |

| | | |
|---|---|---|
| | | Medicine were screened and retrospectively evaluated." |
| EPIDEMIOLOGY INTERNAL VALIDITY: STUDY PERIOD | High risk | Judgement Comment: 338 athletes over median of 13.3 years of follow up. 338.13.3=4495.4 |
| EPIDEMIOLOGY INTERNAL VALIDITY: DATA COLLECTION SOURCE | Unclear risk | Judgement Comment: Not detailed |
| EPIDEMIOLOGY INTERNAL VALIDITY: NUMERATOR AND DENOMINATOR | High risk | Quote: " Follow-Up: Follow-up data were available for the entire cohort of athletes. During follow-up (median, 13.3 years; first and third quartiles, 10.1–17.0 years), 2 deaths, because of non-cardiovascular"<br>Judgement Comment: Denominator will be calculated by us, using median data with Interquartile range. No exact data provided by article. |
| EPIDEMIOLOGY INTERNAL VALIDITY: MODE OF DATA COLLECTION | Unclear risk | Judgement Comment: Not described apart from saying follow up 'available' |
| EPIDEMIOLOGY FINAL SUMMARY: | High risk | Judgement Comment: Article designed for another objective, but the authors report follow up on all athletes. Accurate number of athlete years in doubt, no description of where or how follow up occurred. |
| EPIDEMIOLOGY EXT VALIDITY: NONRESPONSE BIAS/FOLLOW UP OF COHORT | N/A | |

#### *Boden 2013*

| | |
|---|---|
| **Methods** | **Study design:** Prospective cohort study<br>**Prospective cohort:** Data collected prospectively |
| **Participants** | **Included criteria:** HS or College football players; Cases are voluntarily reported to National Center for Catastrophic Sports Injury Research between 1990-2010<br>**Excluded criteria:** None<br>**Age:** 14-25<br>**Gender:** male<br>**Sport:** American football |

| | |
|---|---|
| | **Level of Sport:** combination<br>**ECG Screened:** No |
| **Interventions** | Observational |
| **Outcomes** | **SCA:** No<br>**SCD:** Yes<br>**Exertional:** Not described<br>**Method of ID Cases:** Combination of witness accounts and records (from school, university, medical) if available |
| **Identification** | **Sponsorship source:** none reported<br>**Country:** USA<br>**Setting:** US high school and college football<br>**Comments:** NA<br>**Authors name:** Barry P. Boden MD<br>**Institution:** Department of Physical Education, Exercise and Sport Science, University of North Carolina at Chapel Hill, Chapel Hill, North Carolina<br>**Email:**<br>**Address:** The Orthopaedic Center, 9420 Key West Avenue, #300, Rockville, MD 20850 |
| **Notes** | |

Risk of bias table

| **Bias** | **Authors' judgement** | **Support for judgement** |
|---|---|---|
| EPIDEMIOLOGY EXT VALIDITY: REPRESENTATIVE SAMPLE | Low risk | Quote: “the Center collected data on high school and college football fatalities in the United States.” |
| EPIDEMIOLOGY INTERNAL VALIDITY: CASE DEFINITION | Unclear risk | Judgement Comment: Much data appears to be collected from witnesses, or ATCs associated with cases. It is not clear that many of the cases have autopsy, or medical data included. 63 of100 reported cardiac deaths did not have 'precise diagnosis'. |
| EPIDEMIOLOGY EXT VALIDITY: SAMPLING/SUBJECTS INCLUDED | Low risk | Judgement Comment: Entirety of national us high school and college football athletes. Some sampling may be overlooked in high school population since not all are NFHSA schools but this would be a small number |
| EPIDEMIOLOGY INTERNAL VALIDITY: STUDY PERIOD | Low risk | > 100,000 person years |

| | | |
|---|---|---|
| | | Judgement Comment: 20 year period 1990 to 2110, did compare decades, over all long surveillance period, one country (us) |
| EPIDEMIOLOGY INTERNAL VALIDITY: DATA COLLECTION SOURCE | Low risk | Judgement Comment: Data from same sources. National database surveying all high school and college programs. No medical records appear to have been reviewed, nor autopsy data. Appears only from reports to the agency by school or ATCs. |
| EPIDEMIOLOGY INTERNAL VALIDITY: NUMERATOR AND DENOMINATOR | High risk | Quote: "From July 1990 through June 2010, the Center collected data on high school and college football fatalities in the United States. It annually contacted and requested reports of any catastrophic events in organized school-sponsored sports from high school and college coaches, athletic directors, and athletic trainers; executive offices of state and national athletic organizations; and a national newspaper clipping service. Once information was received concerning a possible catastrophic football injury, the Center contacted the college athletic trainers and the National Federation of State High School Associations (NFSHSA) contacted the high school coaches or athletic trainers to obtain preliminary data (eg, the athlete's name, date of injury, age, diagnosis, school, circumstances such as practice session versus game, and mechanism of injury)." Judgement Comment: Denominator based on fairly standard data from governing agencies for football players. The numerator appears only to be included if the death was reported the Center collecting the information |
| EPIDEMIOLOGY INTERNAL VALIDITY: MODE OF DATA COLLECTION | High risk | Judgement Comment: Collected by national center for catastrophic sports injury only if information was reported to them via annual survey. Additional information from internet searches and eyewitnesses. Would suggest an under-representation of the true number. |
| EPIDEMIOLOGY FINAL SUMMARY: | High risk | Judgement Comment: I expect that we underestimate the true number of cardiac deaths with this data. There is also no real description about how the deaths are categorized as cardiac in the paper. They do provide detailed diagnosis |

| | | |
|---|---|---|
| | | information about the deaths, but it is unclear where this information comes from. |
| EPIDEMIOLOGY EXT VALIDITY: NONRESPONSE BIAS/FOLLOW UP OF COHORT | N/A | |

#### *Chevalier 2009*

| | |
|---|---|
| **Methods** | **Study design:** Prospective cohort study<br>**Prospective cohort:** Data collected on Sudden death and cardiac events that are sport related in 4 hospitals/departments in France. Total population included. |
| **Participants** | **Included criteria:** Total population of athletes 10-80 years old. Sport related cardiovascular events within 1 hour of exercise. We extracted only <35 yo.<br>**Excluded criteria:** None listed.<br>**Age:** <35<br>**Gender:** both<br>**Sport:** combination<br>**Level of Sport:** combination<br>**ECG Screened:** no |
| **Outcomes** | **SCA:** no<br>**SCD:** yes<br>**Exertional:** yes, within 1 hour of sport<br>**Method of ID cases: c**ases presenting to enrolled hospitals |
| **Identification** | **Sponsorship source:** none<br>**Country:** France<br>**Setting:** 4 departments in Southern France<br>**Comments:** Records collected from 4 hospitals. I assume this to mean the only places the participants may turn up with cardiac or sudden death events, but this is not listed in the article.<br>**Authors name:** Laurent Chevalier (1st); Francois Carre (last)<br>**Institution:** Unite ́de Biologie et Medecine du Sport, Hospital Pontchaillou,<br>**Email:**<br>**Address:** Unite ́de Biologie et Me ́decine du Sport, Hospital Pontchaillou, Rue Henri le Guillou, Rennes 35033, France |
| **Notes** | We extracted those data belonging only to the <35 patients, and this includes information from bar chart, as it was not otherwise reported. |

Risk of bias table

| Bias | Authors' judgement | Support for judgement |
| --- | --- | --- |
| EPIDEMIOLOGY EXT VALIDITY: REPRESENTATIVE SAMPLE | Low risk | Judgement Comment: appears to be representative of population, but again is not clear this is athletes, or how it was determined that all subjects were captured |
| EPIDEMIOLOGY INTERNAL VALIDITY: CASE DEFINITION | Unclear risk | Quote: "All cases of sports-related ACVEs that occurred during and/or within 1-h post-sports practice and reported to the four emergency departments have been collected. In each individual case, the same check list has been filled in by the emergency staff. In case of hospitalization, all patients' data have been collected. An MI diagnostic was carried out in accordance with ECG and classical blood markers alterations. A diagnosis of arrhythmia was performed, with the first ECG recording obtained by the emergency team on the field."<br>Judgement Comment: Not clear how SCD cases were determined |
| EPIDEMIOLOGY EXT VALIDITY: SAMPLING/SUBJECTS INCLUDED | Unclear risk | Judgement Comment: Not clear if all subjects are included |
| EPIDEMIOLOGY INTERNAL VALIDITY: STUDY PERIOD | Low risk | > 100,00 person years |
| EPIDEMIOLOGY INTERNAL VALIDITY: DATA COLLECTION SOURCE | Low risk | Judgement Comment: Hospital records all cases |
| EPIDEMIOLOGY INTERNAL VALIDITY: NUMERATOR AND DENOMINATOR | High risk | Quote: "We have compared three population groups, below 35 years of age (n = 732 747; 363 493 women), between 35 and 59 years of age (n = 812 846; 415 519 women) and above 59 years of age (n = 408 789; 224 365 women)"<br>Quote: "This 1-year prospective study carried out between 1 March 2005 and 28 February 2006 concerned three departments of the southwest of France. Four hospital medical emergency departments participated in the study."<br>Judgement Comment: Unclear if the denominator is athletes, vs. general population number. I believe athletes bc of the disparity of male vs. female |

| | | |
|---|---|---|
| | | numbers; and the reference in discussion to comparison to the other studies and incidence rates. Numerator is unclear if it includes all hospitals vs only the 4 in the study. Not clear how this works with the catchment area |
| EPIDEMIOLOGY INTERNAL VALIDITY: MODE OF DATA COLLECTION | Low risk | Judgement Comment: All data collected from hospital records |
| EPIDEMIOLOGY FINAL SUMMARY: | High risk | Judgement Comment: There is uncertainty regarding the numbers put forward in this study. The method for estimating the denominator, for capturing and determining cases does not appear in the article-it is not clear that some cases may be missed by not turning up at one of 4 included hospitals. The methodology is otherwise laid out. |
| EPIDEMIOLOGY EXT VALIDITY: NONRESPONSE BIAS/FOLLOW UP OF COHORT | N/A | |

#### *Corrado 2006*

| | |
|---|---|
| **Methods** | **Study design:** Historically controlled trial comparing athletes with ECG screening, to athletes without ECG screening; Prospectively collected incidence trial<br>**Study grouping:** Athletes dying in the pre-screening era 1979-81 compared to athletes dying in post-screening era 1982-2004<br>**Prospective cohort:** Portion extracted as prospective cohort study as well. |
| **Participants** | **Baseline Characteristics:** None Provided<br>**Included criteria:** Competitive athletes in the Veneto region of Italy from 1979-2004<br>**Excluded criteria:** None listed<br>**Pretreatment:** Not detailed.<br>**Age:** 12-35<br>**Gender:** combination<br>**Sport:** combination<br>**Level of Sport:** competitive<br>**ECG Screened:** majority of athletes screened |
| **Interventions** | **Intervention Characteristics**<br>PPE with no ECG or no PPE vs. |

| | |
|---|---|
| | PPE with ECG |
| **Outcomes** | **SCA:** no<br>**SCD:** yes<br>**Exertional:** all deaths included<br>**Method of ID cases:** combination medical record, autopsy |
| **Identification** | **Sponsorship source:** Veneto Region, Cardiocerebrovascular Pathology Registry, Venice, Italy; the European Commission research contract QLG1 CT-2000 01091; the Ministry of Health, Rome, Italy; and Fondazione Cassa di Risparmio di Padova e Rovigo, Padova, Italy<br>**Country:** Italy<br>**Setting:** Veneto Region Competitive athletes<br>**Comments:** NA<br>**Authors name:** Domenico Corrado, MD (1st); Gaetano Thiene, MD (last)<br>**Institution:** Department of Cardiac, Thoracic, and Vascular Sciences University of Padua Medical School Padua, Italy<br>**Email:**<br>**Address:** Istituto di Anatomia Patologica, Via A. Gabelli, 61- 35121 Padova, Italy |
| **Notes** | **Denominator determination:** Denominator was not detailed year by year, but rather was given over the entirety of the article. Our numbers in each group are the result of dividing this overall number by the 26 year study period.<br>Calculated from the table 1 in the paper. This is a different total number (intervention and control) compared to the number they report (Reported 2938730, calculated from the 3 separate incidence reportings: 2947482)<br>• Used total athlete years 2838730/26 yrs=113028.1/yr<br> ◦ pre-screening group= 14 deaths/339070 no deaths<br> ◦ Post-screening group=41 deaths/2599605 no deaths |

Risk of bias table

| Bias | Authors' judgement | Support for judgement |
|---|---|---|
| Random sequence generation (selection bias) | High risk | Judgement Comment: No clear description of the demographics of the groups pre-screening vs. post screening. Unable to compare the groups being compared in this trial. |
| Allocation concealment (selection bias) | Unclear risk | Judgement Comment: N/A |

| | | |
|---|---|---|
| Blinding of participants and personnel (performance bias) | Unclear risk | Judgement Comment: Not reported |
| Blinding of outcome assessment (detection bias) | High risk | Judgement Comment: Not a blinded trial. Does not appear to have been a pre-specified analysis plan, the authors are aware of all outcomes and data before writing the article. |
| Incomplete outcome data (attrition bias) | Low risk | Judgement Comment: Report 94% of patients in the region should have followed up at involved health centers, making it fairly unlikely that many cases were missed. Reports that all deaths in this age group should be autopsied, also making it unlikely. |
| Selective reporting (reporting bias) | Low risk | Judgement Comment: Outcomes were checked via news reports, and autopsies which were done on all young deaths according to Italian statute. |
| Other bias | High risk | Judgement Comment: the authors suggest the likely cause of change in SCD rate over years; no consideration given to confounding factors, or attempts to describe participants, or to control for other variables that may have affected outcomes over the very different time periods |
| EPIDEMIOLOGY EXT VALIDITY: REPRESENTATIVE SAMPLE | Low risk | Judgement Comment: Cases described from participating medical centers (94%) of available population in these hospitals. |
| EPIDEMIOLOGY INTERNAL VALIDITY: CASE DEFINITION | Low risk | Judgement Comment: Cases ID'd from news reports and, primarily from hospital/autopsy records |
| EPIDEMIOLOGY EXT VALIDITY: SAMPLING/SUBJECTS INCLUDED | Low risk | Judgement Comment: Cases described from participating medical centers (94%) of available population in these hospitals. |
| EPIDEMIOLOGY INTERNAL VALIDITY: STUDY PERIOD | Low risk | > 100,000 person years |
| EPIDEMIOLOGY INTERNAL VALIDITY: DATA COLLECTION SOURCE | Low risk | Judgement Comment: All data collected from same source |
| EPIDEMIOLOGY INTERNAL VALIDITY: NUMERATOR AND DENOMINATOR | Low risk | Judgement Comment: Cases described from participating medical centers (94%) of available population in these hospitals. Some concern regarding the denominator as the population over 26 years is considered 'stable' which seems unlikely- |

| | | |
|---|---|---|
| | | the population would change either up or down over time. |
| EPIDEMIOLOGY INTERNAL VALIDITY: MODE OF DATA COLLECTION | Low risk | Judgement Comment: Data collected and identified from mandatory autopsy reports in the age group included |
| EPIDEMIOLOGY FINAL SUMMARY: | Low risk | |
| EPIDEMIOLOGY EXT VALIDITY: NONRESPONSE BIAS/FOLLOW UP OF COHORT | N/A | |

#### *Drezner 2005*

| | |
|---|---|
| **Methods** | **Study design:** Cross Sectional Survey asking for data occurring at the university in the past.<br>**Cross sectional:** Survey of athletic trainers at division 1 universities. 75% response rate. Survey goal was the assessment of automated external defibrillator use, and the reporting of an incidence rate was a secondary objective and report of the study |
| **Participants** | **Included criteria:** Division 1 university athletes<br>**Excluded criteria:** none reported<br>**Age:** Collegiate athletes, ages not reported<br>**Gender:** both<br>**Sport:** combination<br>**Level of Sport:** elite university athletes<br>**ECG Screened:** not described |
| **Interventions** | Observational |
| **Outcomes** | **SCA:** yes<br>**SCD:** yes<br>**Exertional:** not described<br>**Method of ID cases:** site report based on survey response |
| **Identification** | **Sponsorship source:** none listed<br>**Country:** USA<br>**Setting:** Division 1 NCAA university athletic programs<br>**Comments:** Cross sectional survey of athletic trainers at universities. 75% response rate.<br>**Authors name:** Jonathan Drezner, MD<br>**Institution:** University of Washington<br>**Email:** |

| | |
|---|---|
| | **Address:** Department of Family Medicine University of Washington Box 3547754245 Roosevelt Way NE Seattle, WA 98105 |
| **Notes** | |

Risk of bias table

| **Bias** | **Authors' judgement** | **Support for judgement** |
|---|---|---|
| EPIDEMIOLOGY EXT VALIDITY: REPRESENTATIVE SAMPLE | Low risk | Judgement Comment: 75% response rate to survey, so relatively good response and like representative. |
| EPIDEMIOLOGY INTERNAL VALIDITY: CASE DEFINITION | High risk | Quote: "This study was a retrospective survey. Questionnaires were mailed to head athletic trainers at all Division I universities (N 326) in the NCAA with three follow-up requests sent via e-mail over a 4-month period (August to November 2003). Head athletic trainers are in part responsible for determining the type of medical services at university sporting events and were thought to be the most reliable source of information regarding the presence and past use of AED."<br>Judgement Comment: Not detailed how the cases were determined. |
| EPIDEMIOLOGY EXT VALIDITY: SAMPLING/SUBJECTS INCLUDED | Low risk | Judgement Comment: All NCAA D1 schools were included in the survey attempt. 75% responded. NCAA div 1 athletes |
| EPIDEMIOLOGY INTERNAL VALIDITY: STUDY PERIOD | Low risk | Judgement Comment: Over 100,000 athlete years |
| EPIDEMIOLOGY INTERNAL VALIDITY: DATA COLLECTION SOURCE | Low risk | Judgement Comment: Data appear to have been collected via same source-which was ATC at schools surveyed. |
| EPIDEMIOLOGY INTERNAL VALIDITY: NUMERATOR AND DENOMINATOR | Low risk | Judgement Comment: Denominator appears appropriate based on report of athletes present at the universities included. The 5 events of SCD were reported in response to a survey. Risk of bias that this number is related to response bias, but may only slightly overestimate the incidence of SCD in Division 1 college athletes based on 75% response rate to the survey. |

| | | |
|---|---|---|
| EPIDEMIOLOGY INTERNAL VALIDITY: MODE OF DATA COLLECTION | High risk | Judgement Comment: Done via survey. Unclear how the follow up data was collected. |
| EPIDEMIOLOGY FINAL SUMMARY: | Moderate risk | Judgement Comment: Some risk of bias inherent in any survey, but very high response rate in this study, and may only slightly over-estimate the incidence with response bias. There is lack of clarity on case definition, and how the data on deaths was obtained. It appears this came from ATCs at universities, with no back up data from medical reports/autopsies, etc. The primary goal of this survey was to survey for AED use, not to produce an incidence rate. |
| EPIDEMIOLOGY EXT VALIDITY: NONRESPONSE BIAS/FOLLOW UP OF COHORT | Low risk | Judgement Comment: 75% response rate. Even with high response rate, some concerns about responder/recall bias to the survey (those with events more likely to respond), but this is an overall good rate, believe lessens risk of bias. |

#### *Drezner 2009*

| | |
|---|---|
| **Methods** | **Study design:** Cross-Sectional Survey<br>**Cross-sectional:** Survey of high schools using the national registry for AED use in sport. 11% of 18974 HS responded. |
| **Participants** | **Included criteria:** high school athlete<br>**Excluded criteria:** none listed<br>**Age:** 14-19 (based on level of school)<br>**Gender:** both<br>**Sport:** combination<br>**Level of Sport:** scholastic<br>**ECG Screened:** no |
| **Interventions** | Observational |
| **Outcomes** | **SCA:** yes<br>**SCD:** yes<br>**Exertional:** not described<br>**Method of ID Cases:** report from facility based on survey response |
| **Identification** | **Sponsorship source:** National operating committee on standards for athletic equipment<br>**Country:** USA |

| | |
|---|---|
| | **Setting:** High schools in USA<br>**Comments:**<br>**Authors name:** Jonathan Drezner, MD<br>**Institution:** University of Washington<br>**Email:**<br>**Address:** Department of Family Medicine University of Washington Box 3547754245 Roosevelt Way NE Seattle, WA 98105 |
| **Notes** | |

Risk of bias table

| Bias | Authors' judgement | Support for judgement |
|---|---|---|
| EPIDEMIOLOGY EXT VALIDITY: REPRESENTATIVE SAMPLE | High risk | Quote: "Letters requesting participation in the study were sent to 18,974 member high schools from the National Federation of High Schools in December 2006 and again in March 2007. A school representative (principal, athletic director, or certified athletic trainer) was invited to complete a comprehensive survey on emergency response planning for SCA and to provide details and outcomes of any AED use for SCA occurring within 6 months of survey completion."<br>Judgement Comment: Attempted to contact all high schools in the National federation of High schools on 2 separate occasions to complete survey. Only 11% responded |
| EPIDEMIOLOGY INTERNAL VALIDITY: CASE DEFINITION | Low risk | Quote: "Additional analyses were directed toward the details of resuscitation in cases of SCA. Any school reporting a case of SCA in a student athlete was also contacted by phone to review survey responses and to clarify the details of resuscitation. All surveys were completed between December 2006 and July 2007. The primary outcome measure for effectiveness was survival to hospital discharge after SCA. Secondary outcome measures relative to the adequacy of emergency response planning included the proportion of schools with an established emergency action plan (EAP) for SCA, CPR and AED training for coaches and other school staff members, and review and practice of the EAP at least once annually. Secondary outcome measures in cases of SCA included the presence of seizure-like activity |

| | | |
|---|---|---|
| | | after collapse, provision of by- stander CPR, reported time to CPR and to initial shock deployment, and cause of SCA. This study was approved by"<br>Judgement Comment: followed up by phone interviews, and to gather follow up data on cases |
| EPIDEMIOLOGY EXT VALIDITY: SAMPLING/SUBJECTS INCLUDED | Low risk | Judgement Comment: attempted contact with all high schools suggests attempt at sampling all possible sources. |
| EPIDEMIOLOGY INTERNAL VALIDITY: STUDY PERIOD | Low risk | > 100,000 athlete years |
| EPIDEMIOLOGY INTERNAL VALIDITY: DATA COLLECTION SOURCE | Low risk | Judgement Comment: Collected from contacts at participating high school, as well as phone interviews with witnesses, etc. |
| EPIDEMIOLOGY INTERNAL VALIDITY: NUMERATOR AND DENOMINATOR | High risk | Judgement Comment: Calculation of the denominator is appropriate for this study; however the numerator suffers a high risk of responder bias as this was a x-sectional study, with 11% response from high schools contacted. |
| EPIDEMIOLOGY INTERNAL VALIDITY: MODE OF DATA COLLECTION | Low risk | Judgement Comment: All data collected in same way- via survey and follow up phone calls |
| EPIDEMIOLOGY FINAL SUMMARY: | High risk | Quote: "This study found an annual incidence of SCA in high school student athletes of 4.4 in 100 000. Although this estimate may be influenced by responder bias, it is consistent with recent findings from a prospective, population-based study of pediatric out-of-hospital cardiac arrest in which the incidence of SCA caused by cardiovascular disease in adolescents (age, 14 to 24 years) was found to be 3.75 in 100 000. 31 It is also consistent with findings from the Veneto region of Italy, which uses a regional registry for juvenile sudden death and where a baseline incidence of sudden cardiac death in young competitive athletes (age, 12 to 35 years) of 3.6 in 100 000 was found before the implementation of a national screening program."<br>Judgement Comment: Study designed to detail use and outcomes of automated external defibrillator use in high schools. The incidence statistic provided is result of 11% response to a cross sectional survey- |

| | | |
|---|---|---|
| | | and the authors themselves note the risk for bias in this statistic. For the purposes of the review, this is a study with high risk of bias. |
| EPIDEMIOLOGY EXT VALIDITY: NONRESPONSE BIAS/FOLLOW UP OF COHORT | High risk | Judgement Comment: 11% response to survey requests. Suggests responder bias (acknowledged by authors)/nonresponder bias. |

#### *Durakovic 2012*

| | |
|---|---|
| **Methods** | **Study design:** Retrospective cohort study<br>**Retrospective cohort:** Unclear how the cases were identified or what data base was searched to identify them. Also unclear where the denominator number came from. |
| **Participants** | **Included criteria:** Competitive athletes in Croatia<br>**Excluded criteria**: not described<br>**Age:** 17-29 for deaths, unclear what ages were considered eligible<br>**Gender:** male<br>**Sport:** combination<br>**Level of Sport:** competitive<br>**ECG Screened:** not described |
| **Interventions** | **Intervention Characteristics** |
| **Outcomes** | **SCA:** no<br>**SCD:** yes<br>**Exertional:** not described<br>**Method of ID Cases:** combination of medical records and autopsy reports |
| **Identification** | **Sponsorship source:** none<br>**Country:** Croatia<br>**Setting:** Competitive male athletes in Croatia<br>**Comments:** Unclear how comprehensive the data is<br>**Authors name:** Zijad Durakovic MD, PHd<br>**Institution:** Rebro University School of Medicine and University Hospital Center<br>**Email:** |

| | |
|---|---|
| | **Address:** Dept Internal Medicine Rebro University School of Medicine and University Hospital Center Clinic Hospital Center Kispaticeva Str. 12,10000 Zagreb, Croatia |
| **Notes** | most recent in multiple publications about athlete deaths in Croatia |

Risk of bias table

| **Bias** | **Authors' judgement** | **Support for judgement** |
|---|---|---|
| EPIDEMIOLOGY EXT VALIDITY: REPRESENTATIVE SAMPLE | Unclear risk | Judgement Comment: Record was reported on competitive athletes. There is no description of how the sudden deaths were identified in athletes from non-athletes. There may be under-representation. |
| EPIDEMIOLOGY INTERNAL VALIDITY: CASE DEFINITION | Low risk | Judgement Comment: ID'd cases come from autopsy records. |
| EPIDEMIOLOGY EXT VALIDITY: SAMPLING/SUBJECTS INCLUDED | Unclear risk | Judgement comment: All competitive male athletes in Croatia considered. |
| EPIDEMIOLOGY INTERNAL VALIDITY: STUDY PERIOD | Low risk | > 100,000 person years |
| EPIDEMIOLOGY INTERNAL VALIDITY: DATA COLLECTION SOURCE | Low risk | Judgement Comment: All from autopsy records |
| EPIDEMIOLOGY INTERNAL VALIDITY: NUMERATOR AND DENOMINATOR | Unclear risk | Judgement Comment: Unclear how complete the numerator is; and unclear where the denominator number is derived. |
| EPIDEMIOLOGY INTERNAL VALIDITY: MODE OF DATA COLLECTION | Unclear risk | Judgement Comment: All cases were identified from records from multiple sources. It is unclear however how the authors searched for and identified these cases of death and athletes, and why or why not this was comprehensive. |
| EPIDEMIOLOGY FINAL SUMMARY: | High risk | Judgement Comment: There are significant unkowns in this trial. It is not clear how cases of sudden death were identified in athletes vs. non-athletes; it is unclear how comprehensive the records of death are, how the information was collected at the time of death, how the authors searched for the death records and identification. |
| EPIDEMIOLOGY EXT VALIDITY: NONRESPONSE | N/A | |

| |
|---|
| BIAS/FOLLOW UP OF COHORT |

#### *Eckart 2004*

| | |
|---|---|
| **Methods** | **Study design:** Retrospective cohort study<br>**Retrospective cohort:** Data collected from military records autopsy and death registry data. |
| **Participants** | **Included criteria:** military recruits<br>**Excluded criteria:** none<br>**Age:** N17-35<br>**Gender:** both<br>**Sport:** military<br>**Level of Sport:** military recruits<br>**ECG Screened:** no |
| **Interventions** | Observational |
| **Outcomes** | **SCA:** no<br>**SCD:** yes<br>**Exertional:** no, appears to be all deaths<br>**Method of ID Cases:** autopsy data and military death registry |
| **Identification** | **Sponsorship source:** None listed<br>**Country:** USA<br>**Setting:** Military recruits from 4 major military branches<br>**Comments:** NA<br>**Authors name:** Robert E. Eckart, DO<br>**Institution:** Brooke Army Medical Center, San Antonio, Texas<br>**Email:**<br>**Address:** Cardiac Arrhythmia Service (Cardiovascular Division), Brooke Army Medical Center3851 Roger Brooke Drive San Antonio, Texas 78234 |
| **Notes** | Denominator for annual incidence created by using a multiplier on deaths to create an annualized rate, and then using 6.3 million recruits as denominator (in article). In our data set, the number of recruits was multiplied by 8/52 (proportion of weeks in training) to give a total person years in training. |

Risk of bias table

| Bias | Authors' judgement | Support for judgement |
|---|---|---|
| EPIDEMIOLOGY EXT VALIDITY: REPRESENTATIVE SAMPLE | Low risk | Judgement Comment: Active young population; may be difficult to compare to athletes because recruits and some may be unchanged. |
| EPIDEMIOLOGY INTERNAL VALIDITY: CASE DEFINITION | Low risk | Judgement Comment: Cases defined by autopsy and further military records. Cases without clear cardiac cause were not counted as cardiac. |
| EPIDEMIOLOGY EXT VALIDITY: SAMPLING/SUBJECTS INCLUDED | Low risk | Judgement Comment: Full population of recruits involved in 4 main branches of military |
| EPIDEMIOLOGY INTERNAL VALIDITY: STUDY PERIOD | Low risk | > 100,000 person years |
| EPIDEMIOLOGY INTERNAL VALIDITY: DATA COLLECTION SOURCE | Low risk | Judgement Comment: Data collected from military records autopsy and death registry data. |
| EPIDEMIOLOGY INTERNAL VALIDITY: NUMERATOR AND DENOMINATOR | Low risk | Judgement Comment: Military records used for both. There is a question of using only the identifiable cardiac deaths, vs. cardiac + idiopathic deaths (64+44). Denominator is adjusted for 8 weeks of basic training by multiplying x 6.5 to create an annual incidence rate. |
| EPIDEMIOLOGY INTERNAL VALIDITY: MODE OF DATA COLLECTION | Low risk | Judgement Comment: Cases identified via same method |
| EPIDEMIOLOGY FINAL SUMMARY: | Low risk | Judgement Comment: Methodology for data collection, reasons for classifying/not classifying as cardiac death laid out. Data sources from military records. |
| EPIDEMIOLOGY EXT VALIDITY: NONRESPONSE BIAS/FOLLOW UP OF COHORT | N/A | |

#### *Eckart 2011*

| | |
|---|---|
| **Methods** | **Study design:** Retrospective cohort study |

| | |
|---|---|
| | **Retrospective cohort:** Not clear how this was planned, but appears to be retrospective evaluation of data collected in real time. |
| **Participants** | **Included criteria:** Military active duty and recruits<br>**Excluded criteria:** NA<br>**Age:** All ages on active duty included. The ages extracted are 18-34<br>**Gender:** both<br>**Sport:** military<br>**Level of Sport:** active duty military<br>**ECG Screened:no** |
| **Interventions** | Observational |
| **Outcomes** | **SCA:** no<br>**SCD:** yes<br>**Exertional:** no, all deaths<br>**Method of ID Cases:** |
| **Identification** | **Sponsorship source:** Air Force Medical Research Program<br>**Country:** USA<br>**Setting:** US Military active duty<br>**Authors name:** Robert E. Eckart<br>**Institution:** Cardiac Arrhythmia Service (Cardiovascular Division), Brooke Army Medical Center<br>**Email:**<br>**Address:** Cardiac Arrhythmia Service (Cardiovascular Division), Brooke Army Medical Center3851 Roger Brooke Drive San Antonio,Texas 78234 |
| **Notes** | Data may double count a small proportion of sudden deaths in recruits only from 1998-2001 also included in 2004 Eckart article. |

Risk of bias table

| **Bias** | **Authors' judgement** | **Support for judgement** |
|---|---|---|
| EPIDEMIOLOGY EXT VALIDITY: REPRESENTATIVE SAMPLE | Low risk | Judgement Comment: Intended to be the military population, when compared with the general population, more male, younger. THe authors point out possibility of ascertainment bias as some may have been ruled out of service due to medical/cardiac issues. |
| EPIDEMIOLOGY INTERNAL VALIDITY: CASE DEFINITION | Low risk | Quote: "Of the 2,729 remain- ing cases, there were 1,044 nontraumatic suspected cardiac deaths identified from 1998 to 2008. Of these, we excluded 51 (5.1%) subjects for lack of clinical record or autopsy, 130 (12.5%) subjects for unavailability of records, and 12 (1.2%) subjects for what was |

| | | |
|---|---|---|
| | | determined to be a clear noncardiac etiology. We identified 902 subjects with full records available for review for whom the adjudicated cause of death was of potential cardiac etiology, which serves as our cohort."<br>Judgement Comment: Adjudicated cause of death panel; access to records for about 80% of deaths. |
| EPIDEMIOLOGY EXT VALIDITY: SAMPLING/SUBJECTS INCLUDED | Low risk | Judgement Comment: all active duty military. Deaths of all active duty included. |
| EPIDEMIOLOGY INTERNAL VALIDITY: STUDY PERIOD | Low risk | > 100,000 person years |
| EPIDEMIOLOGY INTERNAL VALIDITY: DATA COLLECTION SOURCE | Low risk | Quote: "For each death, a detailed report to include autopsy is filed with the Armed Forces Institute of Pathology in accordance with established protocol (26). Although not all autopsies were performed at the Armed Forces Institute of Pathology, in Washington, DC, the Office of the Armed Forces Medical Examiner system appoints Regional Medical Examiners, board certified in forensic examination by the American Board of Pathology, to serve as worldwide consultants." |
| EPIDEMIOLOGY INTERNAL VALIDITY: NUMERATOR AND DENOMINATOR | High risk | Quote: "For each death, a detailed report to include autopsy is filed with the Armed Forces Institute of Pathology in accordance with established protocol (26). Although not all autopsies were performed at the Armed Forces Institute of Pathology, in Washington, DC, the Office of the Armed Forces Medical Examiner system appoints Regional Medical Examiners, board certified in forensic examination by the American Board of Pathology, to serve as worldwide consultants."<br>Judgement Comment: Military records for numerator and denominator. Numerator may be undercounted-181 of 1044 possible cases were excluded for lack of records. 902 as final number of deaths included. 715 cardiac deaths, 187 sudden unexplained deaths. < 35 age group 298 total SCD + 123 SUD. Could change the incidence by 25%-but this is laid out and explained by authors |

| | | |
|---|---|---|
| EPIDEMIOLOGY INTERNAL VALIDITY: MODE OF DATA COLLECTION | Low risk | Judgement Comment: All data via military records. Subjects excluded where records not available |
| EPIDEMIOLOGY FINAL SUMMARY: | Low risk | Judgement Comment: Authors very clear about their methodology, their data collection methods appear very appropriate. They final numbers may slightly undercount the sudden cardiac rate as the numbers are presented as sudden unexplained death and sudden cardiac death, and not combined. |
| EPIDEMIOLOGY EXT VALIDITY: NONRESPONSE BIAS/FOLLOW UP OF COHORT | N/A | |

#### *Endres 2019*

| | |
|---|---|
| **Methods** | **Study design:** Retrospective cohort study<br>**Retrospective cohort:** Media search study |
| **Participants** | **Included criteria:** Cases: publicly reported deaths in media outlets in athletes between 6 and 17 years old (not including HS scholastic athletes) who died in middle school athletics, youth leagues or recreational leagues or training for these teams.<br>**Excluded criteria:** Under age 5. Occurred outside of the youth, recreational, or middle school team activity. Occurred in HS or college athletics<br>**Age:** 6-17<br>**Gender:** both<br>**Sport:** combination<br>**Level of Sport:** combination-youth and scholastic<br>**ECG Screened:** no |
| **Interventions** | Observational |
| **Outcomes** | **SCA:** no<br>**SCD:** yes<br>**Exertional:** yes<br>**Method of ID Cases:** media/web reports |
| **Identification** | **Sponsorship source:** None<br>**Country:** USA<br>**Setting:** Organized American youth sports<br>**Authors name:** Brad D. Endres, MS, ATC, CSCS |

| | |
|---|---|
| | **Institution:** Korey Stringer Institute, Department of Kinesiology, University of Connecticut, Storrs<br>**Email:**<br>**Address:** Korey Stringer Institute, Department of Kinesiology, University of Connecticut 2095 Hillside Road U-1110Storrs, CT 06269 |
| **Notes** | **Comments:** Does not include high school athletes scholastic sports in the study, though they are eligible based on age for non school based sports only. |

Risk of bias table

| **Bias** | **Authors' judgement** | **Support for judgement** |
|---|---|---|
| EPIDEMIOLOGY EXT VALIDITY: REPRESENTATIVE SAMPLE | Low risk | Judgement Comment: all youth athletes, excepting high school athletes participating in HS sports |
| EPIDEMIOLOGY INTERNAL VALIDITY: CASE DEFINITION | High risk | Quote: "death in the emergency department. . Sudden death data from cases occurring between August 1, 2007, and December 31, 2015, were obtained via 2 methods: (1) a search using the LexisNexis Academic Database and (2) other Internet searches (eg, Google) of publicly available news reports. The search for sudden death cases began in August of 2013 using the terms death, fatality, exercise, race, sport, practice, and ''game.'' Information related to the athlete (age, sex), event (sport, level of play, event type), and death (date, location, official cause, and speculated cause of death) was obtained. Deaths were reviewed by researchers (B.D.E., R.L.S.) and classified into 5 event types (cardiac/cardiovascular, exertional/heat/ environmental, traumatic injury, other, or inconclusive/ unknown). If an official cause of death was not provided in the media source, 1 member of the research team (B.D.E.) reviewed all available information and provided a speculated cause of death. For example, if an official cause of death was not reported for an athlete who collapsed and died despite an advised shock from an automated external defibrillator, the case was classified as cardiac."<br>Judgement Comment: Using only internet sources; and if no COD was listed, the COD was 'speculated' upon by one of the authors. Risks misclassifying deaths. |

| | | |
|---|---|---|
| EPIDEMIOLOGY EXT VALIDITY: SAMPLING/SUBJECTS INCLUDED | High risk | Judgement Comment: Leaves out incidents in scholastic competitions for high school aged athletes, though they are included in the age group, likely giving an under estimate of the incidence. |
| EPIDEMIOLOGY INTERNAL VALIDITY: STUDY PERIOD | Low risk | > 100,000 person years |
| EPIDEMIOLOGY INTERNAL VALIDITY: DATA COLLECTION SOURCE | High risk | Judgement Comment: Internet and Lexis/Nexis sourcing |
| EPIDEMIOLOGY INTERNAL VALIDITY: NUMERATOR AND DENOMINATOR | High risk | Quote: "An athlete-year was defined as 1 youth athlete participating in 1 sport in a calendar year. 16 Athletes may have participated in more than 1 sport in a calendar year and were counted separately for each sport."<br>Judgement Comment: The total participant count has significant risk of counting athletes who participate in more than one sport per year mutiple times, causing an overestimation of the denominator. Numerator is an underrepresentation of 'youth sports' as it excludes high school athletes, but includes high school aged athletes when playing non school sanctioned sports, under counting the total number of deaths. Likely double counting of many athletes who participated in more than one sport per year (thus counting as 2 athlete years), again likely expanding the denominator. |
| EPIDEMIOLOGY INTERNAL VALIDITY: MODE OF DATA COLLECTION | Low risk | Judgement Comment: all sources had data collected in same way |
| EPIDEMIOLOGY FINAL SUMMARY: | High risk | Judgement Comment: Risks of bias with the numerator (internet/news sources) and denominator (including HS age groups, but not including deaths in HS sports-likely expanding denominator, and decreasing numerator) are significant. There is also ROB with regards to the categorization of cause of death in the subjects. This may be the best one may be able to do in this age group, but still risks a significant amount of bias. |
| EPIDEMIOLOGY EXT VALIDITY: NONRESPONSE | N/A | |

| |
|---|
| BIAS/FOLLOW UP OF COHORT |

#### *Farioli 2015*

| | |
|---|---|
| **Methods** | **Study design:** Retrospective cohort study<br>**Retrospective cohort:** Data collected prospectively, but idea for project, calculation of denominator, etc. done in retrospective manner. |
| **Participants** | **Baseline Characteristics**<br>Overall<br>**Included criteria:** Male firefighters in USA<br>**Age:** 18-34<br>**Gender: m**ale<br>**Sport:** firefighters, considered military for purposes of this review<br>**Level of Sport:** firefighters, considered military for purposes of this review<br>**ECG Screened:** no |
| **Interventions** | Observational |
| **Outcomes** | **SCA: no**<br>**SCD:** yes<br>**Exertional:** no, all deaths which occurred on duty<br>**Method of ID Cases:** Autopsy and other official data collection agency of firefighters |
| **Identification** | **Sponsorship source:** U.S. Department of Homeland Security<br>**Country:** USA<br>**Setting:** Male firefighters in US<br>**Comments:** NA<br>**Authors name:** Andrea Farioli, MD, Stefanos N. Kales, MD, MPH(last)<br>**Institution:** Department of Environmental Health (Environmental & Occupational Medicine & Epidemiology), Harvard TH Chan School of Public Health, Boston<br>**Email:**<br>**Address:** Cambridge Hospital Macht Building 427, 1493 Cambridge Street, Cambridge, MA 02139 |
| **Notes** | *Aaron Lear* on 12/05/2019 23:54<br>**Select**<br>firefighters rather than military-so wrong study population-but should |

| | |
|---|---|
| | probably include bc of similarity with military. Will take some work to get stats right. |

Risk of bias table

| Bias | Authors' judgement | Support for judgement |
|---|---|---|
| EPIDEMIOLOGY EXT VALIDITY: REPRESENTATIVE SAMPLE | Low risk | Judgement Comment: seeks to explain incidence in male firefighters |
| EPIDEMIOLOGY INTERNAL VALIDITY: CASE DEFINITION | Low risk | Quote: "SCD was defined as an unexpected death of cardiac origin that occurred within 1 hour of the onset of symptoms (witnessed) or within 24 hours of having been seen alive and without symptoms (unwitnessed). In our analysis, we included only cardiovascular deaths in which, after the collapse, the person never regained consciousness prior to biological death. " |
| EPIDEMIOLOGY EXT VALIDITY: SAMPLING/SUBJECTS INCLUDED | Low risk | Judgement Comment: Appears to include all possible firefighter deaths, and a reasonable estimate of total firefighters active based on Census statistics |
| EPIDEMIOLOGY INTERNAL VALIDITY: STUDY PERIOD | Low risk | > 100,000 person years |
| EPIDEMIOLOGY INTERNAL VALIDITY: DATA COLLECTION SOURCE | Low risk | Judgement Comment: See quotes on data collection source. From USFA, and NIOSH-mandatory reporting of all firefighter deaths by law in US. USFA collects this data. |
| EPIDEMIOLOGY INTERNAL VALIDITY: NUMERATOR AND DENOMINATOR | Low risk | Quote: " Methods Study Population. The study population is the dynamic cohort of 300 000 US full-time male career firefighters employed between January 1, 1998, and December 31, 2012. Only firefighters aged 18 to 64 years were included in the cohort. The CPS, conducted by the US Census Bureau for the Bureau of Labor Statistics, is the primary source of labor statistics in the United States."<br>Quote: "The USFA actively collects information of firefighter deaths directly from fire services and from many external sources, including the USFA Public Safety Officers' Benefits program administered by the US Department of Justice, NIOSH, the Occupational Safety and Health |

| | | |
|---|---|---|
| | | Administration, the US Department of Defense, the National Interagency Fire Center, and other federal agencies."<br>Quote: "The NIOSH program aims to investigate firefighter line-of- duty deaths for prevention purposes. The NIOSH database is neither representative nor comprehensive; however, all reports present a detailed description of the event and, if relevant, a summary of the clinical history (including emergency medical services records) and postmortem examination findings."<br>Judgement Comment: Potential for undercount, as only on duty deaths were included. Person years were created by using census data for hours worked, and turning this into person years. |
| EPIDEMIOLOGY INTERNAL VALIDITY: MODE OF DATA COLLECTION | Low risk | Quote: "Two physicians independently examined the summary report of each record."<br>Quote: "We examined all USFA records for on-duty fatalities that occurred between January 1998 and December 2012 that were listed as heart attacks, cerebrovascular accidents, heat exhaustion, or “other.” We also reviewed all “medical related” deaths from the NIOSH database. Two physicians independently examined the summary report of each record."<br>Quote: "From NIOSH records, we were also able to retrieve information on the presence of a shockable rhythm during resuscitation efforts (as assessed through the use of automated external defibrillators). We classified an event as on duty if the onset of the symptoms occurred during the firefighter’s work shift. For cases in which the information extracted from the USFA and NIOSH reports were not in agreement, we relied on the more comprehensive narrative information provided by the NIOSH reports."<br>Quote: "We collected on-duty SCD data from the USFA and from the NIOSH Fire Fighter Fatality Investigation and Prevention Program. The USFA maintains a systematic database of all deaths associated with firefighting in the United States since 1981. 14 Of note, identifying and reporting cardiac death among firefighters is mandatory in the |

| | | |
|---|---|---|
| | | United States (section 1201 of the Omnibus Crime Control and Safe Streets Act of 1968 [42 U.S.C. 3796] and Hometown Heroes Survivors Benefits Act of 2003)." |
| EPIDEMIOLOGY FINAL SUMMARY: | Low risk | Judgement Comment: Authors explain their methodology very clearly, including how they developed number of firefighter years with formulas. The data sources appear to be reliable. |
| EPIDEMIOLOGY EXT VALIDITY: NONRESPONSE BIAS/FOLLOW UP OF COHORT | N/A | |

#### *Fuller 1997*

| | |
|---|---|
| **Methods** | **Study design:** Prospective Cohort study<br>**Prospective cohort:** Prospective cohort of community screening project |
| **Participants** | **Included criteria:** High school athlete at participating high school screening program<br>**Excluded criteria:** NA<br>**Age:** 14-19 (High school age)<br>**Gender:** both<br>**Sport:** combination<br>**Level of Sport:** scholastic<br>**ECG Screened:** yes |
| **Interventions** | observational |
| **Outcomes** | **SCA:** yes<br>**SCD:** yes<br>**Exertional:** not described<br>**Method of ID Cases:** not described |
| **Identification** | **Sponsorship source:** Sierra Nevada Cardiology Associates<br>**Country:** USA<br>**Setting:** Reno, NV area high schools<br>**Comments:** NA |

| | |
|---|---|
| | **Authors name:** Collin M. Fuller, MD<br>**Institution:** Sierra Nevada Cardiology Associates; Division of Cardiology<br>**Email:** NA<br>**Address:** Sierra Nevada Cardiology Associates 75 Pringle Way, Suite 401Reno, NV 89502 |
| **Notes** | |

Risk of bias table

| Bias | Authors' judgement | Support for judgement |
|---|---|---|
| EPIDEMIOLOGY EXT VALIDITY: REPRESENTATIVE SAMPLE | Low risk | Judgement Comment: Appears to be a report of community service screening. The authors set out to report this activity, not broadly apply their data. They report an estimated 95% attendance of athletes at 30 area high schools. |
| EPIDEMIOLOGY INTERNAL VALIDITY: CASE DEFINITION | Unclear risk | Judgement Comment: This is not detailed in article |
| EPIDEMIOLOGY EXT VALIDITY: SAMPLING/SUBJECTS INCLUDED | Unclear risk | Quote: "Subjects. In conjunction with the approval of school officials and coaches, male and female student athletes at 30 selected high schools in Northern Nevada were invited to undergo, without charge, preparticipation screening. Schools were chosen based on their geographic proximity to urban and rural cardiology clinics of Sierra Nevada Cardiology Associates. All invited schools participated. Student"<br>Quote: "The exact percentage of student participation is not known, although it was estimated to be greater than 95% by school coaches."<br>Judgement Comment: No breakdown of athlete demographics, nor comparison to general athletic population was given. Proximity to private practice cardiology group performing community service appears to have been driving factor for recruitment. |
| EPIDEMIOLOGY INTERNAL VALIDITY: STUDY PERIOD | High risk | <100,000 person years |
| EPIDEMIOLOGY INTERNAL VALIDITY: DATA COLLECTION SOURCE | Unclear risk | Judgement Comment: Not detailed in article |

| | | |
|---|---|---|
| EPIDEMIOLOGY INTERNAL VALIDITY: NUMERATOR AND DENOMINATOR | Unclear risk | Quote: " Follow-up. During the 3-yr study period (1991-1994), no HAS (high school athlete) sustained sudden death during sports participation. One HSA developed ventricular fibrillation during track practice and was successfully resuscitated. " Judgement Comment: Unclear how the follow up on the students occurred, or how stable the total athletes screened were. Data the result of 3 year ECG screening program. Unclear how close the follow up of the athletes was; this is not detailed except to say that 1 had SCA. No explanation of how the cohort was followed. |
| EPIDEMIOLOGY INTERNAL VALIDITY: MODE OF DATA COLLECTION | Unclear risk | Judgement Comment: Not detailed in article |
| EPIDEMIOLOGY FINAL SUMMARY: | High risk | Judgement Comment: Study was designed to report results of screening program. The 3 year follow up appears to be an afterthought to the general reporting of the results of ECG/echo screening. Important data points about how the cohort was followed, how the records of single SCA event were identified, etc. were not included in the article. |
| EPIDEMIOLOGY EXT VALIDITY: NONRESPONSE BIAS/FOLLOW UP OF COHORT | N/A | |

#### *Grani 2016*

| | |
|---|---|
| **Methods** | **Study design:** Retrospective cohort study<br>**Retrospective cohort:** Data collected by autopsy at sudden death. These records reviewed for sport related sudden death. |
| **Participants** | **Included criteria:** SrSCD (sport related sudden cardiac death) as an unexpected cardiovascular death occurring during or within 1 hour after physical activity.<br>**Excluded criteria:** Deaths that were determined without an autopsy. |

| | |
|---|---|
| | **Pretreatment:** No demographics given<br>**Age:** 10-39<br>**Gender:** both<br>**Sport:** combination<br>**Level of Sport:** combination<br>**ECG Screened:** portion was screened |
| **Interventions** | observational |
| **Outcomes** | **SCA:** no<br>**SCD:** yes<br>**Exertional:** yes<br>**Method of ID Cases:** autopsy data |
| **Identification** | **Sponsorship source:** Unrestricted grant from Swiss Heart foundation<br>**Country:** Switzerland<br>**Setting:** General Population of Switzerland<br>**Comments:** none<br>**Authors name:** Christoph Grani; Matthias Wilhelm (last)<br>**Institution:** University Clinic for Cardiology, Inselspital Bern University Hospital Bern Bern, Switzerland<br>**Email:**<br>**Address:** Preventive Cardiology and Sports Medicine, Department of Cardiology, Inselspital, University Hospital and University of Bern, 3010 Bern, Switzerland |
| **Notes** | |

Risk of bias table

| **Bias** | **Authors' judgement** | **Support for judgement** |
|---|---|---|
| EPIDEMIOLOGY EXT VALIDITY: REPRESENTATIVE SAMPLE | Low risk | Quote: "all forensic reports of German- and French-speaking parts of Switzerland from 1999 to 2010 (with an overall population of 7,030,900) for SrSCDs in young individuals (10–39 years of age). We included only German- and French-speaking parts of Switzerland as some of the SrSCD cases of the Italian-speaking parts of Switzerland were investigated in Italy. "<br>Quote: ". Methods Study population: We retrospectively reviewed all forensic autopsy reports of the German-speaking part of Switzerland (population in the examined region is 5,617,963, which comprises nearly 70% of the Swiss population) for sudden unexpected deaths in individuals aged |

| | | |
|---|---|---|
| | | 10–39 years, occurring Sudden cardiac deaths in Switzerland" |
| EPIDEMIOLOGY INTERNAL VALIDITY: CASE DEFINITION | Low risk | Quote: "SrSCD as an unexpected cardiovascular death occurring during or within 1 hour after physical activity."<br>Judgement Comment: ScSCD determined by autopsy. It was defined as death within 1 hour of activity |
| EPIDEMIOLOGY EXT VALIDITY: SAMPLING/SUBJECTS INCLUDED | Low risk | Judgement Comment: denominator-or those in study from the sports commission sampling of the population Should be MODERATE ROB |
| EPIDEMIOLOGY INTERNAL VALIDITY: STUDY PERIOD | Low risk | > 100,000 person years |
| EPIDEMIOLOGY INTERNAL VALIDITY: DATA COLLECTION SOURCE | Unclear risk | Judgement Comment: Autopsy reports, but over 1/2 of cases were actually just from multiplier, so possibility of bias here. |
| EPIDEMIOLOGY INTERNAL VALIDITY: NUMERATOR AND DENOMINATOR | High risk | Quote: " (www.swissregard.ch). Data analysis: The population data were derived from the Swiss Federal Statistical Office and a survey on sports-participation in Switzerland from the Swiss Federal Office of Sports. The Swiss Federal office of Statistics provided yearly population data of residents aged 10–39 years from the German speaking cantons [14]. The survey on sports participation provided extensive data on sporting activities, as well as volume and intensity of training [15]. A core representative sample of more than 10,000 15- to 75-year-old residents was first questioned by telephone, followed by an online questionnaire on their sports participation. Children aged 10 to 14 years were assessed with specially adapted questionnaires. The survey provided the ratios of people engaged in recreational or competitive sports of the concerned age group. Based on this, denominators for our three categories were formed as follows: all residents aged 10–39 years (denominator for NONE category); subjects engaged in sports (denominator for REC); and participants of competitions (denominator for COMP). Average yearly incidences were calculated for the pre-screening (1999–2001), early screening (2002–2004), mid-screening (2005– 2007) and late screening (2008–2010) periods " |

| | | |
|---|---|---|
| | | Quote: "Since our cohort included only autopsied SCD cases, we multiplied the recorded numbers by a factor of 2.1 to adjust the numbers of SCDs for the average autopsy rate of 47.5% in Switzerland in order to estimate real incidences"<br>Judgement Comment: The use of the multiplier to determine the cases raises significant concerns for the validity of the case number. Risks overestimation of cases. Numerator appears an accurate methodology |
| EPIDEMIOLOGY INTERNAL VALIDITY: MODE OF DATA COLLECTION | Low risk | Quote: "Data were retrospectively collected in an anonomised fashion in the Swiss Registry of Athletic Related Death (www.swissregard.ch). 12"<br>Judgement Comment: From autopsy data |
| EPIDEMIOLOGY FINAL SUMMARY: | Moderate risk | Judgement comment: well done, transparent article. Concern about using the multiplier to come to a denominator number |
| EPIDEMIOLOGY EXT VALIDITY: NONRESPONSE BIAS/FOLLOW UP OF COHORT | N/A | |

#### *Harmon 2015*

| | |
|---|---|
| **Methods** | **Study design:** Prospective cohort study<br>**Prospective cohort:** Data collected prospectively from Parent Heart Watch and NCAA databases. This was the final article of series on this database. |
| **Participants** | **Included criteria:** NCAA athletes with sudden death at any time determined to be Cardiac Related<br>**Excluded criteria:** Deaths for other causes<br>**Pretreatment:** no group data presented<br>**Age:** Athletes 17 to 24 years of age (university age)<br>**Gender:** both<br>**Sport:** combination<br>**Level of Sport:** combination (elite division one, and all university)<br>**ECG Screened:** not described (probably a portion were) |
| **Interventions** | Observational |
| **Outcomes** | **SCA:** no |

| | |
|---|---|
| | **SCD:** yes<br>**Exertional:** no, all deaths<br>**Method of ID Cases:** combination of autopsy, NCAA records, web/media database |
| **Identification** | **Sponsorship source:** One author NIH grant<br>**Country:** USA<br>**Setting:** Collegiate athletes<br>**Comments:** NA<br>**Authors name:** Kim Harmon, MD<br>**Institution:** Univ of Washington<br>**Email:**<br>**Address:** 3800 Montlake Blvd Seattle, WA 98195 |
| **Notes** | |

Risk of bias table

| **Bias** | **Authors' judgement** | **Support for judgement** |
|---|---|---|
| EPIDEMIOLOGY EXT VALIDITY: REPRESENTATIVE SAMPLE | Low risk | Judgement Comment: Study population is NCAA (national collegiate athletic association) athletes, college level (only about 1/3 of Americans attend 4 year colleges) and most people that play in college are higher level than club/scholastic athletes. This sample is clear about what it is meant to represent, which are athletes at this level, but is unlikely to be applicable to broader American public. |
| EPIDEMIOLOGY INTERNAL VALIDITY: CASE DEFINITION | Low risk | Quote: "The medical causes were further broken down into cardiac, cancer, heat stroke, sickle cell trait, sport-related head injury, meningitis, and other. If the cause of death could not be reasonably determined, it was recorded as unknown."<br>Judgement Comment: Cases made, and described as to how they came to conclusion; adjudication committee; combo of reports of NCAA, autopsy, other. Table 6 describes. |
| EPIDEMIOLOGY EXT VALIDITY: SAMPLING/SUBJECTS INCLUDED | Low risk | Judgement Comment: all NCAA athletes included |
| EPIDEMIOLOGY INTERNAL VALIDITY: STUDY PERIOD | Low risk | Judgement Comment: 11 years of data of all NCAA athletes; athlete years numbering into the millions |

| | | |
|---|---|---|
| EPIDEMIOLOGY INTERNAL VALIDITY: DATA COLLECTION SOURCE | Low risk | Quote: "Deaths in NCAA athletes were identified during the school years (July 1 to June 30) from 2003 to 2004 to 2012 to 2013 through (1) the NCAA Resolutions List, (2) the Parent Heart Watch database, and (3) NCAA insurance claims." Judgement Comment: triple resourced to make sure of as few misses as possible. |
| EPIDEMIOLOGY INTERNAL VALIDITY: NUMERATOR AND DENOMINATOR | Low risk | Quote: "Demographic data in NCAA athletes were obtained from the NCAA Sports Sponsorship and Participation Rates Report 20 and the NCAA Student-Athlete Ethnicity Report. 21" |
| EPIDEMIOLOGY INTERNAL VALIDITY: MODE OF DATA COLLECTION | Low risk | Judgement Comment: Triple attempted on all subjects. Not all cases were ID'd in the same manner, but adjudication panel present for all of them. Authors deliver list of which cases were detected which way. Very transparent. |
| EPIDEMIOLOGY FINAL SUMMARY: | Low risk | Judgement comment: Well done article, good sources, and reliably calculated. Transparent methodology. |
| EPIDEMIOLOGY EXT VALIDITY: NONRESPONSE BIAS/FOLLOW UP OF COHORT | N/A | |

#### *Harmon 2016*

| | |
|---|---|
| **Methods** | **Study design:** Prospective cohort study<br>**Prospective cohort:** Data collected prospectively. Database queried by research team. |
| **Participants** | **Included criteria:** HS athletes in 7 states (CA, FL, MN, NJ, OH, TN, TX).<br>**Excluded criteria:** Unwitnessed death. No details about the death<br>**Age:** High School age (14-19)<br>**Gender:** both<br>**Sport:** combination<br>**Level of Sport:** scholastic<br>**ECG Screened:** not described |
| **Interventions** | observational |
| **Outcomes** | **SCA:** yes<br>**SCD:** yes |

| | |
|---|---|
| | **Exertional:** all deaths<br>**Method of ID Cases:** web/media based searches-confirmed with interviews |
| **Identification** | **Sponsorship source:** National Center for Advancing Translational Sciences of the National Institutes of Health<br>**Country:** USA<br>**Setting:** High school athletes in Ohio, California, New Jersey, Florida, Minnesota, Tennessee, Texas<br>**Comments:** Query of prospective collection of SCA/D from Parent Heart Watch Database<br>**Authors name:** Kimberly Harmon, MD<br>**Institution:** University of Washington<br>**Email:**<br>**Address:** University of Washington3800 Montlake Blvd Seattle,WA 98195 |
| **Notes** | |

Risk of bias table

| **Bias** | **Authors' judgement** | **Support for judgement** |
|---|---|---|
| EPIDEMIOLOGY EXT VALIDITY: REPRESENTATIVE SAMPLE | Low risk | Judgement Comment: 7 large diverse states represented in the |
| EPIDEMIOLOGY INTERNAL VALIDITY: CASE DEFINITION | Low risk | Quote: "Sudden cardiac death was defined as a sudden unexpected death due to cardiac cause or a sudden death in a structurally normal heart with no other explanation for death and a history consistent with cardiac-related death."<br>Quote: "Sudden cardiac arrest was defined as an unexpected collapse (with or without physical exertion) due to a cardiac cause in which cardiopulmonary resuscitation was provided or an automated external defibril-lator (AED) deployed a shock in an individual who survived."<br>Quote: "in an individual who survived. Details of SCA/D events were investigated using online queries and information provided by coaches, athletic trainers, and parents to determine whether young athletes participated on a high school athletic team, which sports the athletes participated in, and in the activity during SCA/D (exertion and nonexertion) and whether the activity occurred during a school- |

| | | |
|---|---|---|
| | | sponsored event.</b> Cases involving nonathletes, those without"<br>Quote: "In cases of suspected SCD, autopsy reports were requested. If autopsy reports were not publicly available, attempts were made to con- tact the next of kin for permission. Those re- ports were reviewed and adjudicated by a panel with expertise in SCD in athletes and experience in autopsy review." |
| EPIDEMIOLOGY EXT VALIDITY: SAMPLING/SUBJECTS INCLUDED | Low risk | Judgement Comment: Selection of highly diverse, high population states allowed 35% of HS population to be included. |
| EPIDEMIOLOGY INTERNAL VALIDITY: STUDY PERIOD | Low risk | > 100,000 person years |
| EPIDEMIOLOGY INTERNAL VALIDITY: DATA COLLECTION SOURCE | High risk | Judgement Comment: Data collected via web reports, and when not good data available, by autopsy, and interview with eyewitnesses. |
| EPIDEMIOLOGY INTERNAL VALIDITY: NUMERATOR AND DENOMINATOR | High risk | Quote: "Unwitnessed deaths were not included as cardiac unless additional information such as autopsy results, negative toxicology screen results, or other information was available that could verify the death was cardiac in nature."<br>Quote: "Cases involving nonathletes, those without sufficient information, or those that were noncardiac in origin were excluded from this study."<br>Quote: "The NFHS has tracked participation statistics in high school sports since 1971 and tracks athletes participating in each sport; therefore, multisport athletes are counted in each sport in which they participate. Previous studies 5,8,11 have established a conversion factor to account for multisport athletes on the basis of data of unduplicated athletes participating in high school sports. The same conversion factor of 2.35 was applied in statistics looking at total incidence but not to sport-specific incidence in which exact numbers of athletes participating were available."<br>Quote: "These cases were identified using only media reports; therefore, this represents a minimum estimate. Previous studies have shown that media reports will detect only a proportion of SCD cases. In a Danish study 16 using death |

| | | |
|---|---|---|
| | | certificates as the primary means of identifying cases, media reports recognized only 20% of cases. In a study 1 of college athletes, media reports detected only 70% of SCDs. In that study, progressively fewer deaths were identified in lower profile"<br>Quote: "divisions, with 87% of SCDs identified in Division I, but only 61% in Division II and 44% in Division III; thus, it may be expected that high school athlete deaths would have even less media attention, resulting in only a fraction of cases being identified."<br>Judgement Comment: Unwitnessed deaths not included unless further records could be obtained, such as autopsy results. Authors clear about using media reports only, and clear about risk of undercount, citing other examples in discussion. |
| EPIDEMIOLOGY INTERNAL VALIDITY: MODE OF DATA COLLECTION | Low risk | Judgement Comment: All cases ID'd by internet surveillance method |
| EPIDEMIOLOGY FINAL SUMMARY: | Moderate risk | Judgement Comment: The summative statistic is likely an under-representation of SCA/D, but the authors are transparent about their methodology, and limitations, as well as the likelihood of the undercount due to their methods. |
| EPIDEMIOLOGY EXT VALIDITY: NONRESPONSE BIAS/FOLLOW UP OF COHORT | N/A | |

#### *Holst 2010*

| | |
|---|---|
| **Methods** | **Study design:** Retrospective cohort study<br>**Retrospective cohort:** Review of death certificates |

| | |
|---|---|
| **Participants** | **Included criteria:** All athletes dying during or within 1 hour of exertion, with data included on their death certificates<br>**Excluded criteria:** Death outside of 1 hour of exertion.<br>**Pretreatment:**<br>**Age:** 12-35<br>**Gender:** both<br>**Sport:** combination<br>**Level of Sport:** competitive<br>**ECG Screened:** no |
| **Interventions** | observational |
| **Outcomes** | **SCA:** no<br>**SCD:** yes<br>**Exertional:** yes, in the athletic population<br>**Method of ID Cases:** review of death certificates |
| **Identification** | **Sponsorship source:** Foundation of 17-12-1981, the John and Birthe Meyer Foundation, the Arvid Nilsson Foundation, the Danish National Research Foundation, the Danish Heart Foundation (07-10-R60-A1751-B743-22412), the Research Foundation at the Heart Centre, Rigshospitalet, the Research Foundation of Bispebjerg Hospital, and Bønnelykkefonden<br>**Country:** Denmark<br>**Setting:** Competitive athletes in all of Denmark<br>**Comments:**<br>**Authors name:** Dr. Anders G. Holst<br>**Institution:** Department of Cardiology, Copenhagen University Hospital<br>**Email:**<br>**Address:** Department of Cardiology, Section 2142Copenhagen University Hospital Rigshospitalet, Blegdamsvej 9, DK-2100 Copenhagen, Denmark |
| **Notes** | |

Risk of bias table

| Bias | Authors' judgement | Support for judgement |
|---|---|---|
| EPIDEMIOLOGY EXT VALIDITY: REPRESENTATIVE SAMPLE | Low risk | Judgement Comment: whole population of Denmark included |
| EPIDEMIOLOGY INTERNAL VALIDITY: CASE DEFINITION | Low risk | Quote: "We defined SCD in autopsied cases as the sudden, natural unexpected death of either unknown cause (sudden unexplained death) or cardiac cause (definite SCD); in unwitnessed cases as |

| | | |
|---|---|---|
| | | a person last seen alive and functioning normally fewer than 24 hours before being found, and in witnessed cases as an acute change in cardiovascular status with time to death being less than 1 hour."<br>Quote: "was not considered an SCD. SrSCD was defined as nontraumatic SCD occurring during or within 1 hour after moderate- to high-intensity exercise in a competitive athlete. The deceased was considered a competitive athlete if he or she did physically demanding sports and took part in competitions. We chose only to report SrSCD and not SCD among athletes in general because we could not be sure if information on whether the decedent was an athlete would be mentioned in the sources of information we had access to when death was not exercise-related. Conversely, the circumstances of the death were nearly always mentioned, allowing us to identify SrSCD. " |
| EPIDEMIOLOGY EXT VALIDITY: SAMPLING/SUBJECTS INCLUDED | Low risk | Judgement Comment: All death certificates evaluated in Denmark. |
| EPIDEMIOLOGY INTERNAL VALIDITY: STUDY PERIOD | Low risk | > 100,000 person years |
| EPIDEMIOLOGY INTERNAL VALIDITY: DATA COLLECTION SOURCE | Low risk | Quote: " All death certificates for deceased subjects ages 12 to 35 years dying in the period 2000 to 2006 in Denmark were retrieved as scanned computer files and reviewed independently by 2 physicians for possible SCD. The high level of information available on Danish death certificates allowed us to use these as a primary screening tool for the identification of possible SCD. |
| EPIDEMIOLOGY INTERNAL VALIDITY: NUMERATOR AND DENOMINATOR | Low risk | Quote: " All death certificates for deceased subjects ages 12 to 35 years dying in the period 2000 to 2006 in Denmark were retrieved as scanned computer files and reviewed independently by 2 physicians for possible SCD. The high level of information available on Danish death certificates allowed us to use these as a primary screening tool for the identification of possible SCD. "<br>Quote: "To validate our method of identifying SrSCD among athletes, an extensive retrospective media |

| | | |
|---|---|---|
| | | search was performed as a supplement to the main approach"<br>Quote: "media surveillance database Infomedia (www.infomedia. dk), a database of approximately 400 printed, 2,200 web- based, and the major radio and television Danish media, was used. All national and most regional media are included in the database."<br>Quote: "To estimate the size of the competitive athlete population (the at-risk population) in Denmark, we used data derived from the National Danish Health and Morbidity Study from the year 2005. This is an interview study of 3,848 people ages 16 to 35 years stratified on geographic as well as on socioeconomic parameters. 13 Because the age range in the study did not match ours (12 to 35 years), the assumption was made that we could extrapolate information about the percent of people participating in competition-level sports from the population ages 16 to 35 years to the population ages 12 to 35 years."<br>Judgement Comment: Some risk of incorrect denominator, but similar to other studies using Gov't estimates of athletes, etc. The authors did extend this to the 4 years under 16-which was not sampled in the government survey. Unclear if this would over/underestimate the denominator. Authors clear about their methods. |
| EPIDEMIOLOGY INTERNAL VALIDITY: MODE OF DATA COLLECTION | Low risk | Judgement Comment: All death certificates used for identification |
| EPIDEMIOLOGY FINAL SUMMARY: | Low risk | Judgement Comment: Authors clear about their method of data collection. They cross referenced their death certificate search with media searches to ensure collecting all available data. Only included exertional deaths. |
| EPIDEMIOLOGY EXT VALIDITY: NONRESPONSE BIAS/FOLLOW UP OF COHORT | N/A | |

#### *Jordaens 1996*

| | |
|---|---|
| **Methods** | **Study design:** Retrospective cohort study<br>**Retrospective cohort:** Review of newspapers for reports of cyclists deaths/cardiac events |
| **Participants** | **Included criteria:** Professional and semi-professional cyclists. No description of inclusion of cases, and how the cause of death was determined<br>**Excluded criteria:** none listed<br>**Age:** Unclear, appears to be under 35 based on comparison with control group of non-cyclists. All deaths in those under 26. Elected to include based on these data points<br>**Gender:** not described<br>**Sport:** cycling<br>**Level of Sport:** elite (professional/semi-professional)<br>**ECG Screened:** no |
| **Interventions** | observational |
| **Outcomes** | **SCA:** no<br>**SCD:** yes<br>**Exertional:** not described<br>**Method of ID Cases:** review of death certificates |
| **Identification** | **Sponsorship source:** none listed<br>**Country:** Belgium<br>**Setting:** Professional and semi professional cyclists in Belgium<br>**Comments:** Listed in an abstract in PACE from 1996. No contact details.<br>**Author's contact details**<br>**Name:** Luc Jordaens MD<br>**Institution:**<br>**Email:**<br>**Address:** |
| **Notes** | abstract |

Risk of bias table

| **Bias** | **Authors' judgement** | **Support for judgement** |
|---|---|---|
| EPIDEMIOLOGY EXT VALIDITY: REPRESENTATIVE SAMPLE | Unclear | Judgement Comment: Unclear-appears to include search for all of professional or semi-professional cyclists in Belgium-but poorly described |
| EPIDEMIOLOGY INTERNAL VALIDITY: CASE DEFINITION | Unclear | Not described in abstract |

| | | |
|---|---|---|
| EPIDEMIOLOGY EXT VALIDITY: SAMPLING/SUBJECTS INCLUDED | Unclear | Abstract, very little information provided. |
| EPIDEMIOLOGY INTERNAL VALIDITY: STUDY PERIOD | High risk | 15,722 person years |
| EPIDEMIOLOGY INTERNAL VALIDITY: DATA COLLECTION SOURCE | High risk | data collected from newspapers. |
| EPIDEMIOLOGY INTERNAL VALIDITY: NUMERATOR AND DENOMINATOR | Unclear | No description of the denominator. No description of ID of cases apart from news reports |
| EPIDEMIOLOGY INTERNAL VALIDITY: MODE OF DATA COLLECTION | Unclear | data collected from newspapers. Unclear how search was undertaken. |
| EPIDEMIOLOGY FINAL SUMMARY: | High Risk | Very little information in this abstract about how search was undertaken, and where the denominator comes from. Does not appear to be a corresponding full length article. |
| EPIDEMIOLOGY EXT VALIDITY: NONRESPONSE BIAS/FOLLOW UP OF COHORT | N/A | |

#### *Koskenvuo 1976*

| | |
|---|---|
| **Methods** | **Study design:** Retrospective cohort study<br>**Retrospective cohort:** Review of all sudden deaths in conscripts. |
| **Participants** | **Included criteria:** Finnish conscripts between 1948-1972. Cases were included with or without exertion; these included cases of sudden death only, not only cardiac cases.<br>**Excluded criteria:** none listed<br>**Age:** About 18-24-not clearly stated.<br>**Gender:** male<br>**Sport:** military<br>**Level of Sport:** conscripts<br>**ECG Screened:no** |
| **Interventions** | observational |
| **Outcomes** | **SCA:** no<br>**SCD:** yes |

| | |
|---|---|
| | **Exertional:** not described<br>**Method of ID Cases:** combination of autopsy and military records |
| **Identification** | **Sponsorship source:** none<br>**Country:** Finland<br>**Setting:** Finnish male conscripts to the Finnish armed forces<br>**Comments:**<br>**Authors name:** Mimo Koskenvuo<br>**Institution:** Medical Section General Headquarters Defence Forces,<br>**Email:**<br>**Address:** Medical SectionGeneralHeadquarters Defence Forces 00101 Helsinki 10, Finland |
| **Notes** | |

Risk of bias table

| **Bias** | **Authors' judgement** | **Support for judgement** |
|---|---|---|
| EPIDEMIOLOGY EXT VALIDITY: REPRESENTATIVE SAMPLE | Low risk | Judgement Comment: male military conscripts in Finland. |
| EPIDEMIOLOGY INTERNAL VALIDITY: CASE DEFINITION | Unclear risk | Judgement Comment: Case definition not described. The authors appear to use the autopsy data, and military records as their form of ID. It is not clear how they authors came to the final decision about what type of death they had, except to say that records, and necropsy data was reviewed. |
| EPIDEMIOLOGY EXT VALIDITY: SAMPLING/SUBJECTS INCLUDED | Low risk | Quote: "The study population consisted of nearly 900 000 young men aged about 20 years who were Finnish conscripts in 1948-72. Nearly 660 000 man-years were covered." |
| EPIDEMIOLOGY INTERNAL VALIDITY: STUDY PERIOD | Low risk | > 100,000 person years |
| EPIDEMIOLOGY INTERNAL VALIDITY: DATA COLLECTION SOURCE | Low risk | Judgement Comment: Military autopsy and other military record data regarding the death. |
| EPIDEMIOLOGY INTERNAL VALIDITY: NUMERATOR AND DENOMINATOR | Unclear risk | Quote: "The health data on conscripts who died suddenly, the circumstances and clinical course of the acute illness, and the necropsy findings were analysed. The residence, social class, findings at initial medical examination, and use of medical |

| | | |
|---|---|---|
| | | services during military service were compared in the conscripts who died and their controls." Judgement Comment: Risk of bias here may be in the categorization of death. Different understanding currently about exertional or cardiac death in young active people compared to 40 years ago. The ID, and the denominator appear valid for the time. |
| EPIDEMIOLOGY INTERNAL VALIDITY: MODE OF DATA COLLECTION | Low risk | Judgement Comment: The same for all cases. |
| EPIDEMIOLOGY FINAL SUMMARY: | Moderate risk | Judgement Comment: The authors describe their methods, but leave out exactly how the categorized the deaths. The data source appears appropriate, but how they cases or the files were searched is not clear. Overall, the use of military records is reassuring, and likely presents a lower risk of bias- but without description of above, will consider unclear. |
| EPIDEMIOLOGY EXT VALIDITY: NONRESPONSE BIAS/FOLLOW UP OF COHORT | N/A | |

#### *Kucera 2018*

| | |
|---|---|
| **Methods** | **Study design:** Prospective cohort study<br>**Prospective cohort:** Open cohort, with cases identified by voluntary report only |
| **Participants** | **Included criteria:** Football players death related to football. We included both exertional and non-exertional deaths in active football players.<br>**Excluded criteria:** none listed<br>**Age:** High school and older-including college (14-24)<br>**Gender:** male<br>**Sport:** American Football<br>**Level of Sport:** combination (scholastic and university)<br>**ECG Screened:** no |

| | |
|---|---|
| **Interventions** | observational |
| **Outcomes** | **SCA:** no<br>**SCD:** yes<br>**Exertional:** no, all deaths<br>**Method of ID Cases:** report to national center for catastrophic sport injury; combination of witness interviews and records if available |
| **Identification** | **Sponsorship source:** The National Center for Catastrophic Sport Injury Research is funded by the American Football Coaches Association, the National Collegiate Athletic Association, National Federation of State High School Associations, National Athletic Trainers' Association, the American Medical Society for Sports Medicine, the National Operating Committee on Standards for Athletic Equipment, and The University of North Carolina at Chapel Hill.<br>**Country:** USA<br>**Setting:** All football players in USA<br>**Comments:** Included data only regarding High school and above<br>**Authors name:** Kristen L. Kucera, MSPH, PhD, ATC<br>**Institution:** National Center for Catastrophic Sport Injury Research The University of North Carolina at Chapel Hill<br>**Email:**<br>**Address:** |
| **Notes** | |

Risk of bias table

| **Bias** | **Authors' judgement** | **Support for judgement** |
|---|---|---|
| EPIDEMIOLOGY EXT VALIDITY: REPRESENTATIVE SAMPLE | Low risk | Judgement Comment: all American football players in the US eligible for this study |
| EPIDEMIOLOGY INTERNAL VALIDITY: CASE DEFINITION | High risk | Judgement Comment: Cases IDd after report, and then cause of death assessment is made via interviews, and possibly autopsy reports. It is not clear in the article who makes final decision or how. |
| EPIDEMIOLOGY EXT VALIDITY: SAMPLING/SUBJECTS INCLUDED | High risk | Judgement Comment: For same reason as numerator and denominator. The ID of cases occurred voluntarily, and so not clear that all are included. |
| EPIDEMIOLOGY INTERNAL VALIDITY: STUDY PERIOD | Low risk | > 100,000 person years |

| | | |
|---|---|---|
| EPIDEMIOLOGY INTERNAL VALIDITY: DATA COLLECTION SOURCE | High risk | Judgement Comment: As above, data on cases comes from different sources. |
| EPIDEMIOLOGY INTERNAL VALIDITY: NUMERATOR AND DENOMINATOR | High risk | Quote: "Survey 2017 3 Data Collection : Data were compiled with the assistance of coaches, athletic trainers, athletic directors, executive officers of state and national athletic organizations, online news reports, online reports, and professional associates of the researchers. NCCSIR and the Consortium for Catastrophic Injury Monitoring in Sport have developed an online portal where anyone can report a catastrophic event (https:\\www.sportinjuryreport.org). Throughout the year (January 1 to December 31), upon notification of a suspected football fatality, contact by telephone, email, or personal letter questionnaire was made with the appropriate individuals including state high school association official, school or team administrator, coach, athletic trainer, team physician, and/or the family. Individuals are asked to complete a brief survey about the event at https:\\www.sportinjuryreport.org. Autopsy reports are used when available. All activities are approved by the Institutional Review Board (IRB) of the University of North Carolina at Chapel Hill (IRB# 05-0018). Participation in Football Reports prior"<br>Judgement Comment: Cases are included if voluntarily reported to National center for catastrophic sport injury research. Would seemingly undercount the deaths significantly. Denominator from sporting records. |
| EPIDEMIOLOGY INTERNAL VALIDITY: MODE OF DATA COLLECTION | High risk | Judgement Comment: Some cases have autopsy data, others do not. |
| EPIDEMIOLOGY FINAL SUMMARY: | High risk | Judgement Comment: The risk associated with undercount of cases in this data set is very high. Report of cases relied on voluntary contact to the agency collecting the data. |
| EPIDEMIOLOGY EXT VALIDITY: NONRESPONSE | N/A | |

| |
|---|
| BIAS/FOLLOW UP OF COHORT |

#### *Landry 2017*

| | |
|---|---|
| **Methods** | **Study design:** Retrospective cohort study<br>**Retrospective cohort:** Review of prospectively collected RescuEpistry database of out of hospital cardiac arrests in 6.6 million catchment area in Ontario. |
| **Participants** | **Included criteria:** Athletes competitive or non competitive 12-45 years old suffering out of hospital cardiac arrest within 1 hour of playing sport. We collected only the data on 12-34 year old competitive athletes, and did not extract the data on 35+, or non-competitive athletes.<br>**Excluded criteria:** none listed<br>**Age:** 12-34<br>**Gender:** both<br>**Sport:** combination<br>**Level of Sport:** competitive<br>**ECG Screened:** no |
| **Interventions** | Observational |
| **Outcomes** | **SCA:** yes<br>**SCD:** yes<br>**Exertional:** yes<br>**Method of ID Cases:** combination of medical and autopsy records |
| **Identification** | **Sponsorship source:** The study was supported by the National Heart, Lung, and Blood Institute, the Canadian Institutes of Health Research, and others<br>**Country:** Canada<br>**Setting:** Ontario region with 6.6 million inhabitants<br>**Comments:** Prospectively collected database was retrospectively reviewed. Data base called Rescue Epistry cardiac arrest database is a prospective, population-based registry of consecutive out-of-hospital cardiac arrests attended by EMS personnel who were responding to 911 calls in a specific area of Ontario<br>**Authors name:** Cameron H. Landry MD, Paul Dorian MD (Last)<br>**Institution:** University of Toronto<br>**Email:**<br>**Address:** Division of Cardiology University of Toronto St. Michael's Hospital Toronto, ON M5B 1W8, Canada |

| Notes |
|---|

Risk of bias table

| Bias | Authors' judgement | Support for judgement |
|---|---|---|
| EPIDEMIOLOGY EXT VALIDITY: REPRESENTATIVE SAMPLE | Low risk | Judgement Comment: Whole population in catchment area. Sports related deaths eligible for study. |
| EPIDEMIOLOGY INTERNAL VALIDITY: CASE DEFINITION | Low risk | Quote: "Cases of out-of-hospital cardiac arrest that were related to competitive or noncompetitive sports were defined as cases that occurred during, or within 1 hour after, exertion of more than 3 METs during the activity."<br>Judgement Comment: Exertional deaths only included. |
| EPIDEMIOLOGY EXT VALIDITY: SAMPLING/SUBJECTS INCLUDED | Low risk | Judgement Comment: All subjects dying of SCD in the catchment area in Ontario appear to be included. |
| EPIDEMIOLOGY INTERNAL VALIDITY: STUDY PERIOD | Low risk | > 100,000 person years |
| EPIDEMIOLOGY INTERNAL VALIDITY: DATA COLLECTION SOURCE | Low risk | Judgement Comment: Hospital/EMS/Coroner records. |
| EPIDEMIOLOGY INTERNAL VALIDITY: NUMERATOR AND DENOMINATOR | Low risk | Quote: "The estimated total number of competitive athletes in the region served by the participating EMS agencies (Fig. S1 in the Supplementary Appendix) who were 12 to 45 years of age was calculated on the basis of the total number of competitive athletes who had registered with a sporting organization in Ontario during 2012 (information was obtained through direct correspondence with the Ontario Ministry of Tourism, Culture, and Sport) and was prorated ac- cording to the age-matched population in the geographic area covered by the study with the use of the 2011 Canadian Census. Athletes who were registered in racing events were recorded separately from those who participated in other sports, according to region and age group. 20 It was assumed that the number of athletes did not vary significantly from year to year within the study period. 21" |

| | | |
|---|---|---|
| | | Quote: "3 METs during the activity. We identified such cases by manually sorting through all ambulance call reports and records from the emergency department or hospital for reports of persons who had a cardiac arrest at a recreational facility, university or college, sports field, stadium or arena, athletic facility, golf course, water area, hotel, condominium or apartment, park, or street. Cases were cross-referenced by comparison with additional data sources to obtain a clinical and pathological assessment that was as complete as possible. The data sources that were used included ambulance call reports, fire call reports, in-hospital data (abstracted from emergency department reports, in-hospital medical notes, discharge summaries, consultations, clinical tests, and medical certificates of death)<br>Quote: " medical records from family physicians, coroner investigative statements, autopsy reports, toxicology reports, and records of direct interviews with patients or family members. All out-of-hospital cardiac arrests were classified as either sudden cardiac arrest or cardiac arrest from other causes, as defined above and described previously. Autopsy and Molecular Autopsy Autopsies"<br>Quote: "and the end of 2014. There were an estimated 352,499 registered competitive athletes in the study region in 2012 (which represented 11.4% of the population in the study region), resulting in an estimated total follow-up time of 2.1 mil- lion athlete-years. "<br>Judgement Comment: Authors very clear about their ID of cases, and that the population under observation should be all included, and that cases of SCA should have been captured by their data system. The only question comes from the 'registration' of competitive athletes, and whether all competitive athletes were included. |
| EPIDEMIOLOGY INTERNAL VALIDITY: MODE OF DATA COLLECTION | Low risk | Judgement Comment: EMS run sheets, hospital records, coroner records. |
| EPIDEMIOLOGY FINAL SUMMARY: | Low risk | Judgement Comment: The authors appear to have been very thorough; with an open cohort such as |

| | | |
|---|---|---|
| | | this, they appear to have collected as much data as possible from a prospectively collected data base. |
| EPIDEMIOLOGY EXT VALIDITY: NONRESPONSE BIAS/FOLLOW UP OF COHORT | N/A | |

#### *Malhotra 2018*

| | |
|---|---|
| **Methods** | **Study design:** Prospective cohort study<br>**Prospective cohort:** Not described by authors, but prospectively collected data and follow up of adolescent soccer players at elite level; with multi-source follow up of the cases of SCD. |
| **Participants** | **Included criteria:** Adolescent soccer players being offered professional contracts at the ages of 15-17. All deaths were investigated, and categorized (exertional and non-exertional).<br>**Age:** entered cohort @ age 15-17<br>**Gender:** male<br>**Sport:** soccer<br>**Level of Sport:** Elite (professional)<br>**ECG Screened:** yes |
| **Interventions** | Observational |
| **Outcomes** | **SCA:** no<br>**SCD:** yes<br>**Exertional:** no, all deaths<br>**Method of ID Cases:** combination of medical/autopsy records and media reports |
| **Identification** | **Sponsorship source:** English Football Association, Cardiac Risk in the Young, Charles Wolfson charitable trust<br>**Country:** England<br>**Setting:** Elite adolescent soccer players being offered professional contracts age 15-17<br>**Comments:**<br>**Authors name:** Aneil Malhotra, MB, BChir, PhD; Sanjay Sharma MB, ChB, MD (last) |

| | |
|---|---|
| | **Institution:** Cardiology Clinical Academic Group, St. George's, University of London<br>**Email:**.u<br>**Address:** Cardiology Clinical Academic Group, St. George's, University of London Cranmer Terrace, London SW17 0REUnited Kingdom |
| **Notes** | 2 of the 8 cases of death were identified with abnormalities and advised to stop playing; but continued. |

Risk of bias table

| **Bias** | **Authors' judgement** | **Support for judgement** |
|---|---|---|
| EPIDEMIOLOGY EXT VALIDITY: REPRESENTATIVE SAMPLE | Low risk | Judgement Comment: All youth soccer players turning professional were screened and included in the cohort. 95% male. All entered cohort between 15-17 years of age. |
| EPIDEMIOLOGY INTERNAL VALIDITY: CASE DEFINITION | Low risk | Quote: "Death certificates were obtained from the U.K. government for all deceased persons in the cohort to ascertain the causes of death, which were categorized broadly as accidental, suicide, drug-related, cancer, or cardiac causes. Autopsy data were available in all cases of sudden cardiac death, and diagnoses were based on previously established pathological criteria in conjunction with consultation with an expert cardiac pathologist." |
| EPIDEMIOLOGY EXT VALIDITY: SAMPLING/SUBJECTS INCLUDED | Low risk | Judgement Comment: all youth soccer players being offered professional contracts within the FA around 15-17 years of age. 95% male. |
| EPIDEMIOLOGY INTERNAL VALIDITY: STUDY PERIOD | Low risk | Judgement Comment: 118,000+ years of follow up |
| EPIDEMIOLOGY INTERNAL VALIDITY: DATA COLLECTION SOURCE | Low risk | Quote: "among athletes were ascertained through the development of a database that was compiled from voluntary reports to the FA. A second method to ascertain the number of deaths was through a secure survey that was sent to health professionals at each of the 92 FA-affiliated clubs, asking specifically about deaths from any cause. In addition, regular Internet searches had been performed since 2005, with the use of three different search engines (Google, Yahoo, and MSN search [Bing]) and at least 16 keywords (student, athlete, col- lapsed, died, death, heart, cardiac, |

| | | |
|---|---|---|
| | | arrest, attack, soccer, running, school, unknown, college, defibrillator, and saved). Death certificates were obtained from"<br>Judgement Comment: Deaths from voluntary reports from clubs, death certificates, internet searches, and a survey of clubs. |
| EPIDEMIOLOGY INTERNAL VALIDITY: NUMERATOR AND DENOMINATOR | Low risk | Quote: "The program encompassed all youth academy players (15 to 17 years of age) across the 92 professional clubs in the soccer league system who had excelled within the preceding 5 years."<br>Quote: "Calculation of the follow-up period per athlete was based on the number of years of competition within the FA, which was determined from the FA registry of players."<br>Judgement Comment: Deaths ascertained from multi source follow up, including club report, internet search, death certificates, and survey of clubs. |
| EPIDEMIOLOGY INTERNAL VALIDITY: MODE OF DATA COLLECTION | Low risk | Judgement Comment: The same multi-source data collection method was undergone to identify all deaths. |
| EPIDEMIOLOGY FINAL SUMMARY: | Low risk | Judgement Comment: This study and paper were designed more to report the results of screening program with elite adolescent soccer players. A portion of the paper reported the SCD in this cohort, and details a thorough follow up of those included in this known cohort. Both the cases, and the denominator represent what I believe to be reliable numbers. |
| EPIDEMIOLOGY EXT VALIDITY: NONRESPONSE BIAS/FOLLOW UP OF COHORT | N/A | |

#### *Marijon 2011*

| | |
|---|---|
| **Methods** | **Study design:** Retrospective cohort study<br>**Prospective cohort:** Data collected from EMS database, as well as web report prospectively over 6 year period |

| | |
|---|---|
| **Participants** | **Included criteria:** all those suffering sport related sudden death ages 10-75. Subgroup offered of 10-35 competitive athletes, which we have extracted. There is a corresponding article published which focuses on sex differences between the men and women. This article does not give the number of deaths, nor the number of person years for men or women under age 35, only the full 10-75 age group.<br>**Excluded criteria:** none listed<br>**Pretreatment:**<br>**Age:** 10-35 competitive athletes<br>**Gender:** both<br>**Sport:** combination<br>**Level of Sport:** competitive<br>**ECG Screened:** no |
| **Interventions** | Observational |
| **Outcomes** | **SCA:** no<br>**SCD:** yes<br>**Exertional:** yes<br>**Method of ID Cases:** combination of media reports and autopsy/medical records |
| **Identification** | **Sponsorship source:** INSERM (Institut National de la Santé et de la Recherche Médicale) and IRMES (Institut de Recher-che BioMédicale et d'Épidemiologie du Sport)<br>**Country:** France<br>**Setting:** 60 of 96 districts in France<br>**Comments:**<br>**Authors name:** Eloi Marijon, MD<br>**Institution:** Université Paris Descartes, Assistance Publique Hospitaux de Paris, Hospital Européen Georges Pompidou, Departement de Cardiologie, Paris, Franc<br>**Email:**<br>**Address:** Université Paris, Descartes,UMR-S970, Paris, France |
| **Notes** | |

Risk of bias table

| **Bias** | **Authors' judgement** | **Support for judgement** |
|---|---|---|
| EPIDEMIOLOGY EXT VALIDITY: REPRESENTATIVE SAMPLE | Low risk | Judgement Comment: all population in participating districts considered, no limitations. Cases only considered in 'competitive' athletes, excluding intramural sport while in colleges. |
| EPIDEMIOLOGY INTERNAL VALIDITY: CASE DEFINITION | Low risk | Quote: "death was considered to be sudden if it occurred within 1 hour of symptom onset. 7,8,15 |

| | | |
|---|---|---|
| | | Sports-related SD was defined if death occurred during sport or within 1 hour of cessation of sports activity."<br>Quote: "Sports-related SDs that occurred in competitive athletes outside the context of physical activity were not included. Sports participants who survived cardiac arrest after defibrillation and/or cardiopulmonary resuscitation (CPR) were considered to have experienced sports-related SD for the purpose of the present analysis."<br>Judgement Comment: No deaths outside of sports participation-Exertional only, and SCA included in the SCD count. |
| EPIDEMIOLOGY EXT VALIDITY: SAMPLING/SUBJECTS INCLUDED | Low risk | Quote: "“young competitive athlete” was defined as any person 10 to 35 years old who participated in an organized sports program (team or individual sport) that required regular competition and training according to the National Registry from the Ministere de la Sante ´ et des Sports (individuals participating in college-sponsored intramural sports were not considered young competitive athletes). 16" |
| EPIDEMIOLOGY INTERNAL VALIDITY: STUDY PERIOD | Low risk | Judgement Comment: 5 year, large population observed, greater than 1 million athlete years |
| EPIDEMIOLOGY INTERNAL VALIDITY: DATA COLLECTION SOURCE | Low risk | Judgement Comment: Collected data from French SAMU (EMS) system, as well as cross checked with web based monitoring system. Then events committee reviewed the deaths and adjudicated the likelihood of SD. |
| EPIDEMIOLOGY INTERNAL VALIDITY: NUMERATOR AND DENOMINATOR | Low risk | Quote: "purpose of the present analysis. To maximize case detection, 2 complementary independent methds were instituted that have already been used by other groups for young competitive athletes. 8 We tested the efficacy of the following case-detection methodology in France by performing a preliminary 6-month pilot study over 5 districts in 2004. First, a prospective case reporting system was instituted via the mobile intensive care services (SAMU); the French SAMU system is a nationwide emergency medical service accessed via a well-known nationwide free call number. Thus, SAMU is systematically called in cases of cardiac |

| | | |
|---|---|---|
| | | arrest or sudden collapse. Emergency health technicians in SAMU assess these subjects and start resuscitation where required, on site. For each case of sports-related SD during the study period, a specific detailed report form was completed by SAMU and then transmitted to the data collection center (INSERM U970, Paris, France) within the following 3 months. In parallel, a media search program screened for cases from continuous Web-based monitoring via Google Reader using the Really Simple Syndication system for local, regional, and national newspapers (n 275) with the key words “death,” “cardiac arrest,” and “dizziness.” Manual checking of these reports was performed to identify verifiable sports-related SDs in 10- to 75-year- olds, and these cases were followed up if we had not already been notified of them via SAMU; complementary information was then obtained from written accounts or telephone interviews with the local emergency team of the SAMU. A systematic review of all sports-related SD notifications was undertaken by an independent events committee every 6 months. Three experts in sports-related SD adjudicated cases to exclude SDs unrelated to sports, nonfatal sports-related cardiovascular events, and deaths that did not meet the SD definition, and finally to identify incomplete files that required further information. "<br>Quote: "The specific incidence of sports-related SD among young competitive athletes was calculated according to data from the Ministere de la Sante ´ et des Sports. 16 The participating number of 10- to 35-year-old athletes over the study period was 1 015 293, and this figure was considered stable over time. 16 Because of the constant high level of media coverage of sports-related SDs in young sports participants in France (that is, such reports were considered likely to be comprehensive), we provided only 1 incidence rate calculation for this population."<br>Judgement Comment: Authors clear about methodology. Did a trial run of their collection of numerator method. Separate out the 'young |

| | | |
|---|---|---|
| | | competitive' athlete number and methodology for denominator and numerator. May have missed some deaths as only included exertional deaths during, or directly after sport. Do not include incidents which may be later in day after exercise, or sleep as an example |
| EPIDEMIOLOGY INTERNAL VALIDITY: MODE OF DATA COLLECTION | Low risk | Judgement Comment: clearly explained. |
| EPIDEMIOLOGY FINAL SUMMARY: | Low risk | Judgement Comment: Authors very clear about methods. 2 tiered system for identifying deaths; results of deaths adjudicated by panel as SCD or not. Risks are of including SCA and SCD together as SCD. Young competitive athletes not the primary outcome, but included as a subgroup. |
| EPIDEMIOLOGY EXT VALIDITY: NONRESPONSE BIAS/FOLLOW UP OF COHORT | N/A | |

#### *Maron 1998*

| | |
|---|---|
| **Methods** | **Study design:** Retrospective cohort study<br>**Retrospective cohort:** Review of catastrophic insurance records for deaths occurring while participating in HS sports activity, which is mandatory for Minnesota HS athletes. |
| **Participants** | **Included criteria:** Includes all sudden cardiac deaths in high school athletes grades 10-12.<br>**Excluded criteria:** none listed<br>**Age:** 13-19<br>**Gender:** both<br>**Sport:** combination<br>**Level of Sport:** scholastic<br>**ECG Screened:** no |
| **Interventions** | Observational |
| **Outcomes** | **SCA:** no<br>**SCD:** yes<br>**Exertional:** not detailed, assume yes as included events only occurred during practice/games |

| | |
|---|---|
| | **Method of ID Cases:** autopsy reports/insurance records |
| **Identification** | **Sponsorship source:** none listed<br>**Country:** USA<br>**Setting:** Minnesota High School athletes grade 10-12<br>**Authors name:** Barry Maron, MD<br>**Institution:** Cardiovascular Research Division, Minneapolis Heart Institute Foundation;<br>**Email:**<br>**Address:** Minneapolis Heart Institute Foundation, 920 E. 28th Street, Suite 40, Minneapolis, Minnesota55407 |
| **Notes** | Does not differentiate exertion vs. non-exertion, but would follow that if the insurance is against catastrophic event playing high school sport, that it is exertional, and is only used when death occurs during sports activity. |

Risk of bias table

| Bias | Authors' judgement | Support for judgement |
|---|---|---|
| EPIDEMIOLOGY EXT VALIDITY: REPRESENTATIVE SAMPLE | Low risk | Judgement Comment: All high school athletes accounted for in grades 10-12. |
| EPIDEMIOLOGY INTERNAL VALIDITY: CASE DEFINITION | Low risk | Judgement Comment: deaths confirmed by autopsy, and by insurance records. These would not include deaths due to CV issues outside of HS sports activity |
| EPIDEMIOLOGY EXT VALIDITY: SAMPLING/SUBJECTS INCLUDED | Low risk | Judgement Comment: All high school athletes appear to be accounted for, there was a multiplier used to estimate athletes/year. All athletes dying in competition or with something to do with HS sports appear to be accounted for. |
| EPIDEMIOLOGY INTERNAL VALIDITY: STUDY PERIOD | Low risk | > 100,000 person years |
| EPIDEMIOLOGY INTERNAL VALIDITY: DATA COLLECTION SOURCE | Low risk | Judgement Comment: Collected from autopsy and catastrophic insurance records. Possible that the deaths were undercounted, but it would seem that deaths during HS sports activity were counted appropriately. |
| EPIDEMIOLOGY INTERNAL VALIDITY: NUMERATOR AND DENOMINATOR | High risk | Quote: "This study population was selected primarily because of the long-standing, mandatory insurance plan covering catastrophic injury or death that is provided by the league and routinely administered to all student athletes engaged in interscholastic |

| | | |
|---|---|---|
| | | sports programs at the varsity and junior varsity levels within the state. The records of this indemnity program permitted an accurate assessment of the number of participants in high school sports, as well as the number of deaths during this period of time. The following data relevant to the occurrence of sudden death in competitive interscholastic athletes were obtained from MSHSL:"<br>Quote: " The average number of sports participated in by individual athletes over an academic year, calculated for 1996 –1997, and used to indirectly estimate the number of individual student athlete participants from the number of sports participations for the overall study period. Results Sports participations. "<br>Judgement Comment: Numerator misses students dying outside of sports (as much as 20% in some studies); also would miss student athletes dying outside of HS sports participation, such as summer league and club activity. |
| EPIDEMIOLOGY INTERNAL VALIDITY: MODE OF DATA COLLECTION | Low risk | Judgement Comment: Same data used for all students/cases. |
| EPIDEMIOLOGY FINAL SUMMARY: | Moderate risk | Judgement Comment: There may be an undercount of overall number of athletes of high school age dying due to sports activity as it leaves out the time out of official sport activity (or when out of season and just exercising/working out/training). But authors clear about their methods and appear to account for high school athletes in season dying exertional deaths well. The final numbers almost certainly represent an undercount of the incidence- but this article is clear about its goal, and its findings. |
| EPIDEMIOLOGY EXT VALIDITY: NONRESPONSE BIAS/FOLLOW UP OF COHORT | N/A | |

#### *Maron 2009*

| | |
|---|---|
| **Methods** | **Study design:** Retrospective cohort study<br>**Retrospective cohort:** Authors do not describe whether this was prospective or retrospective, data appear to have been collected prospectively, but reviewed at a later day. |
| **Participants** | **Included criteria:** Lacrosse players in High school and college deaths in these players at any time, there does not appear to be an exertional component to this study.<br>**Age:** 14-23 (all deaths in males)<br>**Gender:** both<br>**Sport:** lacrosse<br>**Level of Sport:** combination (scholastic and university)<br>**ECG Screened:** no |
| **Interventions** | observational |
| **Outcomes** | **SCA:** yes<br>**SCD:** yes<br>**Exertional:** no, all deaths<br>**Method of ID Cases:** combination of medical/autopsy records, media reports |
| **Identification** | **Sponsorship source:** Hearst Foundations ( San Francisco, CA), Medtronic Physio-Control Corp, (Redmond, WA), Philips Medical Systems, Inc (Andover, MA),and National Operating Committee for Standards in Athletic Equipment(Chapel Hill, NC)H<br>**Country:** USA<br>**Setting:** US high school and college lacrosse players<br>**Comments:**<br>**Authors name:** Barry Maron, MD<br>**Institution:** Hypertrophic Cardiomyopathy Center, Minneapolis Heart Institute Foundation, Minneapolis, Minnesota;<br>**Email:**<br>**Address:** Barry J. Maron, MD, Minneapolis Heart Institute Foundation, 920 E 28th St, Suite 620, Minneapolis, MN 5540 |
| **Notes** | |

Risk of bias table

| **Bias** | **Authors' judgement** | **Support for judgement** |
|---|---|---|
| EPIDEMIOLOGY EXT VALIDITY: REPRESENTATIVE SAMPLE | Low risk | Judgement Comment: High school and college lacrosse players. The authors set out to study this group, and do so. |

| | | |
|---|---|---|
| EPIDEMIOLOGY INTERNAL VALIDITY: CASE DEFINITION | Low risk | Judgement Comment: Eyewitness accounts, web based reports, autopsy reports where available. The case definitions appear well explained. |
| EPIDEMIOLOGY EXT VALIDITY: SAMPLING/SUBJECTS INCLUDED | High risk | Judgement Comment: All US lacrosse players in HS and college accounted for in denominator. It appears the cases were accounted for in the same way, primarily by news and web reports. The authors note the count of cases of death increases in the later years of the study, this is possibly related to more lacrosse players, but also may suggest greater access to information with web based search engines vs. older news searches such as LexisNexis All US lacrosse players in HS and college accounted for in denominator. It appears the cases were accounted for in the same way, primarily by news and web reports. The authors note the count of cases of death increases in the later years of the study, this is possibly related to more lacrosse players, but also may suggest greater access to information with web based search engines vs. older news searches such as LexisNexis |
| EPIDEMIOLOGY INTERNAL VALIDITY: STUDY PERIOD | Low risk | > 100,000 person years |
| EPIDEMIOLOGY INTERNAL VALIDITY: DATA COLLECTION SOURCE | Low risk | Quote: "well as US Lacrosse (www.uslacrosse.org). A systematic tracking process was established to assemble detailed in- formation on each case subject, including the autopsy report (with gross anatomic, histologic, and toxicological findings), and pertinent clinical and demographic information. Selected data (eg, circumstances of collapse) were often derived from written ac- counts or telephone interviews when in contact with family members, wit- nesses, or coaches. When necessary, autopsy findings were verified by direct communication with the medical examiner. This project was reviewed by the" Judgement Comment: Once identified as a death/arrest, the data collection sources appear to be appropriate. |
| EPIDEMIOLOGY INTERNAL VALIDITY: NUMERATOR AND DENOMINATOR | High risk | Quote: "The study population was identified by targeted searches using a variety of sources, as reported previousy, ie assembled most prominently by accessing the public record with the LexisNexis |

| | | |
|---|---|---|
| | | archival informational database and news media accounts, as well as US Lacrosse (www.uslacrosse.org)."<br>Quote: "the numbers of participants in sanctioned high school and intercollegiate varsity lacrosse, as well as in 9 other competitive sports, were obtained from the National Federation of High Schools 10 and the National Collegiate Athletic Association. 11 Deaths per 100 000 person- years of participation were calculated by dividing the total number of deaths by the number of participations for all of the years (and multiplying by 100 000)."<br>Judgement Comment: High risk of bias if only ID'd via news and web reports. The cases were verified with autopsy if possible |
| EPIDEMIOLOGY INTERNAL VALIDITY: MODE OF DATA COLLECTION | High risk | Judgement Comment: Information collection appears to be through the same avenues for all cases, as well as the denominator. The cases themselves are collected from multiple sources, primarily news/web reports, as well as US lacrosse. High risk of bias and undercount with collection from news reports |
| EPIDEMIOLOGY FINAL SUMMARY: | High risk | Judgement Comment: Authors clear about their study, how cases and those at risk were identified. The authors do not address the issue with web based/news report identification of cases. This is likely an undercount of the true cases because of the method of identification, but all cases treated the same. While the methodology is clear, the risk of undercount is real, and affects the assessment of the study. |
| EPIDEMIOLOGY EXT VALIDITY: NONRESPONSE BIAS/FOLLOW UP OF COHORT | N/A | |

#### *Maron 2013*

| | |
|---|---|
| **Methods** | **Study design:** Retrospective cohort study |

| | |
|---|---|
| | **Retrospective cohort:** Appears to be a retrospective analysis of data collected prospectively. Data from insurance registry likely makes the more of a retrospective analysis |
| **Participants** | **Included criteria:** Unclear for deaths. Abstract reports deaths occurring in organized sport activity (which was true for all deaths); the methodology and discussion indicates deaths at any time in athletes was considered. All MN scholastic athletes included.<br>**Excluded criteria:** none listed<br>**Age:** 12-19<br>**Gender:** both<br>**Sport:** combination<br>**Level of Sport:** scholastic<br>**ECG Screened:** no |
| **Interventions** | Observational |
| **Outcomes** | **SCA:** no<br>**SCD:** yes<br>**Exertional:** no, all deaths<br>**Method of ID Cases:** combination medical/autopsy reports, web/media reports included in US Registry Sudden Death in Athletes |
| **Identification** | **Sponsorship source:** None listed<br>**Country:** USA<br>**Setting:** Minnesota Scholastic Athletes<br>**Comments:**<br>**Authors name:** Barry Maron, MD<br>**Institution:** Minneapolis Heart Institute Foundation<br>**Email:**<br>**Address:** Hypertrophic Cardiomyopathy Center Minneapolis Heart Institute Foundation920 East 28th St, Suite 620 Minneapolis, MN 55407 |
| **Notes** | |

Risk of bias table

| **Bias** | **Authors' judgement** | **Support for judgement** |
|---|---|---|
| EPIDEMIOLOGY EXT VALIDITY: REPRESENTATIVE SAMPLE | Low risk | Judgement Comment: appears to include all scholastic athletes in MN over 26 year period. |
| EPIDEMIOLOGY INTERNAL VALIDITY: CASE DEFINITION | Unclear risk | Judgement Comment: Case definition not described in this article. |
| EPIDEMIOLOGY EXT VALIDITY: | Low risk | Judgement Comment: All scholastic athletes included. |

| | | |
|---|---|---|
| SAMPLING/SUBJECTS INCLUDED | | |
| EPIDEMIOLOGY INTERNAL VALIDITY: STUDY PERIOD | Low risk | > 100,000 person years |
| EPIDEMIOLOGY INTERNAL VALIDITY: DATA COLLECTION SOURCE | Low risk | Judgement Comment: 12 of 13 deaths had autopsy. |
| EPIDEMIOLOGY INTERNAL VALIDITY: NUMERATOR AND DENOMINATOR | High risk | Quote: "METHODS The forensic case records of the US National Registry of Sudden Death in Athletes were interrogated to identify those events judged to be cardiovascular in origin, occurring in organized competitive interscholastic sports participants."<br>Quote: "Individual athletes were included in the registry when identified through the aforementioned sources if 2 criteria were met: (1) participation in organized team or individual sports that required regular competition against others as a central component, placed a high premium on excellence and achievement, and required systematic and, in most instances, vigorous training (individuals participating in college- sponsored intramural or club sports were not included); and (2) SD r39 years of age. SD was defined as an unexpected collapse (with or without physical exertion) associated with a previously uneventful clinical course."<br>Quote: "In 1998, we reported the frequency of SDs in Minnesota high school athletes (up to 1996) to be 1:217,000 participants. This analysis was confined to deaths occurring specifically during in-season interscholastic high school competition and practice. In the present study, we expanded our analysis of this cohort, both with respect to the length of observation (ie, by 15 additional years) and by including all deaths not necessarily limited to those occurring in season for a particular sport. Consequently, the total number of deaths reported here (n ¼ 13) exceeds by 4-fold that in the initial report, and the present study also includes an at-risk population that is 3-fold larger." |

| | | |
|---|---|---|
| | | Quote: "At the time of collapse, 7 athletes were engaged in organized competition and 6 in practice or training."<br>Judgement Comment: The calculation of the cases is somewhat opaque in this article. The abstract mentions only SCD during exertion in organized HS sport activity, yet the discussion reports including deaths occurring outside of sport. The Methodology in the article does not report this-it states those included in the registry are with and without exertion. In other articles from this registry, deaths in athletes at any time are considered. The article title is also misleading as it reports HS athletes, but includes ages 12-18, which will include middle schoolers. Risk of bias associated with this article is the different methodology for this count compared to other articles relating to this registry. There is also the issue of the authors choosing different time periods to interrogate, other articles report 1980 as the starting point for this registry, yet this article starts in 1986 with no explanation. The authors also include athletes with sudden collapse with no explanation as SCD (probably appropriately), but do not do this in other articles in this series. |
| EPIDEMIOLOGY INTERNAL VALIDITY: MODE OF DATA COLLECTION | Low risk | Quote: "previously uneventful clinical course. A systematic tracking process was established to assemble detailed information on each case, which included the autopsy report (with gross anatomic, histological, and toxicological findings) and pertinent clinical and demographic information. Selected data (eg, circumstances of collapse) were often derived from written accounts or telephone interviews with family members, witnesses, or coaches. When necessary, autopsy findings were verified by direct communication with medical examiners, and primary pathological materials were selectively requested and analyzed." |
| EPIDEMIOLOGY FINAL SUMMARY: | Moderate risk | Judgement Comment: The article presented on its own appears to have low risk of bias. However, when reviewed with other articles in the series published from this registry, there is concern for significant bias relating to small differences in methodology, such as including the MN catastrophic |

| | | |
|---|---|---|
| | | insurance data in this article (which would likely decrease bias); contradictions in who is counted as deaths in this study, the abstract indicates only deaths during organized sport activity (which is true for all 13 deaths), but the discussion and methodology report deaths at any time. There is also the matter of choosing different periods of time to report deaths in the different articles in this series without explanation. This article as a stand alone piece has reason for concern based on counting methods, and conflicts in abstract and methodology; taken in context of other articles published in this series, there is a high risk of bias based on different reporting methods, and the risk of selective outcome reporting |
| EPIDEMIOLOGY EXT VALIDITY: NONRESPONSE BIAS/FOLLOW UP OF COHORT | N/A | |

#### *Maron 2016*

| | |
|---|---|
| **Methods** | **Study design:** Retrospective cohort study<br>**Retrospective cohort:** Review of databased collected for 32 years. Appears to be prospective collection, but methodology and times collected not well described. |
| **Participants** | **Included criteria:** Sudden deaths occurring during sport. Further categorized to cardiac, probable cardiac, and non-cardiac. In this article the inclusion criteria appears different than in previous article; 'Sports participants are considered for inclusion if they engaged in an organized team or individual sport requiring regular training and competition, and experienced sudden death.' In 2009 article, authors sought to include older competitive athletes, as well as younger than high school athletes. This is not commented on in this article. |

| | |
|---|---|
| | **Excluded criteria:** Deaths occurring in club or intramural sports, or resulting from automobile accidents, cancer, and other systemic diseases are not included<br>**Age:** 15-24<br>**Gender:** both<br>**Sport:** combination<br>**Level of Sport:** combination<br>**ECG Screened:** not described |
| **Interventions** | Observational |
| **Outcomes** | **SCA:** no<br>**SCD:** yes<br>**Exertional:** no, all deaths<br>**Method of ID Cases:** combination medical/autopsy reports, web/media reports included in US Registry Sudden Death in Athletes |
| **Identification** | **Sponsorship source:**<br>**Country:** USA<br>**Setting:** All athletes 39 years of age and younger<br>**Comments:**<br>**Authors name:** Barry Maron, MD<br>**Institution:** Minneapolis Heart Institute Foundatio<br>**Email:**<br>**Address:** Minneapolis Heart Institute Foundation, 920 E. 28th Street, Suite 620, Minneapolis, MN 55407 |
| **Notes** | |

Risk of bias table

| **Bias** | **Authors' judgement** | **Support for judgement** |
|---|---|---|
| EPIDEMIOLOGY EXT VALIDITY: REPRESENTATIVE SAMPLE | High risk | Judgement Comment: Ages reported on for incidence are selected for unclear reasons by the authors. |
| EPIDEMIOLOGY INTERNAL VALIDITY: CASE DEFINITION | Low risk | Quote: "464 other athletes experienced virtually instantaneous collapse during or immediately after physical activity, which were judged probable cardiovascular deaths but remained without a definitive clinical and/or autopsy diagnosis (given ambiguous or absent autopsy reports)."<br>Judgement Comment: Appear to be an appropriate decision about cause of death, and assigned as SCD only when data supported this. |
| EPIDEMIOLOGY EXT VALIDITY: | High risk | Quote: "athletes participating in competitive athletics. Sports participants are considered for |

| | | |
|---|---|---|
| SAMPLING/SUBJECTS INCLUDED | | inclusion if they engaged in an organized team or individual sport requiring regular training and competition, and experienced sudden death. 1-3,8,12-20 Deaths occurring in club or intramural sports, or resulting from automobile accidents, cancer, and other systemic diseases are not included. This project was approved by"<br>Judgement Comment: It is not explained why the data on incidence includes only a selection of the athletes based on gender, and based on race. It is also notable that in these two calculations, different age ranges were used. The definition of who was included appears to differ from the 2009 article. |
| EPIDEMIOLOGY INTERNAL VALIDITY: STUDY PERIOD | Low risk | > 100,000 person years |
| EPIDEMIOLOGY INTERNAL VALIDITY: DATA COLLECTION SOURCE | Low risk | Quote: "New York Institutional Review Board. <b>A variety of sources 7,12-14,17-19,21 were used to identify the study population by targeted searches: (1) LexisNexis archival informational database with searchable access to authoritative news, legal, and public records; (2) National Collegiate Athletic Association Memorial Resolutions List; (3) news media accounts systematically assembled through Burrelle's Information Services (Livingston, NJ); (4) internet search engines (eg, Google, Yahoo); (5) reports from the US Consumer Product Safety Commission (Washington, DC); (6) records of the National Center for Catastrophic Sports Injury Research (University of North Carolina, Chapel Hill, NC); (7) reports submitted to the Registry through personal contact with physicians, attorneys, coroners/ medical examiners, schools, and patient advocacy/support organizations "<br>Judgement Comment: Discussed in 2009 article, |
| EPIDEMIOLOGY INTERNAL VALIDITY: NUMERATOR AND DENOMINATOR | High risk | Quote: "To calculate the number of athlete-years, stratified by gender and race, available public domain data on student participation were assembled from the National Federation of State High School Associations 23 and the National Collegiate Athletic Association. 24 In each of these analyses, the number of tabulated participations was con- verted to the number of athlete-years |

| | | |
|---|---|---|
| | | relying on the published correction factors (ie, 1.9 for high school and 1.2 for college). 13 To calculate the incidence of confirmed cardiovascular sudden deaths by gender, the database was reviewed, and deaths among athletes aged 15-24 years were selected. Similarly, to calculate the incidence rate of confirmed cardiovascular sudden deaths by race, the database was reviewed, and deaths among athletes aged 19-24 years were selected."<br>Quote: "Of the 842 athletes, most (452 [54%]) were white, 350 (42%) were African American, and 40 (5%) were other minorities."<br>Quote: "incidence, cardiovascular mortality in African Americans and other minorities exceeded"<br>Quote: "that in whites by a factor of 4.8 (1:12,778 and 1:60,746 athlete-years, respectively; P <.001)."<br>Judgement Comment: No overall denominator given here. Only incidence rates given are by race, and gender. These are determined in different age groups for reasons not explained by author. No overall incidence rate given for unclear reason. 464 athlete deaths in the registry were not counted bc no clear cause of SD ascertained, but died suddenly during or right after activity. 842 of of 2406 total sudden deaths determined as SCD. When extracting data on incidence, it appears that these 842 are accounted for in ages 15-24-but this is only noted after extracting data, and doing backwards calculations. Not clear why this is; unless the authors wished to give more impressive incidence rates. Whole cohort goes to age 39, but only up to age 24 included in incidence rates-may be for reasons of calculating a denominator by using NFHS and NCAA data. When calculating black incidence rate, 40 'other' race deaths were added to total of 350 black SCD for the calculation. No reason given for this. |
| EPIDEMIOLOGY INTERNAL VALIDITY: MODE OF DATA COLLECTION | High risk | Quote: "Reports to the Registry of sudden deaths have increased at 4.4%/year (P <.001; 95% confidence interval [CI] 3.9%- 4.8%), 1.5%/year for males (P <.001; 95% CI 1.4%-1.7%), and 5.7%/year for females (P <.001; 95% CI 5.1%-6.2%)."<br>Judgement Comment: Over the life of the registry, reports have increased annually, suggesting possible |

| | | |
|---|---|---|
| | | in athletes participating in sports, vs. increase in awareness of its existence and better news report search methods (which I believe). It appears the data later in the life of the registry has better information, making the data from early in the life particularly questionable. It is also questionable how valid media reports are as the primary driver of deaths. The authors do not breakdown what proportion of deaths are from what sources. |
| EPIDEMIOLOGY FINAL SUMMARY: | High risk | Judgement Comment: The methods in this article are different from same database report 7 years earlier. Different incidence rates reported in this article; different age criteria for reporting in this article. Not clear on choices for reporting incidence rates in this article, and why no overall incidence rate reported for the whole cohort, vs. the selectively reported incidence rate. |
| EPIDEMIOLOGY EXT VALIDITY: NONRESPONSE BIAS/FOLLOW UP OF COHORT | N/A | |

#### *Maron 2016a*

| | |
|---|---|
| **Methods** | **Study design:** Retrospective cohort study<br>**Prospective cohort:** Includes deaths in HS and college athletes in Hennepin county, Minnesota |
| **Participants** | **Included criteria:** Deaths autopsied and reported as sudden cardiac death, in high school and college athletes.<br>**Excluded criteria:** none listed-only competitive athletes included<br>**Age:** 14-23<br>**Gender:** both<br>**Sport:** combination<br>**Level of Sport:** combination (scholastic and university)<br>**ECG Screened:** no |
| **Interventions** | Observational |
| **Outcomes** | **SCA:** no<br>**SCD:** yes |

| | |
|---|---|
| | **Exertional:** no, all deaths<br>**Method of ID Cases:** autopsy reports |
| **Identification** | **Sponsorship source:**<br>**Country:** USA<br>**Setting:** Hennepin County Minnesota competitive high school and college athletes<br>**Comments:**<br>**Authors name:** Barry Maron, MD<br>**Institution:** Hypertrophic Cardiomyopathy Center Minneapolis Heart Institute Foundation<br>**Email:**<br>**Address:** |
| **Notes** | |

Risk of bias table

| **Bias** | **Authors' judgement** | **Support for judgement** |
|---|---|---|
| EPIDEMIOLOGY EXT VALIDITY: REPRESENTATIVE SAMPLE | Unclear risk | Judgement Comment: It is not clear how many young competitive athletes are not included in this study (who play outside of high school and college sports); but certainly collecting data only on this group will exclude some competitive athletes. |
| EPIDEMIOLOGY INTERNAL VALIDITY: CASE DEFINITION | High risk | Quote: "A competitive student athlete in organized high school and college sports programs was defined as: one who participates in an organized team or individual sport that requires regular competition against others as a central component, places a high premium on excellence and achievement, and requires some form of systematic (and usually intense) training."<br>Judgement Comment: Authors clear about who they are reporting on. |
| EPIDEMIOLOGY EXT VALIDITY: SAMPLING/SUBJECTS INCLUDED | High risk | Judgement Comment: Only including HS and college athletes would appear to underestimate the number of even competitive athletes in that age range. Missing those adolescents who only participate in youth sports clubs. |
| EPIDEMIOLOGY INTERNAL VALIDITY: STUDY PERIOD | Low risk | > 100,000 person years |
| EPIDEMIOLOGY INTERNAL VALIDITY: DATA COLLECTION SOURCE | Low risk | Judgement Comment: Autopsy records |

| | | |
|---|---|---|
| EPIDEMIOLOGY INTERNAL VALIDITY: NUMERATOR AND DENOMINATOR | Low risk | Quote: "all sudden deaths <40 years of age undergo complete autopsy and toxicologic studies."<br>Quote: "The database was assessed to identify naturally occurring sudden cardiovascular deaths, age 14 to 23 years, 2000 to 2014. In addition to the Medical Examiner evaluation, gross and histopathologic cardiac ex- aminations were conducted by expert cardiovascular pathologists at the Jesse E. Edwards Registry"<br>Quote: "To calculate the relative incidence of competitive athlete versus nonathlete events, we constructed the size of at-risk populations within Hennepin County, 2000 to 2014, from publicly available data. Specifically, this included total student enrollment in those high schools (n ¼ 131) and colleges (n ¼ 4) with athletic programs that practice some measure of preparticipation screening for athletes, usually history and physical examination. 1 In Hennepin County, athlete participation rates for individual colleges and high schools were estimated by using combined data from the National Center for Educational Statistics and the Minnesota"<br>Quote: "State High School League Sponsored Activity Participation Survey."<br>Judgement Comment: Cases were ID'd from autopsy-as all deaths under age 40 must be autopsied. Denominator from database-only included HS and college athletes. No club athletes, or athletes competing outside of these teams. |
| EPIDEMIOLOGY INTERNAL VALIDITY: MODE OF DATA COLLECTION | Low risk | Judgement Comment: All data from autopsy records/database. |
| EPIDEMIOLOGY FINAL SUMMARY: | Moderate risk | Judgement Comment: The authors are clear about who is included, but try to generalize this number as a good representation of the death rate in competitive athletes, without acknowledging that only including scholastic athletes may undercount the number of deaths. The number counted appears accurate, and there appears to be little room for bias when reporting this rate in HS and collegiate athletes in this county, but misses those not playing these sports |

| | |
|---|---|
| EPIDEMIOLOGY EXT VALIDITY: NONRESPONSE BIAS/FOLLOW UP OF COHORT | N/A |

#### *Maurice 2018*

| | |
|---|---|
| **Methods** | **Study design:** Annual phone survey over 5 year period to local Buenos Aires rugby club for cardiovascular events |
| **Participants** | **Included criteria:** Rugby players at clubs participating in the phone survey.<br>**Excluded criteria:** none listed<br>**Age:** Unknown-not described<br>**Gender:** not described, assumed to be both<br>**Sport:** rugby<br>**Level of Sport:** competitive<br>**ECG Screened:** not described |
| **Interventions** | Observational |
| **Outcomes** | **SCA:** yes<br>**SCD:** yes<br>**Exertional:** yes<br>**Method of ID Cases:** club response to phone survey |
| **Identification** | **Sponsorship source:** none listed<br>**Country:** Argentina<br>**Setting:** Rugby Clubs in the Buenos Aires Rugby Union, and provincial rugby unions in Argentina<br>**Comments:**<br>**Authors name:** Mario Fitz Maurice<br>**Institution:** Hospital Bernardino Rivadavia<br>**Email:**<br>**Address:** Servicio de Cardiologia Hopsital Bernardino Rivadavia Av. Las Heras2670 (1425) CABA Buenos Aires, Argentina |
| **Notes** | |

Risk of bias table

| **Bias** | **Authors' judgement** | **Support for judgement** |
|---|---|---|
| EPIDEMIOLOGY EXT VALIDITY: REPRESENTATIVE SAMPLE | Unclear risk | Judgement Comment: Demographics not reported, but this includes 78% of rugby clubs in Buenos Aires region, would seem to be a good representation of |

| | | |
|---|---|---|
| | | rugby athletes in this area. Without any demographic data however, the risk of bias remains unclear. |
| EPIDEMIOLOGY INTERNAL VALIDITY: CASE DEFINITION | High risk | Judgement Comment: Case definition appears to be that an AED was used after a collapse. |
| EPIDEMIOLOGY EXT VALIDITY: SAMPLING/SUBJECTS INCLUDED | Low risk | Judgement Comment: The study includes 78% of the clubs available; the study should provide a reasonable picture of members of rugby clubs in the Buenos Aires region. |
| EPIDEMIOLOGY INTERNAL VALIDITY: STUDY PERIOD | Low risk | > 100,000 person years<br>Judgement Comment: 85000+ athletes for 5 years |
| EPIDEMIOLOGY INTERNAL VALIDITY: DATA COLLECTION SOURCE | High risk | Judgement Comment: Data came from phone survey simply asking:' How many subjects required AED?", was it a "player or spectator"?, "how many of those people died?", and lastly, "How many patients died before arriving at a healthcare institution?" No further data was reported to have been collected. |
| EPIDEMIOLOGY INTERNAL VALIDITY: NUMERATOR AND DENOMINATOR | Unclear risk | Judgement Comment: Cases identified with phone report, with no detail given about how they were ID'd as SCA/D cases apart from AED usage. Over 30 clubs in the consortium did not have AEDs, and were not included in the survey. |
| EPIDEMIOLOGY INTERNAL VALIDITY: MODE OF DATA COLLECTION | Unclear risk | Judgement Comment: Phone survey was used annually for five years. There is no reporting on the overall participation, except to say that 107 of 137 clubs participated, and that the 30 that did not did not have an AED. |
| EPIDEMIOLOGY FINAL SUMMARY: | High risk | Judgement Comment: Phone survey with little detail regarding the identification of SCA/D events; exclusion of clubs without AED bias’s sample. |
| EPIDEMIOLOGY EXT VALIDITY: NONRESPONSE BIAS/FOLLOW UP OF COHORT | Low risk | Judgement Comment: 78% participation in the survey 78% participation in the survey |

#### *Peterson 2020*

| | |
|---|---|
| **Methods** | **Study design:** Prospective Cohort |

| | |
|---|---|
| **Participants** | **Included criteria:** competitive athletes from ages of 11-29; incidence only for high school, and collegiate athletes<br>**Excluded criteria:** suspected but not confirmed cases of SCA/D<br>**Age:** 11-29<br>**Gender: all**<br>**Sport: all**<br>**Level of Sport:** competitive<br>**ECG Screened:** not reported |
| **Interventions** | Observational |
| **Outcomes** | **SCA:** Yes<br>**SCD:** Yes<br>**Exertional:** Yes<br>**Method of ID Cases:** Media traditional and social; reporting to NCCSIR, UW center for sports cardiology; search of student-athlete deaths on the NCAA Resolutions List; direct communication with the NFHS; and regular review of cases collected in the Parent Heart Watch database. |
| **Identification** | **Sponsorship source:** NCCSIR; NCAA; NFSHSA, AFCA; NATA; NOCSAE; AMSSM; AOA student research fellowship<br>**Country:** USA<br>**Setting:** club; scholastic; HS; University and professional athletes<br>**Comments:**<br>**Authors name:** Danielle F Peterson; Senior author Jonathan Drezner<br>**Institution:** Oregon Health and Science University, Portland, OR<br>**Email:**<br>**Address:** Sports Medicine Center, University of Washington, Seattle, WA 98195 |
| **Notes** | |

Risk of bias table

| **Bias** | **Authors' judgement** | **Support for judgement** |
|---|---|---|
| EPIDEMIOLOGY EXT VALIDITY: REPRESENTATIVE SAMPLE | Low Risk | Judgement comment: 'Competitive athletes at the middle school (ages 11–13), high school including premiere/select level (ages 14–18), college (ages 19–23), semiprofessional and professional levels (ages 24–29) who experienced SCA/D were included. A competitive athlete was defined as an individual at least 11 years of age involved in regular training in an organised individual or team sport with an emphasis on competition and performance. Furthermore, former athletes were included if the cardiac event occurred within 1year of competitive |

| | | |
|---|---|---|
| | | sportsparticipation in one of the previously described categories.'<br>Appears to include all athletes in the US-only incidence reported in high school and college level |
| EPIDEMIOLOGY INTERNAL VALIDITY: CASE DEFINITION | Low Risk | Judgement Comment: All cases of confirmed SCA/D in a competitive athlete were included regardless of the activity at the time of the event, including those occurring during exercise, within 1 hour of exer- cise, at rest or during sleep. Cases occurring during exercise in which autopsy or medical records could not be obtained were included as cardiac in nature if the event details supported an abrupt collapse requiring cardiac resuscitation. Cases of possible SCA/D were excluded if participation as a competitive athlete could not be confirmed, a cardiac aetiology could not be deter- mined (including autopsy negative cases) when occurring during rest or sleep, or a non-cardiac diagnosis was identified.<br><br>Thorough accounting of cases; adjudicated by panel to ID deaths |
| EPIDEMIOLOGY EXT VALIDITY: SAMPLING/SUBJECTS INCLUDED | Low risk | Judgement Comment: Yes-as above, all included athletes identified |
| EPIDEMIOLOGY INTERNAL VALIDITY: STUDY PERIOD | Low risk | > 100,000 person years<br>Judgement Comment: Yes-large cohort over four years prospectively. |
| EPIDEMIOLOGY INTERNAL VALIDITY: DATA COLLECTION SOURCE | Low risk | Judgement Comment: Multiple sources used for ID of athletes, including national centers accepting reporting on sudden cardiac death; sport institutions receving reporting, and backed up by social and traditional media searches for information. |
| EPIDEMIOLOGY INTERNAL VALIDITY: NUMERATOR AND DENOMINATOR | Low risk | Judgement Comment: 'Athlete population statistics were retrieved from the NFHS and NCAA in which participation data is provided per sport. Participation data for middle school athletics, the National Asso- ciation of Intercollegiate Athletics, junior/community colleges, semiprofessional and professional levels was not available, and therefore, incidence rates |

| | | |
|---|---|---|
| | | could not be calculated at these levels. Race participation data were available for NCAA athletes; however, race statistics are not reported by the NFHS. All inci- dence rates are reported per athlete-years (AY) of participation. At the high school level, some athletes play multiple sports but should account for only one AY of participation. In calculating the overall incidence at the high school level, a conversion factor of 2.35 was applied to account for multisport athletes as used in prior studies.4 7 18 19 A conversion factor was not used for calcu- lation of sport-specific incidence rates in high school athletes as the actual participation statistics were available. Sport-specific incidence was calculated for sports in which five or more SCA/D events occurred during the study period. Incidence was further subdivided by race and NCAA division at the collegiate level. The relative risk of SCA/D in male college athletes by sport and race was compared with the risk in male high school athletes.<br>Because case capture and reporting mechanisms vary per state within the USA, we also conducted an exploratory analysis of the incidence of SCA/D in male high school athletes. State-specific participation statistics for high school athletes was obtained for each state based on NFHS data. We chose to calculate the inci- dence of SCA/D in male high school athletes in states with five or more SCA/D cases as incidence rates with fewer than five events may be unreliable. A similar analysis could not be conducted for female high school athletes due to the low number of SCA/D cases.'<br><br>Appears very thorough. |
| EPIDEMIOLOGY INTERNAL VALIDITY: MODE OF DATA COLLECTION | Unclear risk | Judgement Comment: As above, multiple data sources started prospectively and followed for 4 years. |
| EPIDEMIOLOGY FINAL SUMMARY: | Low risk | Judgement Comment: Low risk. Some concern about the selective reporting of subgroup-but overall appears well constructed and as thorough as possible in the United States. |

| | | |
|---|---|---|
| EPIDEMIOLOGY EXT VALIDITY: NONRESPONSE BIAS/FOLLOW UP OF COHORT | NA | NA |

#### *Phillips 1986*

| | |
|---|---|
| **Methods** | **Study design:** Retrospective cohort study<br>**Retrospective cohort:** Not explicitly described, but appears to be retrospective review of autopsy records. |
| **Participants** | **Included criteria:** Healthy Air Force recruits who have passed their entrance physicals. Exertional deaths only were included. Deaths with activity, or within 1 hour.<br>**Excluded criteria:** none listed<br>**Age:** 18-28<br>**Gender:** both<br>**Sport:** military<br>**Level of Sport:** military<br>**ECG Screened:** no |
| **Interventions** | Observational |
| **Outcomes** | **SCA:** no<br>**SCD:** yes<br>**Exertional:** yes<br>**Method of ID Cases:** autopsy reports from military records |
| **Identification** | **Sponsorship source:** None Listed<br>**Country:** USA<br>**Setting:** Air Force Basic Training Recruits<br>**Comments:**<br>**Authors name:** Major Matthew Phillips, MC, USAF<br>**Institution:** Department of Cardiology, Wilford Hall Medical Center, Lackland Air Force Base<br>**Email:**<br>**Address:** Dept of Cardiovascular Pathology Armed Forces Institute of Pathology Washington, DC 20306-6000 |
| **Notes** | Calculated 100,000 person years from the 100,000 participants for 42 days they provide. |

Risk of bias table

| **Bias** | **Authors' judgement** | **Support for judgement** |
|---|---|---|

| | | |
|---|---|---|
| EPIDEMIOLOGY EXT VALIDITY: REPRESENTATIVE SAMPLE | Low risk | Judgement Comment: Only reported for air force recruits. There is no demographic data reported. |
| EPIDEMIOLOGY INTERNAL VALIDITY: CASE DEFINITION | Low risk | Quote: "Death was defined as sudden if it occurred within one hour of the onset of symptoms. Death was considered to be cardiac in origin if there was either pathologically confirmed heart disease or clinical cardiac arrest." |
| EPIDEMIOLOGY EXT VALIDITY: SAMPLING/SUBJECTS INCLUDED | Low risk | Judgement Comment: All basic training recruits for 21 years included. |
| EPIDEMIOLOGY INTERNAL VALIDITY: STUDY PERIOD | Low risk | > 100,000 person years<br>Judgement Comment: Calculated person years of basic training. 180,000+ |
| EPIDEMIOLOGY INTERNAL VALIDITY: DATA COLLECTION SOURCE | Low risk | Judgement Comment: Military and autopsy records. |
| EPIDEMIOLOGY INTERNAL VALIDITY: NUMERATOR AND DENOMINATOR | Low risk | Quote: "A total of 1 606 167 recruits participated in the 42-day basic military training program during the period 1965 through 1985."<br>Quote: "The Basic Air Force Military Training Program was a 42-day program with 30 active training days that included a minimum of one hour per day of exercise."<br>Judgement Comment: Closed cohort of air force basic training recruits. The deaths were counted and COD from autopsy; the denominator developed from total recruits (1606167) for 42 days/365. Developed via 1606167 x (42/365)=184819 person years of basic training. |
| EPIDEMIOLOGY INTERNAL VALIDITY: MODE OF DATA COLLECTION | Low risk | Quote: "The history, clinical records, and circumstances of death were analyzed. Microscopic sections from 19 of 21 cardiac death victims and 17 of the 22 noncardiac deaths were reviewed blindly and independently by two cardiovascular pathologists. A total of 158 microscopic sections of the heart were examined (range, one to 19 sections; mean, five per case)."<br>Judgement Comment: Military records, including autopsy records. |

| | | |
|---|---|---|
| EPIDEMIOLOGY FINAL SUMMARY: | Low risk | Judgement Comment: Appears to be well done study on deaths in Air force recruits. Deaths during basic training reviewed; all basic training recruits included. |
| EPIDEMIOLOGY EXT VALIDITY: NONRESPONSE BIAS/FOLLOW UP OF COHORT | N/A | |

#### *Risgaard 2014*

| | |
|---|---|
| **Methods** | **Study design:** Retrospective cohort study<br>**Retrospective cohort:** Review of death certificate and autopsy data |
| **Participants** | **Included criteria:** Exertional deaths for sport related sudden cardiac death. All citizens of Denmark ages 12-49 included in the cohort.<br>**Excluded criteria:** none listed<br>**Age:** 12-49 for total cohort. We extracted 12-35 in competitive athletes only.<br>**Gender:** both<br>**Sport:** combination<br>**Level of Sport:** combination<br>**ECG Screened:** no |
| **Interventions** | Observational |
| **Outcomes** | **SCA:** no<br>**SCD:** yes<br>**Exertional:** yes<br>**Method of ID Cases:** combination of autopsy reports/medical records and web/media reports |
| **Identification** | **Sponsorship source:** Danish National Research Foundation Centre for Cardiac Arrhythmia (DARC), University of Copenhagen, Copenhagen, Denmark; Laboratory of Molecular Cardiology, The Heart Centre, Department of Cardiology, University Hospital Rigshospitalet, Copenhagen, Denmark; The John and Birthe Meyer Foundation; The Danish Heart Foundation (12-04-R91-A3790-22689); and The Research Fund of Rigshospitalet, Copenhagen University Hospital. The Danish Health Profile 2010 was funded by The Capital Region, Region Zealand, The South Denmark Region,The Central Denmark Region, The North Denmark Region, The Danish Ministry of Interior and Health, and the National Institute of Public Health, University of Southern Denmark.<br>**Country:** Denmark |

| | |
|---|---|
| | **Setting:** All citizens of Denmark<br>**Comments:** Breakdown of deaths into age groups, all citizens, competitive and non-competitive athletes.<br>**Authors name:** Bjarke Risgaard, MD<br>**Institution:** Danish National Research Foundation Centre for Cardiac Arrhythmia (DARC), Copenhagen, Denmark<br>**Email:**<br>**Address:** Laboratory of Molecular Cardiology, The Heart Centre Copenhagen University Hospital Rigshospitalet, 9312, Juliane Maries Vej 20,2100 Copenhagen |
| **Notes** | |

Risk of bias table

| **Bias** | **Authors' judgement** | **Support for judgement** |
|---|---|---|
| EPIDEMIOLOGY EXT VALIDITY: REPRESENTATIVE SAMPLE | Low risk | Judgement Comment: Appears to well represent Northern Europeans. |
| EPIDEMIOLOGY INTERNAL VALIDITY: CASE DEFINITION | Low risk | Quote: "We defined sudden unexpected death as sudden, natural unexpected death; in witnessed cases as an acute change in cardiovascular status with time to death being o1 hour and"<br>Quote: " in unwitnessed cases as a person last seen alive and functioning normally 24 hours before being found dead. Sudden cardiac death (SCD) in autopsied cases was defined as the natural unexpected death of unknown or cardiac cause; in witnessed cases as an acute change in cardiovascular status with time to death being 1 hour and in unwitnessed cases as a person last seen alive and functioning normally 24 hours before being found dead. 16–19 Autopsied SCD was subdivided into 2 groups: (1) explained SCD, in which a cardiac cause of death was established, and (2) sudden unexplained death (SUD), in which a cause of death after autopsy remained unknown. In nonautopsied deaths, the same criteria were used in cases presumed to be of cardiac origin based on the circum- stances relating to the death, including all information from death certificates and discharge summaries. Nonautopsied cases with no competing causes of death were considered to be of cardiac origin. Sport related sudden cardiac death (SrSCD) |

| | | |
|---|---|---|
| | | was defined as an SCD occurring during or within 1 hour after moderate- to high-intensity exercise. Using all the information available,"<br>Quote: "Using all the information available, we subdivided SrSCD into whether the deceased was considered a competitive or a noncompetitive athlete. The deceased was considered a competitive athlete if he or she did moderate- to high- intensity sports on a regular level and took part in competitions. For the purpose of this study, a noncompetitive athlete was defined as a person not participating in competitions but who did moderate- to high-intensity sports on a regular level in the months before death." |
| EPIDEMIOLOGY EXT VALIDITY: SAMPLING/SUBJECTS INCLUDED | Low risk | Judgement Comment: All Danish citizens included. Article details deaths in competitive athletes, non-competitive athletes, and non-athletes. As well as breakdown of ages 12-35, and 36-49. |
| EPIDEMIOLOGY INTERNAL VALIDITY: STUDY PERIOD | Low risk | > 100,000 person years |
| EPIDEMIOLOGY INTERNAL VALIDITY: DATA COLLECTION SOURCE | Low risk | Quote: "Death certificates were read independently by 2 physicians in order to identify deaths that were sudden and unexpected. In case of disagreement, a consensus were reached after reevaluating the circumstances surrounding the death, including a review of previous medical history, which was taken into account in every single case. Danish death certificates are informative and suitable for identifying sudden unexpected deaths because they have a supplemental information field (see Online Supplemental Data and Winkel et al). This field contains information describing circum- stances surrounding the death, including interviews with eyewitnesses and relatives, previous medical conditions, an external examination of the body, and the preliminary conclusion before autopsy. This field is mandatory in all medicolegal external examinations (external examinations), including cases in which it is decided not to conduct an autopsy. Danish death certificates can only be issued by a medical doctor. In cases where citizens or patients are found dead and/or the death is |

| | | |
|---|---|---|
| | | sudden and unexpected, external examinations are mandatory by law."<br>Quote: "To apply a method used by others when identifying SrSCD, an additional extensive retrospective media search was performed as a supplement to the main approach. The Danish media surveillance database Infomedia (www. infomedia.dk), a database of approximately 400 printed, 2200 web-based, and major radio and television Danish media, was used."<br>Quote: "For the purpose of this study, respondents who chose category 1 was considered competitive athletes, and respondents who chose category 2 was considered noncompetitive athletes. Because the age range in the "How Are You?" study did not match ours, the assumption was made that we could extrapolate information from the population aged 16-49 years to the population aged 12-49 years." |
| EPIDEMIOLOGY INTERNAL VALIDITY: NUMERATOR AND DENOMINATOR | Low risk | Quote: "To estimate the size of the background population performing competitive and noncompetitive sports activities, on a regular level (the denominator), we used data from the Danish National Institute of Public Health."<br>Quote: "From the review of the death certificates, registry information on previous medical history, discharge summaries, and media search, we identified 881 SCD, of which 44 cases were SrSCD. Thirty-three of the 44 SrSCD occurred in noncompetitive athlete, whereas 11 cases occurred in competitive athletes (Figure 2). Of the 2 cases identified through media search, 1 was identified from the review of the death certificates. The remaining case was identified and included after review of discharge summaries among the 26 cases with incomplete death certificate data (page 2 of death certificate missing; Figure 1)."<br>Judgement Comment: Only risk here appears to be that of extrapolating data for denominator from the national reporting survey on age 16-49, to ages 12-49. Otherwise very well explained. |

| | | |
|---|---|---|
| EPIDEMIOLOGY INTERNAL VALIDITY: MODE OF DATA COLLECTION | Low risk | Quote: "In brief, this is a nationwide retrospective study using the availability of all death certificates and the registration of all inpatient and outpatient activity in Danish hospitals and emergency rooms together with access to all medical records and autopsy reports. We included all deaths in a 3-year period (2007-2009) in persons aged 12-49 years." Judgement Comment: Also included web search as a backup. |
| EPIDEMIOLOGY FINAL SUMMARY: | Low risk | Judgement Comment: Very nice study. The database appears very thorough, especially when compared with web search, and finding good agreement. The authors are very thorough in describing data source, case definition, numerator and denominator, and overall methodology. |
| EPIDEMIOLOGY EXT VALIDITY: NONRESPONSE BIAS/FOLLOW UP OF COHORT | N/A | |

#### *Roberts 2013*

| | |
|---|---|
| **Methods** | **Study design:** Retrospective cohort study<br>**Retrospective cohort:** Catastrophic insurance records of deaths occurring while participating in organized scholastic sport activity |
| **Participants** | **Included criteria:** Deaths occurring while participating in Minnesota scholastic sport activity organized (such as practice, or game). Exertional. All MN scholastic athletes included in denominator<br>**Excluded criteria:** none listed<br>**Age:** 12-19<br>**Gender:** both<br>**Sport:** combination<br>**Level of Sport:** scholastic<br>**ECG Screened:** no |
| **Interventions** | Observational |
| **Outcomes** | **SCA:** no<br>**SCD:** yes<br>**Exertional:** yes<br>**Method of ID Cases:** catastrophic insurance records for scholastic athletes in Minnesota |

| | |
|---|---|
| **Identification** | **Sponsorship source:** none listed<br>**Country:** USA<br>**Setting:** Minnesota Scholastic Athletes<br>**Comments:**<br>**Authors name:** William O. Roberts, MD<br>**Institution:** Department of Family Medicine and Community Health, University of Minnesota<br>**Email:**<br>**Address:** University of Minnesota Medical School Family Medicine and Community Health Phalen Village Clinic1414 Maryland Avenue East St. Paul, Minnesota 55106 |
| **Notes** | |

Risk of bias table

| **Bias** | **Authors' judgement** | **Support for judgement** |
|---|---|---|
| EPIDEMIOLOGY EXT VALIDITY: REPRESENTATIVE SAMPLE | Low risk | Quote: "limitation to extrapolating these data is that Minnesota has a lower proportion of African Americans than the general U.S. population, (5.4% vs. 13.1%, respectively), according to the last U.S. census statistics (9), and previous reports found that African-American athletes experience SCD at higher rates than whites (3). The data also should not be extrapolated to older age groups based on the NCAA study"<br>Judgement Comment: Meant to include MN scholastic athletes. The racial breakdown limits extrapolation to the US in general. Authors clear about their findings, and risk of applying these findings population in general. |
| EPIDEMIOLOGY INTERNAL VALIDITY: CASE DEFINITION | High risk | Quote: "The database does not include athletes who experienced a sudden cardiac arrest during MSHSL practice or games and lived, nor does it include SCD that occurred outside the auspices of MSHSL sports activities. The MSHSL has informally tracked cardiac events in Minnesota HS students since the year 2000 to look at the effectiveness of the League-sponsored emergency action plan that shows 12 student (presumed) cardiac events, including 8 students who lived due to the use of an automatic external defibrillator or an emergency action plan. Three of the 4 deaths occurred away from school campuses, and 7 students were not involved in HS sports (e.g., |

| | | |
|---|---|---|
| | | graduation party, open gym, gym class, basketball spectator). Thus, there were more cardiac events in this age group than were counted by our methods, but the denominator for this informal cohort is all Minnesota HS students rather than screened athletes alone."<br>Judgement Comment: No detail on how determination of cause of death was determined, apart from insurance records. Discussion reports the limitations of this methodology, and the likely undercount resulting from it. |
| EPIDEMIOLOGY EXT VALIDITY: SAMPLING/SUBJECTS INCLUDED | Low risk | Judgement Comment: Insurance database includes all those participating in MN scholastic sports. |
| EPIDEMIOLOGY INTERNAL VALIDITY: STUDY PERIOD | Low risk | > 100,000 person years |
| EPIDEMIOLOGY INTERNAL VALIDITY: DATA COLLECTION SOURCE | High risk | Judgement Comment: This methodology appears to miss a significant proportion of those SCA or SCD outside of organized sport activity. |
| EPIDEMIOLOGY INTERNAL VALIDITY: NUMERATOR AND DENOMINATOR | High risk | Quote: "In 1993, the MSHSL began to record individual athlete- years, in addition to the cumulative sports season participation that is reported annually to the National Federation of High Schools. The MSHSL requires that each athlete have catastrophic incident insurance, and the policy provides payment for sports-related deaths that occur during MSHSL games and practices. Payments are an accurate reflection of deaths that occur during MSHSL activities, and the numerator and denominator are concordant with the screened population at risk."<br>Quote: " The catastrophic insurance (required for all MSHSL athletes) records were used to find cardiac deaths that occurred during HS-related practice or games. All deaths that occur during MSHSL sports practices and games are reported for insurance purposes, and events not associated with MSHSL activities do not enter the reporting system. "<br>Quote: "Over 19 academic-years (from 1993/1994 through 2011/ 2012), there were 3,925,512 athlete-seasons, 2,085,366 boys and 1,739,168 girls. There were 1,666,509 unduplicated athletes participating in |

| | | |
|---|---|---|
| | | 1 sport per academic-year (2.35 sports seasons/athlete/academic-year)."<br>Judgement Comment: Demonstrated by other articles that including only the insurance data to gather cases underrepresents deaths in athletes. he authors themselves note this in the discussion. |
| EPIDEMIOLOGY INTERNAL VALIDITY: MODE OF DATA COLLECTION | Low risk | Judgement Comment: All cases from same insurance database. |
| EPIDEMIOLOGY FINAL SUMMARY: | High risk | Judgement Comment: As discussed in other articles, this methodology has risk of undercount of cases. The authors themselves note this in the discussion section of the article. For the purposes of counting deaths in athletes in high school, this article appears to be at high risk of bias. |
| EPIDEMIOLOGY EXT VALIDITY: NONRESPONSE BIAS/FOLLOW UP OF COHORT | N/A | |

#### *Santos Lozano 2017*

| | |
|---|---|
| **Methods** | **Study design:** Retrospective cohort study<br>**Retrospective cohort:** Retrospective web/news-based search for deaths of professional soccer players |
| **Participants** | **Included criteria:** Professional soccer players deaths within 1 hour of exertion/playing deaths confirmed with autopsy.<br>**Excluded criteria:** none listed<br>**Age:** no limitations listed, but deaths reported 16-34<br>**Gender:** both<br>**Sport:** soccer<br>**Level of Sport:** elite (professional)<br>**ECG Screened:** not detailed, portion was likely screened based on available information regarding European soccer leagues |
| **Interventions** | Observational |
| **Outcomes** | **SCA:** no<br>**SCD:** yes<br>**Exertional:** yes<br>**Method of ID Cases:** media/web reports |
| **Identification** | **Sponsorship source:** None listed |

| | |
|---|---|
| | **Country:** Spain<br>**Setting:** Worldwide professional soccer players<br>**Comments:**<br>**Authors name:** Alejandro Santos-Lozano, PhD<br>**Institution:** European University Miguel de Cervantes<br>**Email:**<br>**Address:** European University Miguel de Cervantes Padre Julio Chevalier, 247012 Valladolid, Spain |
| **Notes** | |

Risk of bias table

| **Bias** | **Authors' judgement** | **Support for judgement** |
|---|---|---|
| EPIDEMIOLOGY EXT VALIDITY: REPRESENTATIVE SAMPLE | Unclear risk | Quote: "Of note, we report a null incidence of SCD among players who underwent pre-participation screening, that is, those playing in the major European competitions."<br>Judgement Comment: It is unclear how likely players in developing countries are to have their deaths reported via searchable methods. The authors note that the cases that might be SCD but are not included are mainly in Africa and Asia. |
| EPIDEMIOLOGY INTERNAL VALIDITY: CASE DEFINITION | Low risk | Quote: " To be considered an SCD, each death had to fully comply with 3 stringent definition criteria (4): 1) unexpected as a result of natural causes; 2) post-mortem confirmation of the primary cause in the heart or great vessels (noncardiac causes excluded); and 3) occurring 1 h of the onset of collapse symptoms."<br>Quote: "One methodological limitation of these findings is the lack of reliable information on under-lying cardiac disease, autopsy data, and time from symptom onset to actual death for 21 of the 59 identified cardiovascular deaths (the vast majority in Asian and African countries), some of which might have fulfilled SCD criteria. If the aforementioned 21 cases were classified as SCDs, the incidence would have been 2.13 suspected SCDs/100,000 person-years (95% CI: 1.53 to 2.90). In the major European leagues (data on 45,506 players and 26,208 matches), only 1 cardiovascular death (occurring after resuscitated exertional cardiac arrest) was |

| | | |
|---|---|---|
| | | registered, yielding an incidence of 0 SCDs/100,000 person-years."<br>Judgement Comment: The included data appears to be autopsy only. There is the matter of 'suspected' cardiac death, which is not well explained, and the idea that 59 deaths were identified as cardiovascular events, and only 20 of these were included in the study due to what the authors describe as stringent criteria for case definition. |
| EPIDEMIOLOGY EXT VALIDITY: SAMPLING/SUBJECTS INCLUDED | Unclear risk | Judgement Comment: Authors report including all professional soccer players in this research. There is no detail on how they ensured or included all subjects. |
| EPIDEMIOLOGY INTERNAL VALIDITY: STUDY PERIOD | Low risk | > 100,000 person years |
| EPIDEMIOLOGY INTERNAL VALIDITY: DATA COLLECTION SOURCE | Low risk | Judgement Comment: Data were collected from autopsy, and witnesses. |
| EPIDEMIOLOGY INTERNAL VALIDITY: NUMERATOR AND DENOMINATOR | Unclear risk | Quote: "these, 59 were due to cardiovascular events (all men; mean age at death 25 Æ 5 years; incidence of 3.07 cardiovascular deaths/100,000 person-years; 95% confidence interval [CI]: 2.34 to 3.96), and 20 fully complied with the criteria of SCD, yielding an estimated worldwide incidence of 1.04 SCDs/100,000 person-years (95% CI: 0.85 to 1.26; mean age at death 24 Æ 5 years)"<br>Judgement Comment: Method of ID deaths is only via web report/news report. High risk of undercount in this setting. The denominator, or the method of determination of the denominator is not described. |
| EPIDEMIOLOGY INTERNAL VALIDITY: MODE OF DATA COLLECTION | High risk | Judgement Comment: News/web searches to ID the cases. |
| EPIDEMIOLOGY FINAL SUMMARY: | High risk | Judgement Comment: The risk of bias is high based on the use of web searches to ID the cases of SCD. There is also no real description of how denominators were identified. The authors also point out that more than 50 % of the cases they believe to be SCD, are not included in the final incidence number, due to lack of corroborating data. |

| | |
|---|---|
| EPIDEMIOLOGY EXT VALIDITY: NONRESPONSE BIAS/FOLLOW UP OF COHORT | N/A |

#### *Smallman 2016*

| | |
|---|---|
| **Methods** | **Study design:** Retrospective cohort study<br>**Retrospective cohort:** Review of detailed military data on deaths occurring with or associated with exercise |
| **Participants** | **Included criteria:** Active duty service members. Cases included if death occurred with exercise, or within 1 hour of exercise.<br>**Age:** all military members; < 35 yrsbreakdown given, which was extracted for our data<br>**Gender:** both<br>**Sport:** military<br>**Level of Sport:** military<br>**ECG Screened:** no |
| **Interventions** | Observational |
| **Outcomes** | **SCA:** no<br>**SCD:** yes<br>**Exertional:** yes<br>**Method of ID Cases:** military records and autopsy data |
| **Identification** | **Sponsorship source:** none listed<br>**Country:** USA<br>**Setting:** US active duty service members 4 major branches<br>**Comments:**<br>**Authors name:** Dr. Darlene P Smallman<br>**Institution:** Uniformed Services University of the Health Sciences<br>**Email:**<br>**Address:** Dr. Darlene P Smallman c/o Shelley McCallum USUHS Department of Preventive Medicine and Biostatistics Bldg A, Rm 1040A, 4301Jones Bridge Road Bethesda, MD 20814-4799USA |
| **Notes** | |

Risk of bias table

| **Bias** | **Authors' judgement** | **Support for judgement** |
|---|---|---|

| | | |
|---|---|---|
| EPIDEMIOLOGY EXT VALIDITY: REPRESENTATIVE SAMPLE | Low risk | Judgement Comment: Applies to military population. The article does present data which makes comparing this to athletes questionable. Of the cases < 35, 19% smokers, 42% overweight, and 26% obese-which would be an unlikely finding in athletes. |
| EPIDEMIOLOGY INTERNAL VALIDITY: CASE DEFINITION | Low risk | Quote: "Deaths were deemed cardiac in origin if there was (1) autopsy- confirmed heart disease with clinical circumstances consistent with a potential cardiac aetiology of death or (2) if the circum- stances of death were consistent with a cardiac arrhythmia in the absence of other conditions that could explain the death. 1 42 SCD/E was defined as (1) a death or initiation of terminal life support within 1 h of physical exertion or (2) an unwitnessed but unexpected death where the circumstances indicated that the individual had been exercising prior to death. If there was no pathology observed at autopsy to explain the sudden death, the case was classified as idiopathic SCD/E (iSCD/E)." Quote: "Toxicology screening was performed on 84% of patients with SCD/E, of which four were positive for illicit substances: one for ephedrine, one for amphetamine and two for tetrahydro- cannabinol; none was deemed causative." |
| EPIDEMIOLOGY EXT VALIDITY: SAMPLING/SUBJECTS INCLUDED | Low risk | Judgement Comment: All active duty members of 4 major military branches. |
| EPIDEMIOLOGY INTERNAL VALIDITY: STUDY PERIOD | Low risk | > 100,000 person years |
| EPIDEMIOLOGY INTERNAL VALIDITY: DATA COLLECTION SOURCE | Low risk | Quote: "All potential cases of SCD/E were obtained by querying the Armed Forces Medical Examiner System's (AFMES) Armed Forces Medical Examiner Tracking System (AFMETS) for deaths with a physical activity code during the surveillance period of 1 January 2005, through 31 December 2010. AFMETS is a real- time surveillance system for service member fatalities that contains demographic information, autopsy reports, death certificates, and official investigation reports with witness interviews. The primary data fields acquired |

| | | |
|---|---|---|
| | | were the cause of death, demo- graphic variables, and activity prior to arrest (eg, running or swimming)." |
| EPIDEMIOLOGY INTERNAL VALIDITY: NUMERATOR AND DENOMINATOR | Low risk | Quote: "Autopsies were available for review on 95% of all SCD/E cases."<br>Quote: "Defense Medical Epidemiology Database person-year (py) data (for the Active Component military population, 2005–2010) was used to calculate the incidence rate of SCD/E overall and stratified by sex, race, age, and branch of service. As this database covers only the full-time Active Component population, and not the Reserve/Guard population, we only report SCD/E incidence rates for the Active Component population."<br>Judgement Comment: Likely slightly overestimates the incidence, as there are 12 deaths from active reserve/guard members, but their units are not considered in the denominator. |
| EPIDEMIOLOGY INTERNAL VALIDITY: MODE OF DATA COLLECTION | Low risk | Judgement Comment: Review of detailed military data. Autopsy, event reports, medical history. |
| EPIDEMIOLOGY FINAL SUMMARY: | Low risk | Judgement Comment: Only areas for concern are applying this data to athletes overall; also the risk of overestimation of the incidence based on decreased denominator size because the number of active duty reserve/guard members are not included in the denominator, but are included in the numerator. |
| EPIDEMIOLOGY EXT VALIDITY: NONRESPONSE BIAS/FOLLOW UP OF COHORT | N/A | |

#### *Solberg 2010*

| | |
|---|---|
| **Methods** | **Study design:** Retrospective cohort study<br>**Retrospective cohort:** Review of Norwegian Cause of death records |
| **Participants** | **Included criteria:** Norwegians between the ages of 17 and 34. Only incidence of sudden death in men was reported, but women were included in the study. The data extracted only included men due to |

| | |
|---|---|
| | the confusion regarding the numerator, denominator and inclusion of men or men vs. women<br>**Excluded criteria:** none listed<br>**Age:** 15-34<br>**Gender:** male<br>**Sport:** combination<br>**Level of Sport:** combination<br>**ECG Screened:** no |
| **Interventions** | Observational |
| **Outcomes** | **SCA:** no<br>**SCD:** yes<br>**Exertional:** yes<br>**Method of ID Cases:** autopsy/death certificate records; Norwegian cause of death registry |
| **Identification** | **Sponsorship source:** None listed<br>**Country:** Norway<br>**Setting:** Athletes under 35<br>**Comments:**<br>**Authors name:** Erik Ekker Solberg, MD, PhD<br>**Institution:** Department of Medicine, Diakonhjemmet Hospital<br>**Email:**<br>**Address:** Department of Medicine Diakonhjemmet Hospital, PO Box 23, Vindern 0319, Oslo, Norway |
| **Notes** | |

Risk of bias table

| **Bias** | **Authors' judgement** | **Support for judgement** |
|---|---|---|
| EPIDEMIOLOGY EXT VALIDITY: REPRESENTATIVE SAMPLE | Unclear risk | Judgement Comment: It appears that this study includes people exercising, as well as competitive athletes. The authors do not give significant detail about this, except to say that there is no accepted definition for a sudden death associated with sport; and when discussing the denominator reference men exercising 3 days/week in Norway. |
| EPIDEMIOLOGY INTERNAL VALIDITY: CASE DEFINITION | Low risk | Quote: "There is no generally accepted method of how cases of sudden cardiovascular death in sports should be registered. In this study, several steps were used to identify the deaths (Fig. 1). The inclusion criteria were diagnoses of sudden death according to the International Classification of Diseases" |

| | | |
|---|---|---|
| | | Quote: "People who suffered cardiac arrest during, or in connection with, physical activity were included."<br>Quote: "If the deaths were sports related, they were further evaluated by inspecting additional information from hospitals and autopsies (Fig. 1)."<br>Quote: "Deaths not related to sports, cases with previously known serious heart disease, and deaths that occurred abroad, were excluded."<br>Judgement Comment: Autopsy and cause of death data. |
| EPIDEMIOLOGY EXT VALIDITY: SAMPLING/SUBJECTS INCLUDED | Low risk | Judgement Comment: Sample appears to be the active population in Norway, including men and women. It is not clear that only competitive athletes are included in this project. Only sport associated deaths. |
| EPIDEMIOLOGY INTERNAL VALIDITY: STUDY PERIOD | Low risk | > 100,000 person years |
| EPIDEMIOLOGY INTERNAL VALIDITY: DATA COLLECTION SOURCE | Low risk | Quote: "In Norway, after examining the deceased, the doctor issues the death certificate, which is further entered in the CoD Registry. This comprises of all deceased persons registered as resident in Norwegian Civil Population Registers at the time of death. Cases with an unknown cause of death are also included. The coverage of the Registry is close to 100% of all deaths as a result of the cooperation with the Norwegian Civil Population Register. Furthermore, Norway is a small country with low migration making a fairly good general overview of the population possible. The CoD Registry collects additional information from autopsy or other postmortem examinations. About 10% of causes of death in the mortality registry are based on the autopsy result. When a sudden unexpected death occurs, a post-mortem investigation is mandatory by Norwegian law."<br>Judgement Comment: 17% of deaths did not have follow up information available. |
| EPIDEMIOLOGY INTERNAL VALIDITY: NUMERATOR AND DENOMINATOR | High risk | Quote: "Of all possible inclusive cases, 17% could not be further classified"<br>Quote: "The number of deceased is a minimum figure; 17% (49 deaths) of the 294 deaths could not |

| | | |
|---|---|---|
| | | be included or excluded because of lack of information."<br>Quote: "Correction of the latter source of error would most probably have increased the incidence. The correct number of deaths would be between 23 and 28 if the proportion of deaths were similar in this (n=49) group as in the total study sample."<br>Quote: "In the middle of the study period, the population of men aged 15–34 years was 649 301. The incidence estimate was based on the information from a national health survey carried out by Statistics, Norway, showing that among these young men, around 50% were physically active; two to three physical exercise periods per week or more. The incidence, thus, was calculated to be one death per 117 238 young physically active males, or 0.9 per 100 000."<br>Judgement Comment: The numerator figure appears relatively accurate, but as the authors point out, has the chance to be around 20% higher based on lack of data on 17% of deaths in young; the Denominator is in doubt as the authors point to the number of eligible men in the study, and appear to calculate the incidence rate with this number, yet the study included men and women. Presumably then making the overall incidence 50% lower? All deaths were included in the calculation (including the one female), it follows that the denominator should then include women and men. The abstract is clear about this, but it is not as clear in the text of the paper how the incidence is calculated. |
| EPIDEMIOLOGY INTERNAL VALIDITY: MODE OF DATA COLLECTION | Low risk | Judgement Comment: same methodology for all cases |
| EPIDEMIOLOGY FINAL SUMMARY: | High risk | Judgement Comment: The risk of bias with the denominator is significant. While much of the data included in this study is very accurate, the confusion about the denominator lends significant risk of bias. Sport, in this study is also unclear. If exercising is considered sport, both the numerator and denominator are affected. While much of the study is well done, well explained, and clear, what are |

| | | |
|---|---|---|
| | | likely the most important parts of the study risk bias in the calculation of the incidence rate. |
| EPIDEMIOLOGY EXT VALIDITY: NONRESPONSE BIAS/FOLLOW UP OF COHORT | N/A | |

#### *Steinvil 2011*

| | |
|---|---|
| **Methods** | **Study design:** Historically controlled trial and retrospective cohort<br>**Study grouping:** Israeli athletes<br>**Retrospective cohort:** Incidence portion of the study is considered retrospective cohort<br>**Historically Controlled trial:** Cohorts from 12 years before and after 1997 law requiring ECG annually and every 4 year exercise stress test for competitive athletes 17-34, and annual exercise stress test for those 35+. Also give incidence rate for 24 year period (retrospective cohort portion). |
| **Participants** | **Included criteria:** Competitive athletes in Israel. The age inclusion is not clear, they appear to determine the denominator based on competitive athletes between the ages of 10 and 40, but deaths included are from 12-44. Deaths included are based on news reports in Israel's two major newspapers which cover 90% readership in country. The determination of SCD is determined by consensus of 3 investigators working on the project.<br>**Excluded criteria:** Non sudden cardiac death<br>**Age:** Unclear. Denominator calculated with ages 10-40. Deaths included in calculations are 12-44 (only one 44 year old out of 10-40 age range).<br>**Gender:** both<br>**Sport:** combination<br>**Level of Sport:** competitive<br>**ECG screened:** portion |
| **Interventions** | **Intervention Characteristics**<br>Intervention group: PPE with ECG, and stress test (1998-2009)<br>Control group: PPE with no ECG, or no PPE (1985-1997) |
| **Outcomes** | **SCA:** yes<br>**SCD:** yes<br>**Exertional:** not described, believed to be all deaths |

| | |
|---|---|
| | **Method of ID Cases:** web/media reports in two national newspapers covering 90% of readership in Israel |
| **Identification** | **Sponsorship source:** none listed<br>**Country:** Israel<br>**Setting:** Competitive athletes in Israel<br>**Comments:** NA<br>**Authors name:** Arie Steinvil, MD, Sami Viskin, MD (last)<br>**Institution:** Department of Internal Medicine "D," Tel-Aviv Sourasky Medical Center and Sackler School of Medicine, Tel Aviv University, Tel-Aviv, Israel<br>**Email:**<br>**Address:** Dr. Sami Viskin, Department of Cardiology, The Tel-Aviv Sourasky Medical Center6 Weizman Street, Tel-Aviv, Israel |
| **Notes** | Both comparative study, and cohort observational study |

Risk of bias table

| **Bias** | **Authors' judgement** | **Support for judgement** |
|---|---|---|
| Random sequence generation (selection bias) | Unclear risk | Judgement Comment: There is no sequence occurring in this historically controlled trial. There is no description of the demographics of groups being compared. |
| Allocation concealment (selection bias) | Unclear risk | Judgement Comment: Again, no formal randomization process undertaken, so not clear that this ROB can be assessed |
| Blinding of participants and personnel (performance bias) | Unclear risk | Judgement Comment: There does appear to be any discussion of blinding those searching for deaths in the newspapers as to what they were looking for/comparing |
| Blinding of outcome assessment (detection bias) | High risk | Judgement Comment: No description of blinding of the professional media researchers looking through the two national papers used for the project. |
| Incomplete outcome data (attrition bias) | High risk | Judgement Comment: Data was extracted exclusively from 2 national newspapers under the assumption that these rare and dramatic events are reported nearly universally. This is addressed by stating that they are only examining "competitive athletes," again supposing that this increases the likelihood of newspaper coverage. However, there is still a significant possibility that the papers did not cover some events or perhaps changed their scope of reporting over the 24 years in question. |

| | | |
|---|---|---|
| Selective reporting (reporting bias) | Low risk | Quote: "All the reports of deaths or dramatic medical events in competitive athletes were brought to the consideration of 3 investigators to determine (by consensus) whether the report could be considered as an athlete's sudden cardiac death or cardiac arrest."<br>Judgement Comment: The study relies on the two national newspapers to report sudden deaths in athletes. It is likely that outcomes were missed, as has been demonstrated in other articles published critiquing the use of media sources as the single identifier of outcomes. The authors also note the newspapers reach 90% of readership in Israel, but it is unclear how this impacts the reporting of sudden death in athletes. |
| Other bias | High risk | Judgement Comment: The comparison of two different time points presents significant risks of bias. The make up of competitive athletes is likely to include more females when comparing 1980s and 90s to the late 90s and 2000s. There is also the issue of likely improved EMS response over time, the distribution and use of AED in public spaces. There have been published articles in European countries reporting on improved rates of survival in out of hospital cardiac arrest; and with athletes in particular in these settings, their survival is better than the general population. These issues would likely confound the comparison of two different time points. This article does use equal lengths of time, to their benefit, to attempt to control for random variation in deaths occurring, or deaths reported on. |
| EPIDEMIOLOGY EXT VALIDITY: REPRESENTATIVE SAMPLE | Low risk | Judgement Comment: Only competitive athletes included in both cases and the denominator |
| EPIDEMIOLOGY INTERNAL VALIDITY: CASE DEFINITION | High risk | Quote: "Overall, 36 incidents of potential sudden death events in competitive athletes were identified by the professional media researchers. Twelve of these incidents were excluded by the investigators because of the following reasons: 6 events were unequivocally the result of trauma (accidental head trauma with brain concussion, intracranial bleeding, |

| | | |
|---|---|---|
| | | or both in 5 and severe chest trauma during a hockey game in 1 athlete); 2 incidents occurred before the assigned study period; and 4 events involved 2 referees, 1 coach, and 1 former athlete. Thus, we identified 24 events of presumed sudden cardiac death or cardiac arrest in athletes."<br>Judgement Comment: "witnessed instantaneous death with futile/successful resuscitation" - case definition problematic because it includes all cause sudden death, potential for many non-cardiac causes, particularly heat related illness given the local climate. Could be over-estimating incidence based on case definition. The only description is that the reporting of sudden death meets a likely definition of SCD in an athlete. There is no confirmatory reporting on the incidence of the deaths. |
| EPIDEMIOLOGY EXT VALIDITY: SAMPLING/SUBJECTS INCLUDED | Low risk | Judgement Comment: The denominator includes Israeli competitive athletes. |
| EPIDEMIOLOGY INTERNAL VALIDITY: STUDY PERIOD | Low risk | > 100,000 person years<br>Judgement Comment: Lengthy (12 years x2) and equivalent periods studied. |
| EPIDEMIOLOGY INTERNAL VALIDITY: DATA COLLECTION SOURCE | High risk | Judgement Comment: Newspaper reports only. No other records, eye witness accounts, autopsy reports or additional data were mentioned. |
| EPIDEMIOLOGY INTERNAL VALIDITY: NUMERATOR AND DENOMINATOR | High risk | Quote: " the number of registered athletes who engaged in competitive sports during 2009 was 45,000. We extrapolated these data to the growth of the Israeli population who were 10 to 40 years of age during the last 24 years as available from the Israeli Central Bureau of Statistics. In addition, because some data suggest that the percentage of the adult population engaging in sportive activities has increased by 50% during the last decade, we repeated our calculations of the number of athletes at risk, assuming a gradual doubling of the percentage of athletes over the 24 years of our study "<br>Judgement Comment: Media identification of cases is known to be a high risk of bias. The Denominator |

| | | |
|---|---|---|
| | | calculation appears to be focused on 10 to 40 years olds, but the cases include one person over 40 years of age. Calling into question the calculation of incidence based on these data. |
| EPIDEMIOLOGY INTERNAL VALIDITY: MODE OF DATA COLLECTION | Low risk | Judgement Comment: Same for all subjects Individually reviewed 24 years of 2 newspapers day-by day. Definite issues with source, but as far as collection goes, it would be unlikely much data was missed. |
| EPIDEMIOLOGY FINAL SUMMARY: | High risk | Judgement Comment: Use of media reports highly biases this paper. It likely under-reports the amount SCD in athletes. |
| EPIDEMIOLOGY EXT VALIDITY: NONRESPONSE BIAS/FOLLOW UP OF COHORT | N/A | |

#### *Toresdahl 2014*

| | |
|---|---|
| **Methods** | **Study design:** Prospective cohort study<br>**Prospective cohort:** Prospective study monitoring high schools with AEDs registered in a national registry for events of SCA on scholastic campuses, in both non-athletes, as well as athletes.<br>**Retrospective cohort:** |
| **Participants** | **Included criteria:** Events of SCA/D occurring on school campus determined by eyewitness and media accounts. The events were reported by participating schools, and details of the deaths were then used to determine the cause of death.<br>**Excluded criteria:** events occurring outside of school campus<br>**Age:** High school age, 14-19<br>**Gender:** both<br>**Sport:** combination<br>**Level of Sport:** scholastic<br>**ECG Screened:** no |
| **Interventions** | Observational |
| **Outcomes** | **SCA:** yes<br>**SCD:** yes<br>**Exertional:** no, all deaths |

| | |
|---|---|
| | **Method of ID Cases:** report from prospectively enrolled schools, with interviews of witnesses, and staff involved; use of web/media report for further details |
| **Identification** | **Sponsorship source:**<br>**Country:** USA<br>**Setting:** High Schools with Automated External Defibrillators participating in Prospective Registry for schools with AED<br>**Comments:**<br>**Authors name:** Brett G Toresdahl, MD, Jonathan A. Drezner, MD (Last)<br>**Institution:** University of Washington<br>**Email:**<br>**Address:** Department of Family Medicine University of Washington, Box 354060Seattle, WA 98195. |
| **Notes** | |

Risk of bias table

| **Bias** | **Authors' judgement** | **Support for judgement** |
|---|---|---|
| EPIDEMIOLOGY EXT VALIDITY: REPRESENTATIVE SAMPLE | Low risk | Quote: "Eighty-six percent of the participating schools were public and 14% private; 48% of schools were rural, 33% suburban, 15% urban, and 4% inner city. At the end of the study, 2045 of 2149 (95%) schools confirmed participation for the entire 2-year study period and the presence or absence of SCA cases on school campus." |
| EPIDEMIOLOGY INTERNAL VALIDITY: CASE DEFINITION | High risk | Quote: "A student athlete was defined as a student participating in any official school sponsored interscholastic team or individual sport requiring regular practice and competition."<br>Quote: "A case was classified as SCA if (1) the person was determined unconscious with the absence of pulse and respirations, (2) cardiopulmonary resuscitation was provided or an AED deployed a shock, (3) a primary cardiac etiology was found, or (4) a traumatic blow to the chest occurred and was consistent with commotio cordis. Only those cases occurring on school campus during the study period were included."<br>Judgement Comment: Eyewitness accounts and media accounts without any autopsy or medical reports for deaths, may overestimate the number of events of SCA. |

| | | |
|---|---|---|
| EPIDEMIOLOGY EXT VALIDITY: SAMPLING/SUBJECTS INCLUDED | Low risk | Quote: "The incidence of SCA in student athletes was 1.14 per 100,000 (95% CI 0.68–1.80) and was higher in male student athletes (1.78 per 100,000; 95% CI 0.99–2.81) than in female student athletes (0.31 per 100,000; 95% CI 0.04–1.11)."<br>Judgement Comment: Large cohort of high schools with broad differences in location, public vs private, urban vs rural. |
| EPIDEMIOLOGY INTERNAL VALIDITY: STUDY PERIOD | Low risk | > 100,000 person years |
| EPIDEMIOLOGY INTERNAL VALIDITY: DATA COLLECTION SOURCE | High risk | Quote: "When a potential case of SCA was reported, the details of the case were reviewed over the phone with the school representative or another staff member who had witnessed the event or was involved in the emergency response. In some cases, media reports were used to clarify case demographics or supplement the information provided."<br>Judgement Comment: No medical data, or autopsy data was included in the study. The decision to report as SCA was determined by review of details of eyewitnesses, occasionally with media reports adding extra information. It is possible this could have led to misclassification of some of the deaths, overestimating the number of SCA |
| EPIDEMIOLOGY INTERNAL VALIDITY: NUMERATOR AND DENOMINATOR | Low risk | Quote: "total of 2149 US high schools distributed throughout all 50 states participated in the study. The mean number of students and student athletes per school as reported by the school representative were 963 and 367, respectively. For the 2-year study period, this provided more than 4.1 million total student-years and more than 1.5 million student athlete- years of surveillance."<br>Quote: "One hundred twenty-nine potential cases of SCA were reported by school representatives. Fifty-nine cases met the inclusion criteria."<br>Quote: "Twenty-six cases of SCA occurred in students."<br>Quote: "Eighteen (69%) cases occurred in student athletes, and all of them were associated with physical activity." |

| | | |
|---|---|---|
| | | Quote: "This study underestimates the true incidence of SCA in high school students and student athletes because it accounts for only those cases that occurred on high school campuses. This study does not account for cases that occurred outside the school campus, at home, on weekends, or when school was not in session. In prior studies, up to 20% of the cases of SCD in young athletes included in the incidence calculations occurred while away from school, at rest, or during sleep."<br>Judgement Comment: Only including events at school removes a substantial portion of time. This includes time away on weekends, breaks, nights, etc. Some studies suggest up to 20% of SCD in athletes occurs outside of exertion, and this study would miss these events. The authors however, are clear about their methods, and limitations, making the risk of bias low about this proportion of missed cases. In the end, this paper likely underestimates the incidence of SCA in athletes. |
| EPIDEMIOLOGY INTERNAL VALIDITY: MODE OF DATA COLLECTION | Low risk | Judgement Comment: Data collected in same manner for all student athletes |
| EPIDEMIOLOGY FINAL SUMMARY: | Moderate risk | Judgement Comment: Overall low risk of bias. The authors understand that their methodology may underestimate SCA in athletes because only the events at schools are included. They do not acknowledge, however, that their methodology for determining cause of death may bias the result, using only eyewitness and media accounts rather than including medical data and autopsy information. With this in mind, the study is well done and well reported and is determined to be a moderate risk. |
| EPIDEMIOLOGY EXT VALIDITY: NONRESPONSE BIAS/FOLLOW UP OF COHORT | N/A | |

#### *Young 1999*

| | |
|---|---|
| **Methods** | **Study design:** Retrospective cohort study<br>**Retrospective cohort:** Review of autopsy, and forensic pathology data on Aboriginal sports men in the Northern Territory. Only data on Aussie Rules football extracted as this came with incidence data. |
| **Participants** | **Included criteria:** Aboriginal men playing sport ages 15-37. Incidence data from those playing Aussie Rules Football only.<br>**Excluded criteria:** none listed<br>**Age:** 15-37<br>**Gender:** male<br>**Sport:** Australian football<br>**Level of Sport:** competitive<br>**ECG Screened:** no |
| **Interventions** | Observational |
| **Outcomes** | **SCA:** yes<br>**SCD:** yes<br>**Exertional:** yes<br>**Method of ID Cases:** autopsy reports |
| **Identification** | **Sponsorship source:** Sports Program, and Australian Sports Commission<br>**Country:** Australia<br>**Setting:** Northern Territory Aboriginal Sportsmen<br>**Comments:**<br>**Authors name:** Mark C Young, MB, BS<br>**Institution:** University of Canberra<br>**Email:**<br>**Address:** Australian Institute of Sport Leverrier Crescent, Bruce, ACT 2617 |
| **Notes** | |

Risk of bias table

| Bias | Authors' judgement | Support for judgement |
|---|---|---|
| EPIDEMIOLOGY EXT VALIDITY: REPRESENTATIVE SAMPLE | Low risk | Judgement Comment: Appears to be representative of young Aboriginal sportsmen in the Northern Territory. |
| EPIDEMIOLOGY INTERNAL VALIDITY: CASE DEFINITION | Low risk | Quote: "Sudden cardiac death due to atherosclerotic IHD was defined as unexpected, atraumatic death due to cardiac arrest within an hour of previously normal |

| | | |
|---|---|---|
| | | health, when autopsy examination showed a critical narrowing (~70%) in a coronary artery."<br>Quote: "A young competitive sportsman was defined as a male aged 15-37 years who participated in an organised team or individual sport in which regular com-petition is a component and in which a premium is placed on achievement."<br>Judgement Comment: Causes of death noted on autopsy report used. Deaths within 1 hour of playing sport. |
| EPIDEMIOLOGY EXT VALIDITY: SAMPLING/SUBJECTS INCLUDED | Low risk | Quote: "Six of the eight deaths were associated with Australian football (which is played only in the wet season in the Top End) and one each was associated with soccer and touch football."<br>Judgement Comment: Deaths with reported incidence only include Aussie Rules football, but 2 other deaths described without incidence. All deaths in Aboriginal men, while playing sport was sought. Only data on Aussie football was available for denominator. Appears to be low risk. |
| EPIDEMIOLOGY INTERNAL VALIDITY: STUDY PERIOD | High risk | Judgement Comment: only between 25-30,000 athlete years. |
| EPIDEMIOLOGY INTERNAL VALIDITY: DATA COLLECTION SOURCE | Low risk | Quote: "Details of these deaths were provided by the Australian Bureau of Statistics (ABS) from information collected initially by the NT Registrar of Births, Deaths and Marriages. Permission to view autopsy records was granted by the NT Coroner. (Under the NT Coroners Act, all unexpected deaths are reported to the Coroners Office, and autopsies are usually performed to establish the cause of death.) Deaths identified by the autopsy reports as being related to sporting participation were included as cases."<br>Quote: "included as cases. Secondly, a computer search was undertaken of NT forensic pathology records for the period 1991-1996 to cross-check ABS figures and provide information for 1996. Search terms were "football", "footy", "sport", "exertion", "rugby", "basketball", "athletics", "running" and "swimming". All deaths were confirmed as being caused by IHD from the autopsy findings. The Aboriginality of the deceased was also determined from the autopsy records, as the NT death registration system has provided for Aboriginal |

| | | |
|---|---|---|
| | | identification only since 1988. Analyses Participation was estimated for"<br>Judgement Comment: This appears low. The authors do comment there may be cases that are missed, but they appear to feel good about the numerator/cases identified. |
| EPIDEMIOLOGY INTERNAL VALIDITY: NUMERATOR AND DENOMINATOR | High risk | Quote: "were identified by two methods. Firstly, deaths of males aged 15-37 years registered in the NT in the period 1982-1995 with the cause of death coded as IHD (ICD-9 codes 410.0-414.9 2 ) were considered."<br>Quote: "identification only since 1988. Analyses: Participation was estimated for Australian football from the number of teams in each league and the likely pro- portions of Aboriginal and non-Aboriginal players, provided by the NT Football Development Foundation. Each team was assumed to have 25 players. As team numbers were available only for 1996, the number of player- years for the period 1982-1996 was estimated in two ways. Firstly, the likely maximum number of player-years was estimated by assuming that the 1996 participation applied for each year in the period. Secondly, the number of player- years was extrapolated from the total NT Aboriginal population calculated from ABS data. This method is likely to underestimate total player-years."<br>Quote: "We identified eight sport-related sudden cardiac deaths due to IHD among Aboriginal sportsmen aged 15-37 years in the NT."<br>Quote: "The NT Football Development Foundation reported that 110 Australian rules football teams played in the 1996-1997 season, giving an estimated 2750 players - 2095 Aboriginal and 655 non-Aboriginal. Of the Aboriginal players, 578 played in an urban competition and 1421 (74 %) played in a com- munity league, with a small percentage participating in both. Based on the two methods of estimation, Aboriginal Australian footballers had between 25050 and 31 425 player-years in the period 1982-1996 (inclusive). From these data, the incidence of sudden cardiac death due to IHD in young NT Aboriginal Australian footballers was 1 per 4175-5240 player-years (19-24 per 100000 player-years)."<br>Quote: "There is clearly some doubt about the precise incidence of these deaths among Aboriginal footballers. Firstly, the number of cases might have been under- |

| | | |
|---|---|---|
| | | estimated if deaths were not reported to the Coroner or if relevant cases were missed because files could not be located. Secondly, calculation of the total number of player-years was based on estimated participation rates, and no specific information was available before 1996. However, there can be little doubt that the incidence of IHD-related sudden cardiac death among Australian footballers is very much greater for Aboriginal players in the NT than for players in Victoria."<br>Judgement Comment: Authors point out the likely undercount of those playing football in this data. This is important with a high incidence being reported. They also suggest the possibility of undercount of cases. |
| EPIDEMIOLOGY INTERNAL VALIDITY: MODE OF DATA COLLECTION | Low risk | Judgement Comment: All data collected from autopsy report and forensic pathology official data. |
| EPIDEMIOLOGY FINAL SUMMARY: | Moderate Risk | Judgement Comment: Article appears to primarily have low risk of bias; However, the authors note that there may be an undercount in the denominator due to the risk of undercount of intended subjects likely playing Aussie football. This is critical to the understanding of the article, and puts the incidence count number in question, and have decided to categorize as moderate risk for this reason. |
| EPIDEMIOLOGY EXT VALIDITY: NONRESPONSE BIAS/FOLLOW UP OF COHORT | N/A | |

#### Appendix F: Notable Excluded studies

A 1995 article[7] focusing on high school and collegiate athletes in the United States was excluded as the information included in the database reported on, was also used for Maron, et. al., (2016) which presented an overall larger data set. For this reason, we elected not to include this article in our review.

A 2014 article[8] on university level athletes in the United States by Maron, et. al. was excluded for two reasons. Primarily it was a subgroup of the larger 2016 article published by the same author. The time periods reported overlapped substantially with Harmon, et. al. (2015). Both papers used a database from the National Collegiate Athletic Association, while Harmon, et. al. drew from a separate database as well, and reported greater detail for subgroup analysis and extraction. For these reasons, Harmon, et. al. (2015) was included.

### Appendix G: Subgroup Analysis

**Incidence of SCA/D by age**

**Age <25**

High quality studies reporting on this subject are including in the primary analysis as the competitive athletes. Only one low ROB study reported SCA. There was substantial heterogeneity overall in the studies presenting information on this subgroup, with the large majority being either moderate or high risk of bias. Most of the risk of bias results from the uncertainty in collecting this data, and likely undercounts based on data collection of cases. As is discussed in the main text, the zero count studies presenting estimates for SCD with small denominators should be assessed with caution.

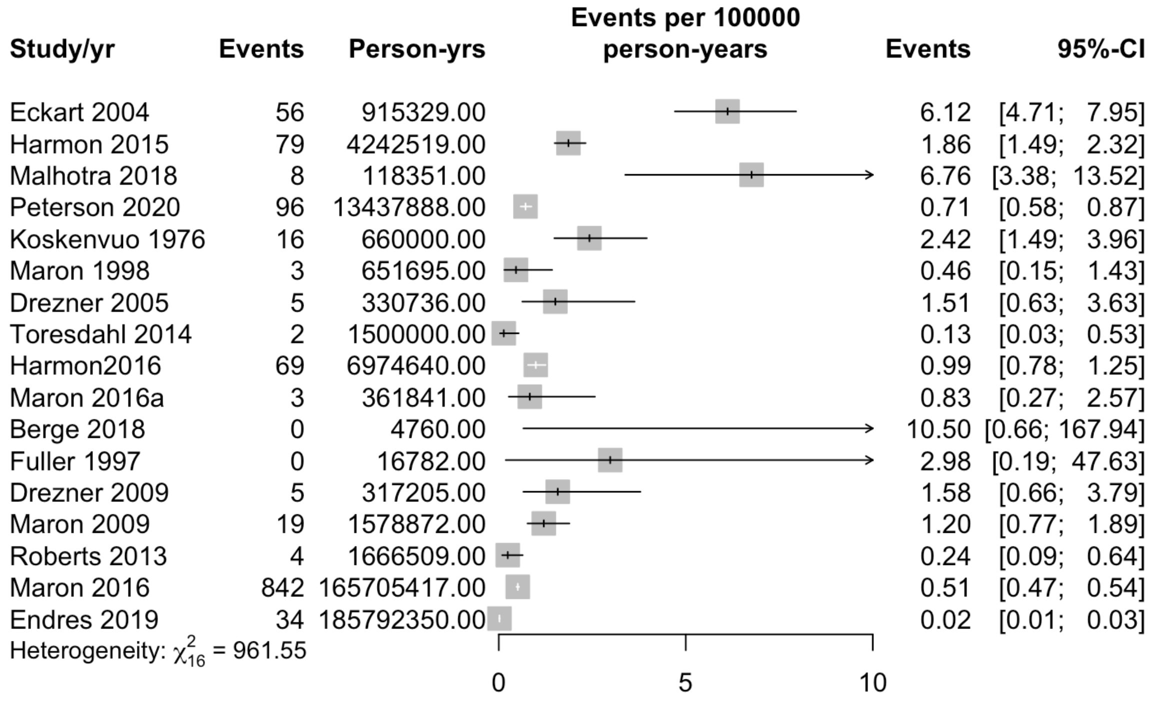

**Figure S1:** Incidence of SCD in athletes ≤25 years of age in included studies

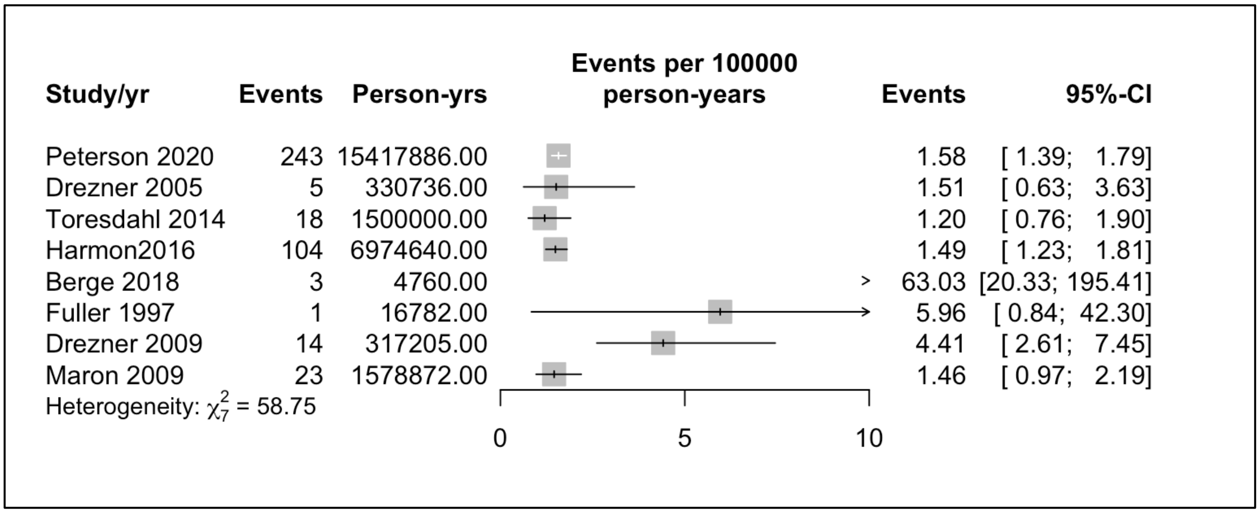

**Figure S2:** Incidence of SCA in athletes <u><</u>25 years of age in included studies

##### Age>25

Only one study included extractable data on those between 25 and 40 years of age. Eckart, et. al. (2004) included this data, and the incidence rate was calculated at 14.4 episodes of SCD per 100,000 (95%CI: 7.2-28.79) person years in a population of military recruits. Other included studies reported incidence rates on populations that included those in this age rage, but no other studies specifically reported raw data, nor incidence rates on those specifically in this age group. No included studies included data on SCA only in this age group.

##### Incidence of SCA/D sex

##### Male

16 articles reported extractable data on the incidence of SCD in males, with only the top three in figure S3 at low risk of bias. The numbers presented here overall do appear on visual inspection to be elevated when compared to female athletes/military.

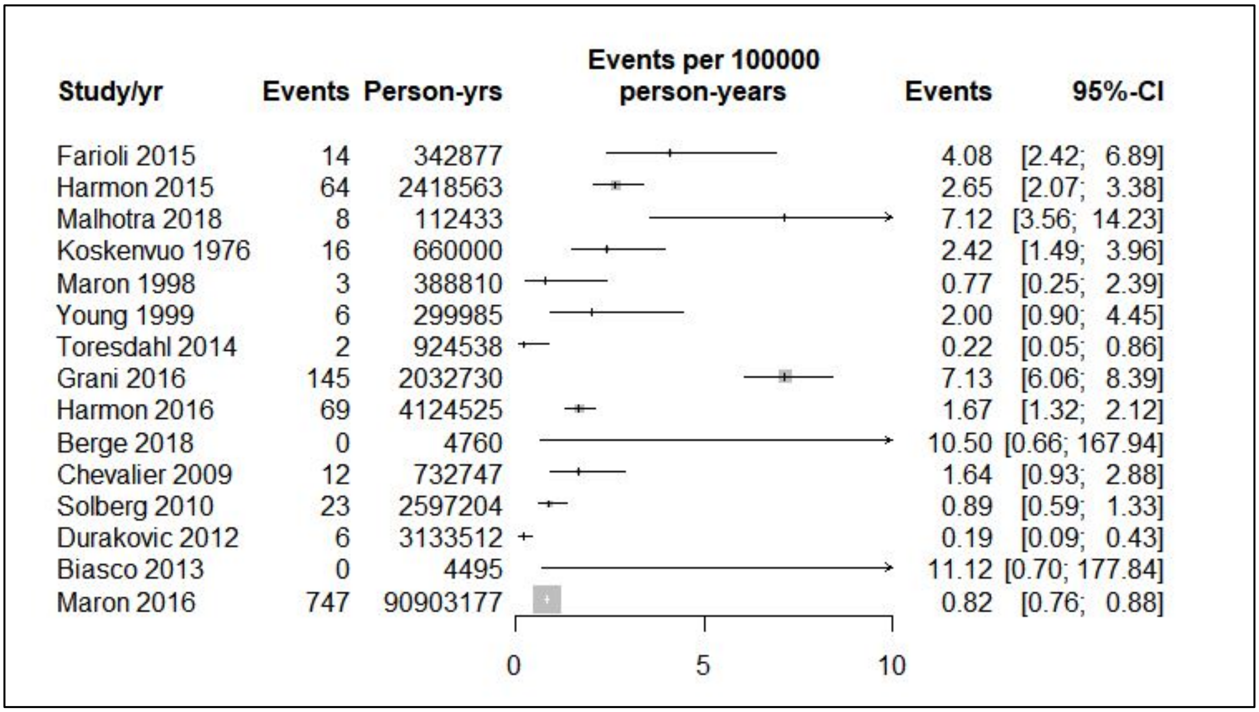

**Figure S3:** Incidence of SCD in male athletes/military in included studies

Three studies reported incidence rates of SCA in males, and this data was not meta-analyzed. Peterson, et. al. (2020) is the lone article with low ROB, including high school and collegiate athletes.  Two separate reports with moderate ROB on U.S. high school athletes [9,10] found similar rates, with Toresdahl reporting 1.73 (95% CI: 1.06,2.82) and Harmon reporting 2.23 (95%CI: 1.82,2.74) per 100,000 person years. It is notable in this case, that the Toresdahl article is a prospective database from participating high schools, while the Harmon study uses a database collected from web and media reports.  The third study reporting a rate of SCA is male professional soccer players in Norway[11], and reported a rate of 63.03 (95%CI: 20.33, 195.41) per 100,000 person years.

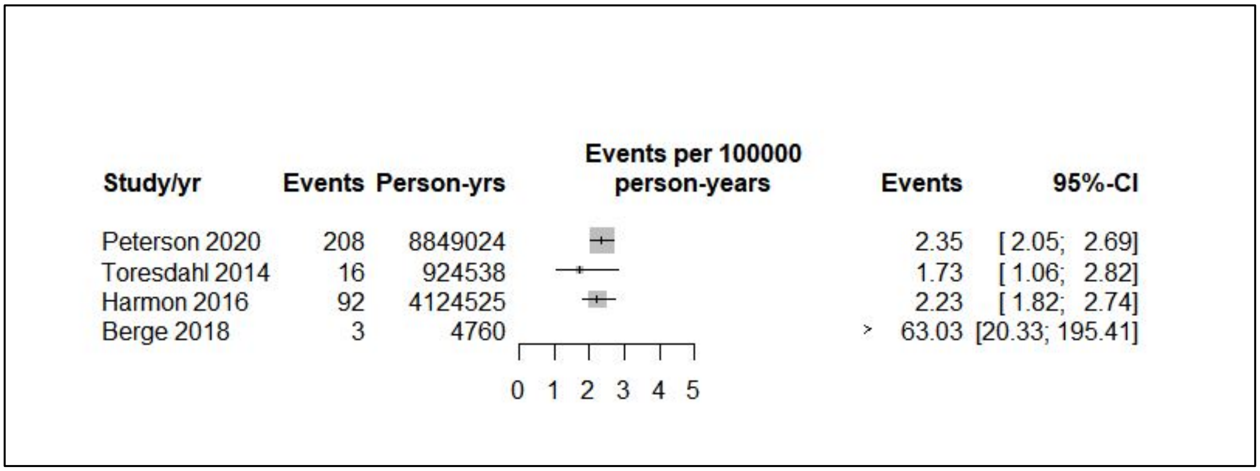

**Figure S4:** Included studies reporting SCA in male athletes

#### Female

7 studies reported rates of SCD in female athletes, the initial two in figure S5 with low ROB. As above, the zero count studies should be assessed with caution. All included studies were of athletes, with no military populations. 5 of the included studies reported no incidents of SCD in women within their data set [10,12–15].

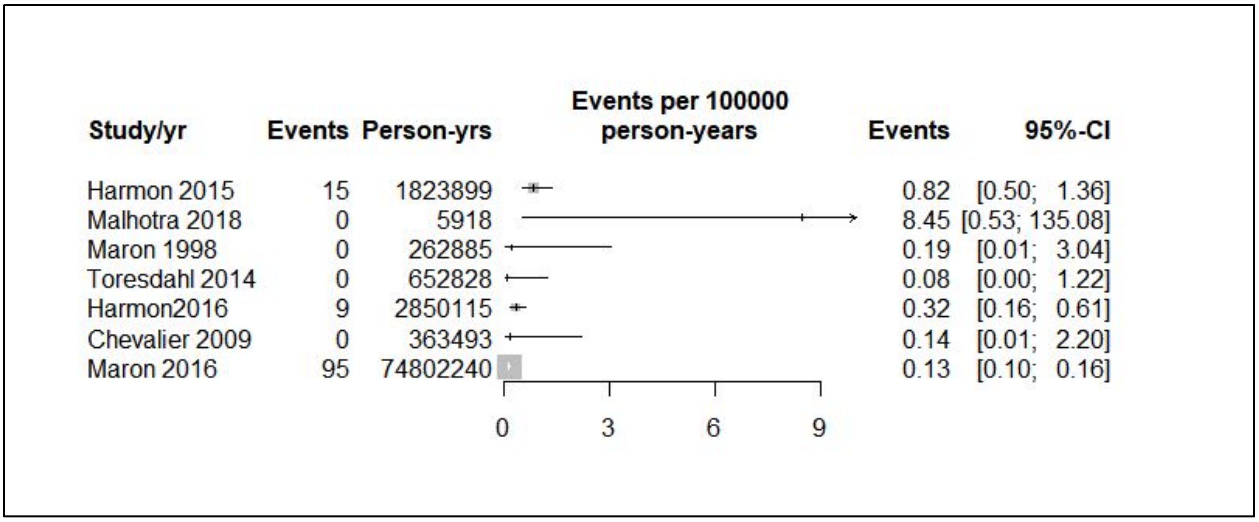

**Figure S5**: SCD in female athletes reported by included studies

Only three articles reported extractable data on SCA in female athletes. Peterson, et. al. (2020) again is the lone low ROB study. On visual inspection as above, the numbers with all three articles presented are lower than those presented in male athletes.

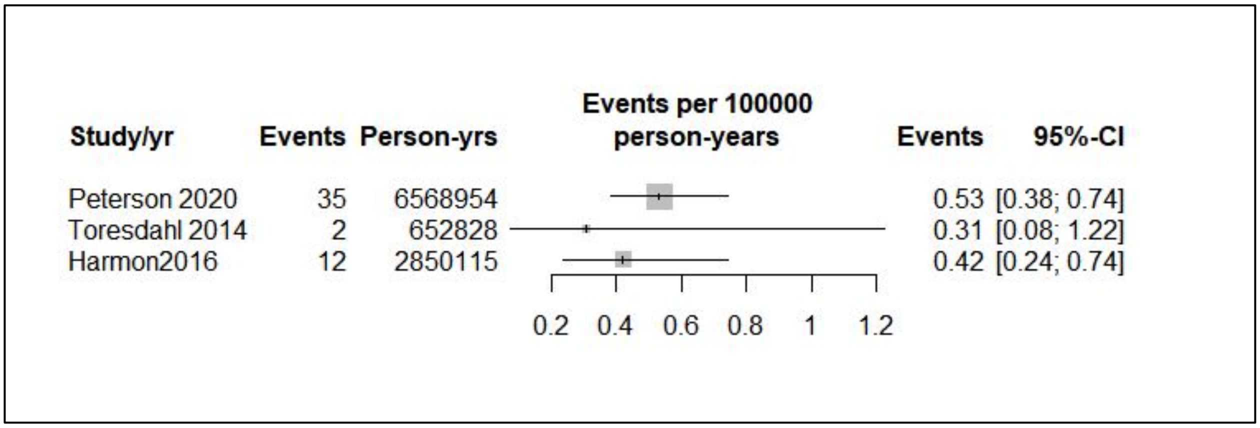

**Figure S6**: Two included articles reporting SCA in female athletes

##### Incidence of SCA/D by athlete level

###### Elite

7 articles reported data on professional [11,15–18] or National Collegiate Athletic Association (NCAA) division one university athletes [19,20]. There are two low ROB articles in reporting on this group, both included in the primary meta-analyses, with Harmon, et. al. (2015) a subgroup of NCAA division one athletes from their overall study. Visual inspection of the results in this category is shows generally elevated incidence rates when compared with athletes in other categories, but as in all cases, estimates made from studies with zero counts should be interpreted with caution. It is notable that in the case of the professional athlete entries, nearly all of them are of male athletes, or a large proportion of the subjects are males.

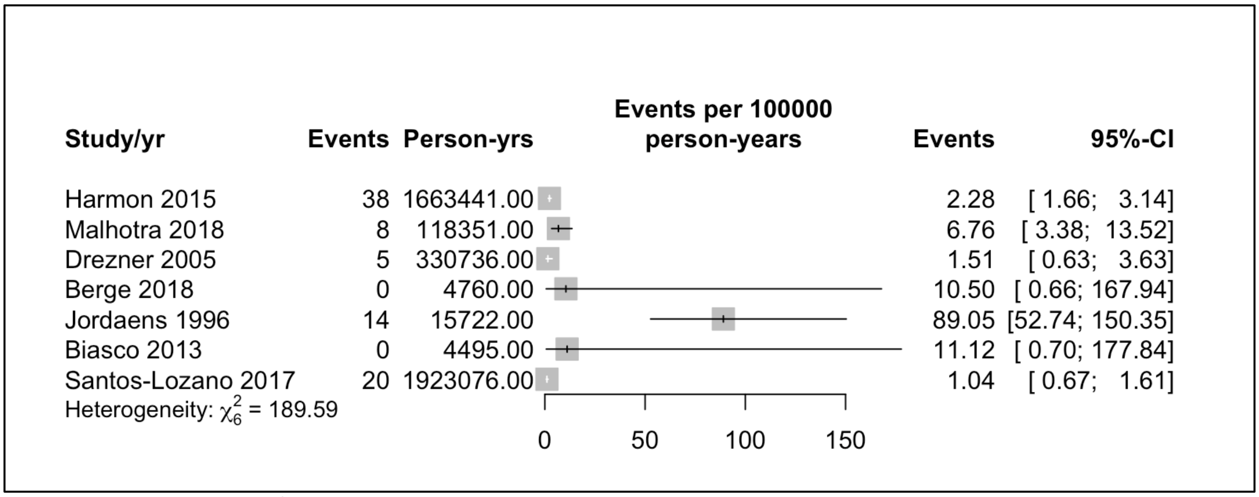

**Figure S7**: Incidence of SCD in Elite athletes in included studies

Only two included studies reported on events of SCA in elite athletes. Drezner, et. al. (2005) reported 5 cases, all of whom died as detailed above, with a rate of 1.51 per 100,000 person years (95%CI: 0.63,3.63). While Berge, et. al. (2018) reported 3 episodes of SCA, none of whom died. Their calculated rate is 63.03 (95%CI: 20.33, 195.41) per 100,000 person years.

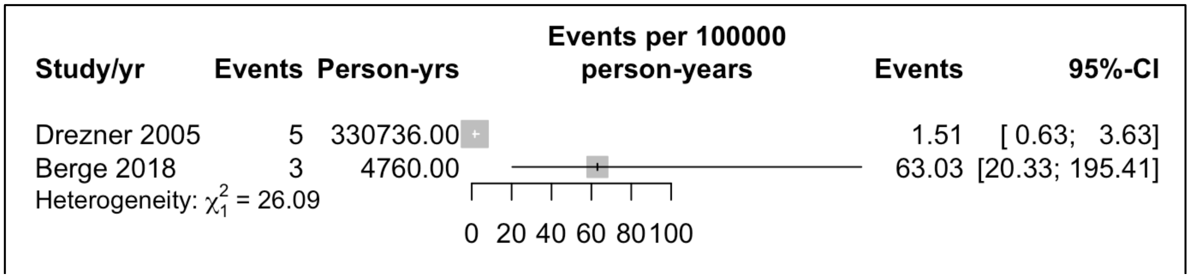

**Figure S8:** Included studies reporting SCA in elite athletes

**University/College**

Two included studies provided data on university athletes at all levels, which was not meta-analyzed. Boden, et. al. (2013) is a high ROB study, reporting only on American football players, reporting a rate of 0.89 per 100,000 person years (95%CI: 0.53, 1.51). Harmon, et. al. (2015) is a low ROB study, reported on a large population of college athletes over a 10 year period. The reported rate for this cohort is report 1.86 (95% CI:1.49,2.32) per 100,000 person years. Only Peterson, et. al. (2020), a low ROB study, reported on SCA in university level athletes reporting a rate of 1.97 [95%CI: 1.44; 2.7] in a study reporting 39 events over 1,979,938 athlete years.

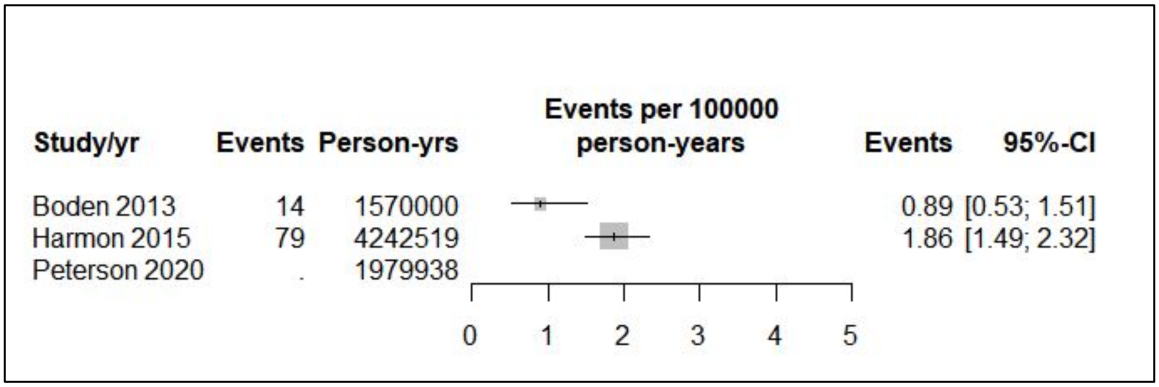

**Figure S9:** Included studies reporting SCD in university athletes

**Scholastic**

The 10 included articles reporting rates of SCD in scholastic athletes, considered athletes participating in sport prior to graduation from high school, include 8 articles which focus solely on athletes in middle or high school[9,10,12,13,21–24]. The other two providing subsets of data on scholastic athletes, both of which focus on American football athletes, and are all male[25,26]. Only Peterson, et. al (2020) was judged to have low ROB, with the remainder having moderate or high risk of bias, but visual inspection of their estimates suggest relatively low incidence, with the only point estimate over 2.00 coming from a zero count study which should be interpreted with caution.

As noted in the text of Peterson, et. al. (2020), there is reason to believe that events in this population may be subject to undercount, when compared to more elite athletes.  While the numbers here are mostly under 1.00 per 100,000 athlete years, there is reason to believe the true number may be somewhat higher compared to this, but there is not good evidence to suspect that it is much higher.

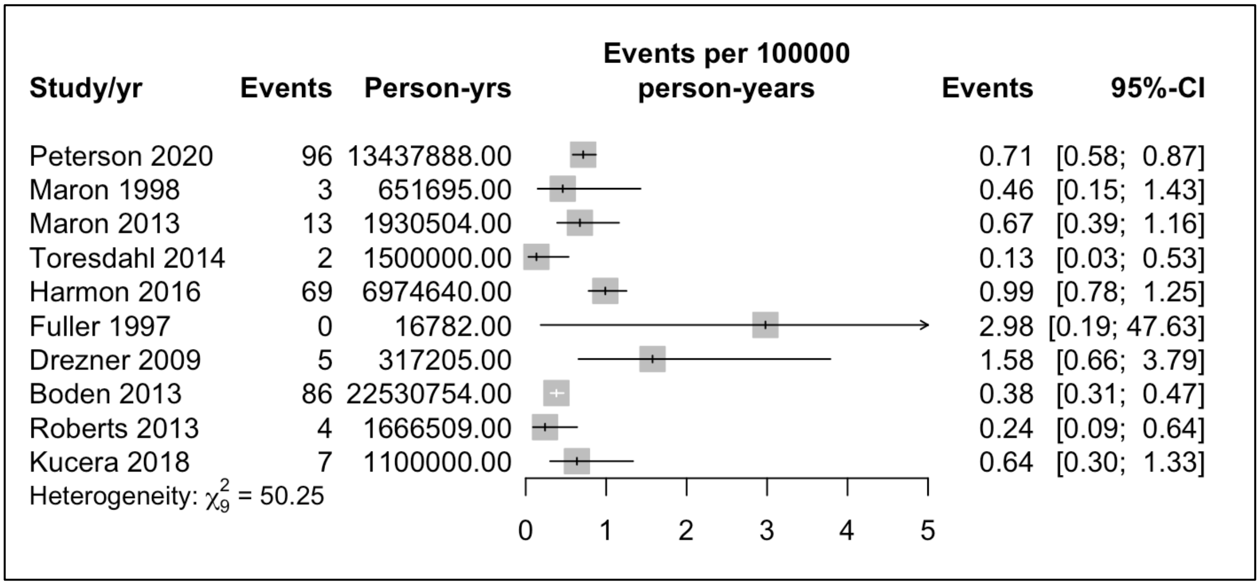

**Figure S10**: Incidence of SCD in scholastic athletes in included studies

Incidence of SCA in scholastic athletes is available below, and all five included studies present data on a range of scholastic athletes, that includes both male and female subjects.  Peterson, et. al. (2020) is the only included study judged to be low ROB.  The three studies with numbers under two are large studies, with more reliable estimates in our opinion. It is notable that both Toresdahl, et. al. (2014), and Harmon et. al., (2016) were judged moderate ROB due to their likely undercount of events.  Fuller, et. al. (1997) is a very small study of a group of athletes screened by a practice in NV, USA and followed for a short period of time; while Drezner, et. al. (2009) is a survey study. Both of these have elevated estimates, with reasons for concern about their results.

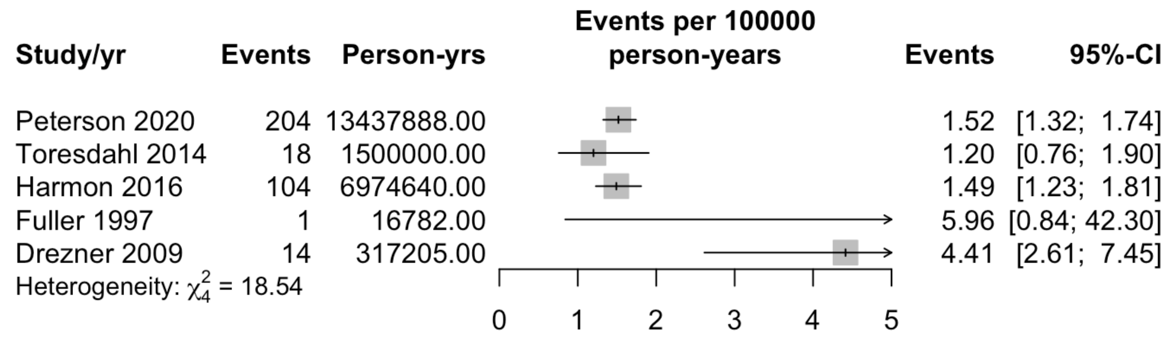

**Figure S11:** Incidence of SCA in scholastic athletes in included studies

**Competitive**

The 12 studies included in this category reported their subjects as competitive athletes. This includes those participating in organized sport which could span any category in elite, university, scholastic, or club athlete, it does exclude recreational athletes. 3 studies reported rates only in one sport, with Maurice, et. al. (2018) reporting events in rugby, Young, et. al. (1999) in Australian football, both of these studies either exclusively, or primarily in men, and Maron, et. al. (2009) in lacrosse. The low ROB studies included here were meta-analyzed in the primary text.

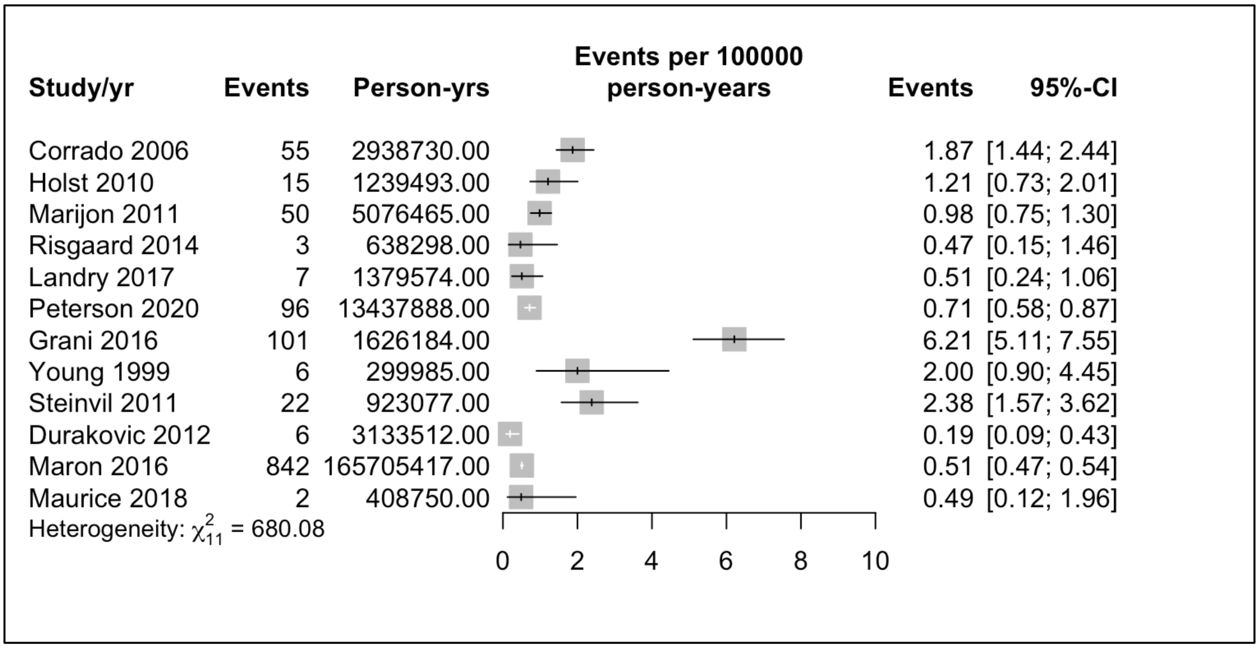

**Figure S12**: Incidence of SCD in competitive athletes in included studies

5 studies reported events of SCA in competitive athletes, with two population based data [27,28], and two studies single sports studies, one including both genders [29], one of which is primarily men [30]. Maron (2009) is a subset of the larger data set from the same author group, but includes data on SCA, which is where this study is included. Low ROB studies here are discussed in main text.

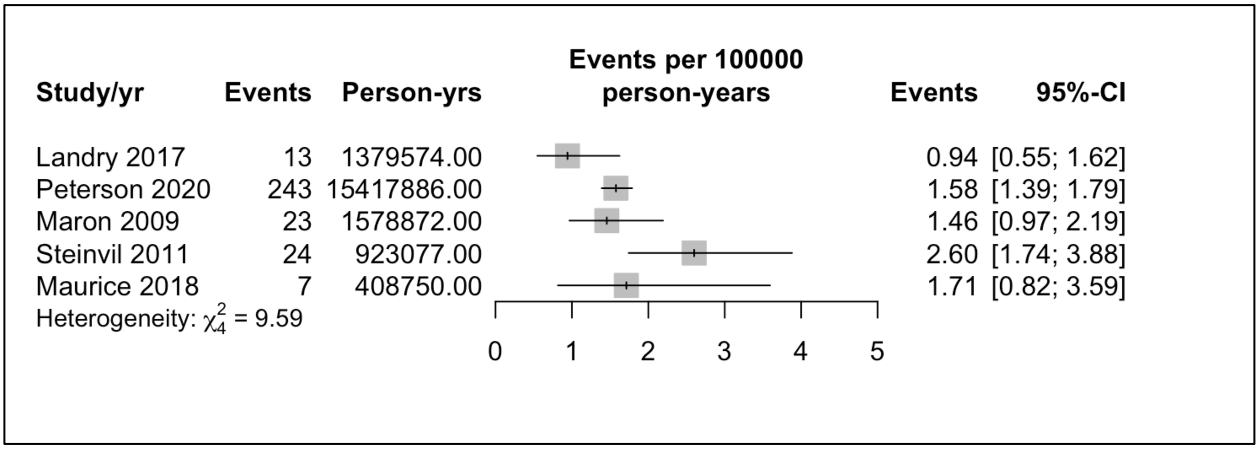

**Figure S13:** Incidence of SCA in competitive athletes in included studies

##### Recreational

Only one included article reported specifically on the incidence rate of recreational athletes as a subset of their full data collection. Grani, et. al. (2016) reported 44 SCDs in a sample of 406546, for a rate of 6.21 events per 100,000 person years (95%CI: 5.11,7.55). No articles reported on SCA in recreational athletes.

##### Military

Reporting on military studies is discussed in the main text.

##### Incidence of SCA/D by race

Two studies included subgroup analysis of white athletes[20,31], and these estimates are similar, and lower than those seen in the other reported races. Harmon, et. al. (2015) also reports estimates for black and Hispanic NCAA athletes; while Eckart reports black, and non-black estimates from a military study. In the same manner, Maron, et. al. (2016) reports white, and non-white categories, the non-white category includes 350 cases from black athletes, and 40 cases from 'other' category. Peterson, et. al, (2020) gives detail on black men, but not an overall black population, and is not discussed listed here.

| Author/year | Race | Incidence per 100,000 person-years | 95% Confidence Interval |
|---|---|---|---|
| Harmon 2015 | White | 1.46 | 1.09, 1.96 |
| Maron 2016 | White | 1.65 | 1.50,1.81 |
| Eckart 2004 | Non-black | 5.30 | 3.90, 7.20 |
| Eckart 2004 | Black | 12.00 | 7.98, 18.06 |
| Harmon 2015 | Black | 4.65 | 3.25, 6.66 |
| Harmon 2015 | Hispanic | 1.78 | 0.57, 5.51 |
| Maron 2016 | Non-white | 7.83 | 7.09, 8.65 |

**Table S1:** Included studies with SCD by race as categorized by authors

#### Incidence of SCA/D by sport

##### Soccer

5 studies reported data on SCD in soccer athletes.  4 of 5 were on professional soccer players [11,15,17,18], and only Malhotra, et. al. (2018) included female athletes, albeit less than 10% of the cohort. It is notable when considering the Malhotra, et. al. (2018) data, that 2 of the 8 athletes suffering SCD were identified as at risk during screening, and advised to withdraw from sport, but chose not do so.  Soccer is discussed in more detail in primary test.

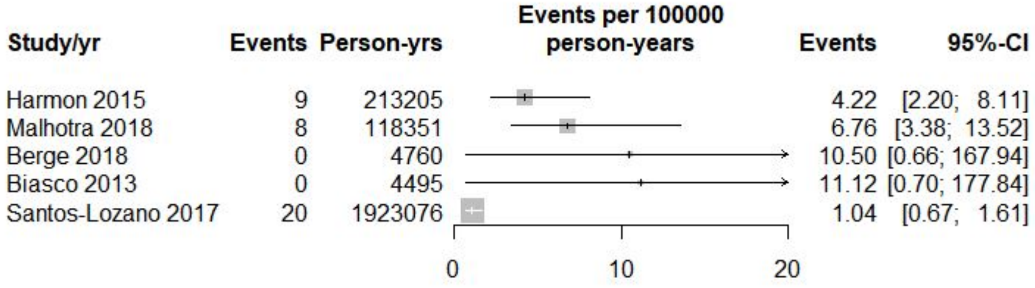

**Figure S14:** Incidence of soccer athlete SCD in included studies

Two studies reported a rate of SCA in soccer athlete, Berge et. al. (2018) report on Norwegian professional males, reporting a rate of 63.0 (95%CI: 20.33, 194.41), and Peterson, at. al. (2020) reporting a rate in high school and collegiate male and female athletes

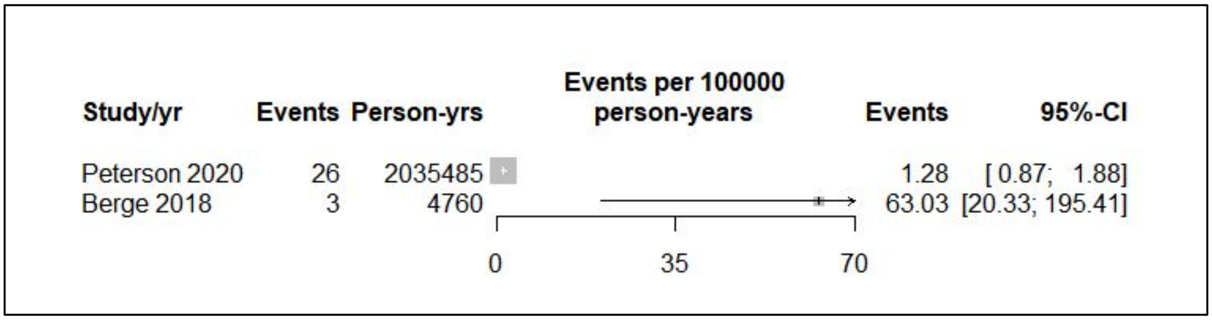

**Figure S15:** Incidence of soccer athlete SCA in included studies

**American Football**

4 included studies reported data on SCD in American football athletes. Harmon, et. al. (2020) was the only study judged low ROB, showing a much higher rate than other included studies.

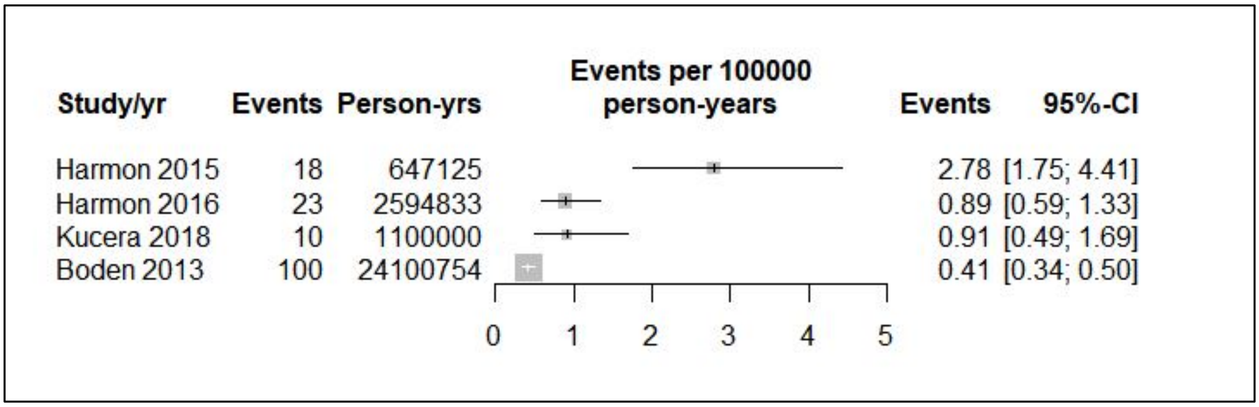

**Figure S16** Incidence of SCD in American football players in included studies

Peterson, et. al. (2020) is the only low ROB study reporting SCA, including high school and collegiate players. Harmon, et. al. (2016) reported a lower rate, and this was in high school players only.

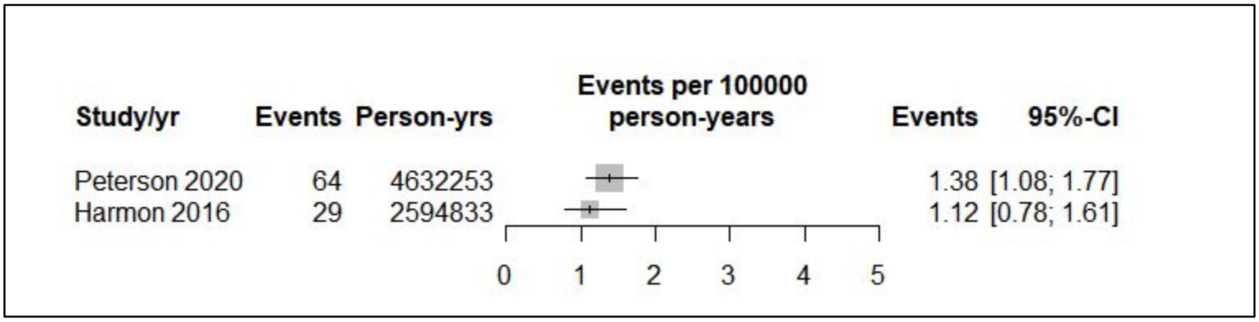

**Figure S17** Incidence of SCA in American football players in included studies

The remaining included studies did not produce enough data to allow meta-analysis on further individual sports. Table 2 lists the sports reported from included studies. The majority of the data included is drawn from Harmon, et. al. (2015)'s report on NCAA athletes. It is notable that only NCAA sports with more than one death had data published in this article, allowing this incidence calculation. Young, et. al. (1999) reports only on male Aboriginal Australian Rules football players.

| Author/Year | Sport | SCA per 100,000 | 95% Confidence Interval | SCD per 100,000 | 95% Confidence Interval |
|---|---|---|---|---|---|
| Young 1999 | Australian Football (M) | NA | NA | 2.00 | 0.90, 4.45 |
| Harmon 2015 | Baseball (M) | NA | NA | 2.00 | 0.90, 4.45 |
| Peterson 2020 | Baseball (M) | 0.46 | 0.24, 0.89 | NA | NA |
| Harmon 2015 | Basketball (M) | NA | NA | 11.14 | 7.10, 17.46 |
| Peterson 2020 | Basketball (M) | 2.84 | 2.22,3.63 | NA | NA |
| Drezner 2014 | Basketball (M) | NA | NA | 4.70 | 2.11, 10.46 |
| Harmon 2015 | Basketball (F) | NA | NA | 1.30 | 0.32, 5.12 |
| Jordaens 1996 | Cycling (M) | NA | NA | 89.05 | 52.74, 150.35 |
| Peterson 2020 | Ice Hockey (M) | 4.25 | 1.91, 9.45 | NA | NA |
| Harmon 2015 | Lacrosse (M) | NA | NA | 2.18 | 0.55, 8.72 |

| | | | | | |
|---|---|---|---|---|---|
| Maron 2009 | Lacrosse | 1.46 | 0.97, 2.19 | 1.23 | 0.77, 1.89 |
| Maurice 2018 | Rugby (M) | 1.71 | 0.82, 3.60 | 0.49 | 0.122, 1.20 |
| Harmon 2015 | Track (M)/Cross Country (M/F) | NA | NA | 1.57 | 0.78, 3.13 |
| Peterson 2020 | Track | 0.38 | 0.22, 0.89 | NA | NA |
| Harmon 2015 | Swimming | NA | NA | 1.99 | 0.75, 5.31 |
| Harmon 2015 | Volleyball (F) | NA | NA | 2.03 | 0.66, 6.30 |
| Peterson 2020 | Volleyball | 0.50 | 0.21, 1.20 | NA | NA |

**Table S2:** Individual sports extracted from included studies.
M indicates only male athletes included in analysis; F indicates only female athletes included in analysis.

### Appendix H: Assessment of publication bias

Visual assessment of publication bias with a funnel plot was performed . Given the small number of included studies for objective two, a funnel plot analysis was not performed. The funnel plot does show asymmetry with studies with lower standard error showing larger incidence rates to the right of the funnel, with the lower incidence rate studies to the left of the plot. This argues against the idea of publication bias, with lower incidence rates in smaller studies. The significant between study heterogeneity noted in the analyses above appears at least as likely to be the source of the funnel plot asymmetry, as any publication bias [4,32] may be in this case.

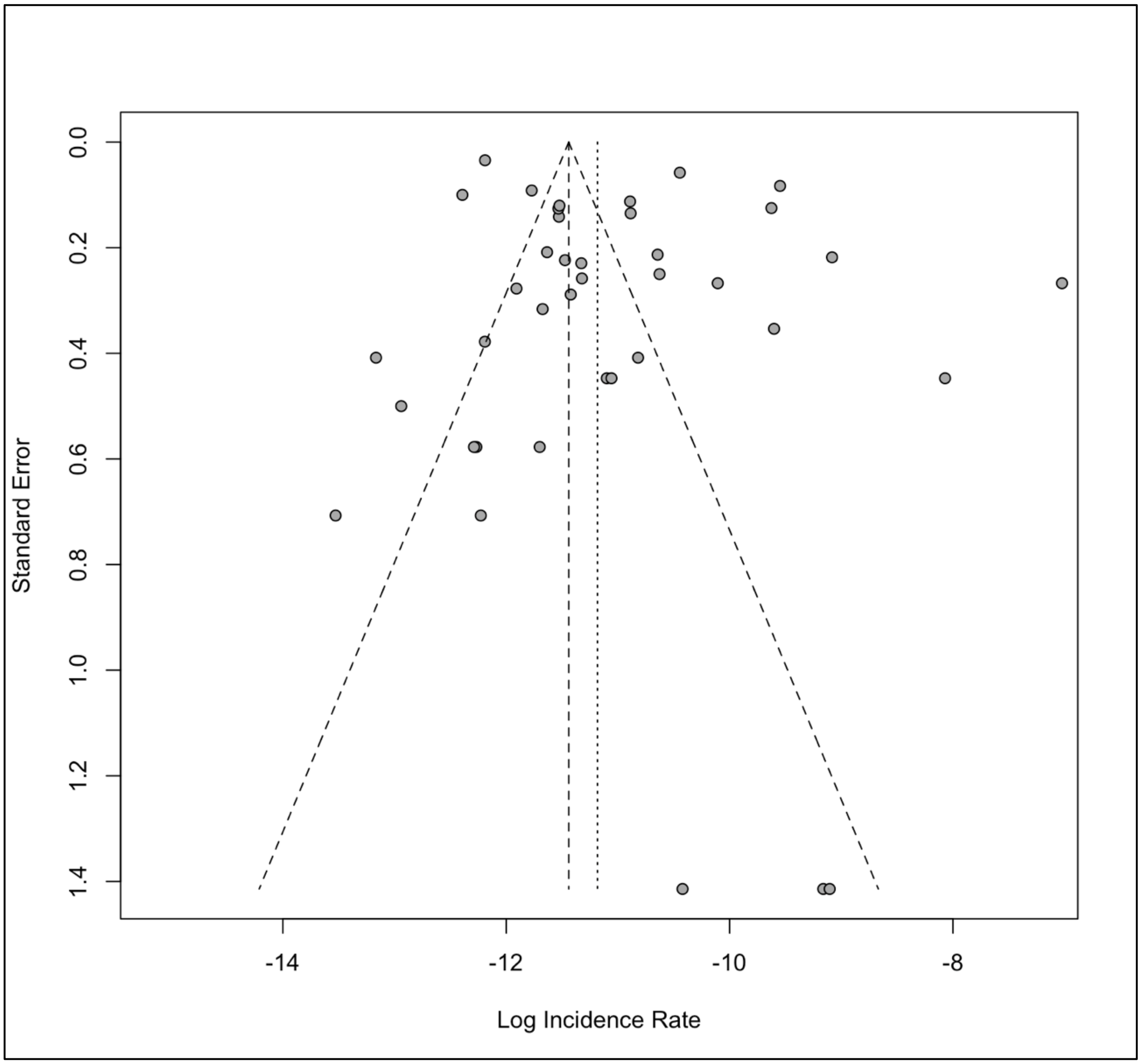

**Figure S18 :** Funnel plot for publication bias of included incidence studies reporting SCD

#### References

1. Drezner JA, Sharma S, Baggish A, et al. International criteria for electrocardiographic interpretation in athletes: Consensus statement. *Br J Sports Med*. 2017;51(9):704-731. doi:10.1136/bjsports-2016-097331
2. Harmon KG, Drezner JA, Wilson MG, Sharma S. Incidence of sudden cardiac death in athletes: A state-of-the-art review. *Br J Sports Med*. 2014;48(15):1185-1192. doi:10.1136/bjsports-2014-093872
3. Higgins JPT, Green S, eds. *Cochrane Handbook for Systematic Reviews of Interventions*. Chichester, West Sussex, England: The Cochrane Collaboration and John Wiley and Sons; 2009.
4. Higgins JP, Green S, eds. *Cochrane Handbook for Systematic Reviews of Interventions*. 5.1.0. Cochrane Collaboration; 2011. www.handbook.cochrane.org.
5. Wells G, Shea B, O'connell J, et al. The Newcastle-Ottawa Scale (NOS) for assessing the quality of nonrandomised studies in meta-analysis.
6. Hoy D, Brooks P, Woolf A, et al. Assessing risk of bias in prevalence studies: Modification of an existing tool and evidence of interrater agreement. *J Clin Epidemiol*. 2012;65(9):934-939. doi:10.1016/j.jclinepi.2011.11.014
7. Van Camp SP, Bloor CM, Mueller FO, Cantu RC, Olson HG. Nontraumatic sports death in high school and college athletes. / Mort survenant dans la pratique de sports non traumatisants chez des athletes de niveau college et dans les etablissements d ' enseignement secondaire. *Med Sci Sport Exerc*. 1995;27(5 PG-641-647):641-647.

http://articles.sirc.ca/search.cfm?id=375751

http://search.ebscohost.com/login.aspx?direct=true&AuthType=ip,cookie,url,uid&db=s3h&AN=SPH375751&site=ehost-live NS -.

8. Maron BJ, Haas TS, Murphy CJ, Ahluwalia A, Rutten-Ramos S. Incidence and causes of sudden death in U.S. college athletes. *J Am Coll Cardiol*. 2014;63(16):1636-1643. doi:10.1016/j.jacc.2014.01.041

9. Harmon KG, Asif IM, Maleszewski JJ, et al. Incidence and Etiology of Sudden Cardiac Arrest and Death in High School Athletes in the United States. *Mayo Clin Proc*. 2016;91(11 PG-1493-1502):1493-1502. http://resolver.ebscohost.com/openurl?sid=OVID:medline&id=pmid:27692971&id=doi:10.1016%2Fj.mayocp.2016.07.021&issn=0025-6196&isbn=&volume=91&issue=11&spage=1493&date=2016&title=Mayo+Clinic+Proceedings&atitle=Incidence+and+Etiology+of+Sudden+Cardiac+Arrest.

10. Toresdahl BG, Rao AL, Harmon KG, Drezner JA. Incidence of sudden cardiac arrest in high school student athletes on school campus. *Hear Rhythm*. 2014;11(7 PG-1190-4):1190-1194. http://resolver.ebscohost.com/openurl?sid=OVID:medline&id=pmid:24732370&id=doi:10.1016%2Fj.hrthm.2014.04.017&issn=1547-5271&isbn=&volume=11&issue=7&spage=1190&date=2014&title=Heart+Rhythm&atitle=Incidence+of+sudden+cardiac+arrest+in+high+school+student+at.

11. Berge HM, Andersen TE, Bahr R. Cardiovascular incidents in male professional football players with negative preparticipation cardiac screening results: an 8-year follow-up. *Br J*

*Sports Med*. October 2018:bjsports-2018-099845. doi:10.1136/bjsports-2018-099845

12. Maron BJ, Haas TS, Ahluwalia A, Rutten-Ramos SC. Incidence of cardiovascular sudden deaths in Minnesota high school athletes. *Hear Rhythm*. 2013;10(3):374-377.

13. Maron BJ, Gohman TE, Aeppli D. Prevalence of sudden cardiac death during competitive sports activities in Minnesota high school athletes. *J Am Coll Cardiol*. 1998;32(7 PG-1881-4):1881-1884. http://resolver.ebscohost.com/openurl?sid=OVID:medline&id=pmid:9857867&id=doi:&issn=0735-1097&isbn=&volume=32&issue=7&spage=1881&date=1998&title=Journal+of+the+American+College+of+Cardiology&atitle=Prevalence+of+sudden+cardiac+death+during+competitive+spo.

14. Chevalier L, Hajjar M, Douard H, et al. Sports-related acute cardiovascular events in a general population: a French prospective study. *Eur J Cardiovasc Prev Rehabil*. 2009;16(3 PG-365-70):365-370. http://resolver.ebscohost.com/openurl?sid=OVID:medline&id=pmid:19318955&id=doi:10.1097%2FHJR.0b013e3283291417&issn=1741-8267&isbn=&volume=16&issue=3&spage=365&date=2009&title=European+Journal+of+Cardiovascular+Prevention+%26+Rehabilitation&atitle=Sports-r.

15. Malhotra A, Dhutia H, Finocchiaro G, et al. Outcomes of Cardiac Screening in Adolescent Soccer Players. *N Engl J Med*. 2018;379(6):524-534.

16. Jordaens L, Van Lierde J, Van De Velde D, De Backer G. Competitive cyclging in Belgium is associated with an increased risk of sudden death. *Pacing Clin Electrophysiol*. 1996;19(4

PG-602-603):602-603.

http://search.ebscohost.com/login.aspx?direct=true&AuthType=ip,cookie,url,uid&db=s3h&AN=17417795&site=ehost-live NS -.

17. Biasco L, Cristoforetti Y, Castagno D, et al. Clinical, electrocardiographic, echocardiographic characteristics and long-term follow-up of elite soccer players with J-point elevation. *Circ Arrhythmia Electrophysiol*. 2013;6(6):1178-1184.

18. Santos-Lozano A, Martin-Hernandez J, Baladron C, et al. Sudden Cardiac Death in Professional Soccer Players. *J Am Coll Cardiol*. 2017;70(11 PG-1420-1421):1420-1421. doi:10.1016/j.jacc.2017.07.738

19. Drezner JA, Rogers KJ, Zimmer RR, Sennett BJ. Use of automated external defibrillators at NCAA Division I universities. *Med Sci Sport Exerc*. 2005;37(9 PG-1487-92):1487-1492. http://resolver.ebscohost.com/openurl?sid=OVID:medline&id=pmid:16177599&id=doi:&issn=0195-9131&isbn=&volume=37&issue=9&spage=1487&date=2005&title=Medicine+%26+Science+in+Sports+%26+Exercise&atitle=Use+of+automated+external+defibrillators+at+NCAA+Division+.

20. Harmon KG, Asif IM, Maleszewski JJ, et al. Incidence, Cause, and Comparative Frequency of Sudden Cardiac Death in National Collegiate Athletic Association Athletes: A Decade in Review. *Circulation*. 2015;132(1 PG-10-9):10-19. http://resolver.ebscohost.com/openurl?sid=OVID:medline&id=pmid:25977310&id=doi:10.1161%2FCIRCULATIONAHA.115.015431&issn=0009-7322&isbn=&volume=132&issue=1&spage=10&date=2015&title=Circulation&atitle=Inci

dence%2C+Cause%2C+and+Comparative+Frequency+of+Sudde.

21. Fuller CM, McNulty CM, Spring DA, et al. Prospective screening of 5,615 high school athletes for risk of sudden cardiac death. *Med Sci Sport Exerc*. 1997;29(9):1131-1138.

22. Drezner JA, Rao AL, Heistand J, Bloomingdale MK, Harmon KG. Effectiveness of emergency response planning for sudden cardiac arrest in United States high schools with automated external defibrillators. *Circulation*. 2009;120(6 PG-518-25):518-525. http://resolver.ebscohost.com/openurl?sid=OVID:medline&id=pmid:19635968&id=doi:10.1161%2FCIRCULATIONAHA.109.855890&issn=0009-7322&isbn=&volume=120&issue=6&spage=518&date=2009&title=Circulation&atitle=Effectiveness+of+emergency+response+planning+for+sudden.

23. Roberts WO, Stovitz SD. Incidence of sudden cardiac death in Minnesota high school athletes 1993-2012 screened with a standardized pre-participation evaluation. *J Am Coll Cardiol*. 2013;62(14):1298-1301.

24. Peterson DF, Kucera K, Thomas LC, et al. Aetiology and incidence of sudden cardiac arrest and death in young competitive athletes in the USA: A 4-year prospective study. *Br J Sports Med*. 2020:1-9. doi:10.1136/bjsports-2020-102666

25. Kucera KL, Klossner D, Colgate B, Cantu RC. *Annual Survey of Football Injury Research*. Chapel Hill, NC: University of North Carolina, Chapel Hill; 2018. NS -.

26. Boden BP, Breit I, Beachler JA, Williams A, Mueller FO. Fatalities in high school and college football players. *Am J Sports Med*. 2013;41(5 PG-1108-16):1108-1116. http://resolver.ebscohost.com/openurl?sid=OVID:medline&id=pmid:23477766&id=doi:10.1177%2F0363546513478572&issn=0363-

5465&isbn=&volume=41&issue=5&spage=1108&date=2013&title=American+Journal+of+Sports+Medicine&atitle=Fatalities+in+high+school+and+college+fo.

27. Landry CH, Allan KS, Connelly KA, et al. Sudden Cardiac Arrest during Participation in Competitive Sports. *N Engl J Med*. 2017;377(20 PG-1943-1953):1943-1953. http://resolver.ebscohost.com/openurl?sid=OVID:medline&id=pmid:29141175&id=doi:10.1056%2FNEJMoa1615710&issn=0028-4793&isbn=&volume=377&issue=20&spage=1943&date=2017&title=New+England+Journal+of+Medicine&atitle=Sudden+Cardiac+Arrest+during+Participation+in.

28. Steinvil A, Chundadze T, Zeltser D, et al. Mandatory electrocardiographic screening of athletes to reduce their risk for sudden death proven fact or wishful thinking? *J Am Coll Cardiol*. 2011;57(11):1291-1296.

29. Maron BJ, Doerer JJ, Haas TS, Estes NA, Hodges JS, Link MS. Commotio cordis and the epidemiology of sudden death in competitive lacrosse. *Pediatrics*. 2009;124(3 PG-966-71):966-971. http://resolver.ebscohost.com/openurl?sid=OVID:medline&id=pmid:19706581&id=doi:10.1542%2Fpeds.2009-0167&issn=0031-4005&isbn=&volume=124&issue=3&spage=966&date=2009&title=Pediatrics&atitle=Commotio+cordis+and+the+epidemiology+of+sudden+death+in+competitive.

30. Maurice MF, Di Tommaso F, Barros Pertuz MC, Mendoza WA, Spagnuolo D, Lucas V. Sudden cardiac death in rugby clubs. *Rev Argent Cardiol*. 2018;86(1 PG-40-44):40-44. http://resolver.ebscohost.com/openurl?sid=OVID:embase&id=pmid:&id=doi:10.7775%2Frac.v86.i1.12263&issn=0034-

7000&isbn=&volume=86&issue=1&spage=40&date=2018&title=Revista+Argentina+de+Cardiologia&atitle=Muerte+subita+en+clubes+deportivos+de+rugby&aulast=Mau.

31. Maron BJ, Haas TS, Ahluwalia A, Murphy CJ, Garberich RF. Demographics and Epidemiology of Sudden Deaths in Young Competitive Athletes: From the United States National Registry. *Am J Med*. 2016;129(11 PG-1170-1177):1170-1177. http://resolver.ebscohost.com/openurl?sid=OVID:medline&id=pmid:27039955&id=doi:10.1016%2Fj.amjmed.2016.02.031&issn=0002-9343&isbn=&volume=129&issue=11&spage=1170&date=2016&title=American+Journal+of+Medicine&atitle=Demographics+and+Epidemiology+of+Sudden+D.

32. Sterne JAC, Sutton AJ, Ioannidis JPA, et al. Recommendations for examining and interpreting funnel plot asymmetry in meta-analyses of randomised controlled trials. *BMJ*. 2011;343(7818):1-8. doi:10.1136/bmj.d4002
